# Cross-dataset Evaluation of Dementia Longitudinal Progression Prediction Models

**DOI:** 10.1101/2024.11.18.24317513

**Authors:** Chen Zhang, Lijun An, Naren Wulan, Kim-Ngan Nguyen, Csaba Orban, Pansheng Chen, Christopher Chen, Juan Helen Zhou, Keli Liu, B.T. Thomas Yeo, the Alzheimer’s Disease Neuroimaging Initiative, the Australian Imaging Biomarkers and Lifestyle Study of Aging

## Abstract

**Introduction:** Accurately predicting Alzheimer’s Disease (AD) progression is useful for clinical care. The 2019 TADPOLE (The Alzheimer’s Disease Prediction Of Longitudinal Evolution) challenge evaluated 92 algorithms from 33 teams worldwide. Unlike typical clinical prediction studies, TADPOLE accommodates (1) variable number of observed timepoints across patients, (2) missing data across modalities and visits, and (3) prediction over an open-ended time horizon, which better reflects real-world data. However, TADPOLE only used the Alzheimer’s Disease Neuroimaging Initiative (ADNI) dataset, so how well top algorithms generalize to other cohorts remains unclear.

**Methods:** We tested five algorithms in three external datasets covering 2,312 participants and 13,200 timepoints. The algorithms included FROG, the overall TADPOLE winner, which utilized a unique Longitudinal-to-Cross-sectional (L2C) transformation to convert variable-length longitudinal histories into feature vectors of the same length across participants (i.e., same-length feature vectors). We also considered two FROG variants. One variant unified all XGBoost models from the original FROG with a single feedforward neural network (FNN), which we referred to as L2C-FNN. We also included minimal recurrent neural networks (MinimalRNN), which was ranked second at publication time, as well as AD Course Map (AD-Map), which outperformed MinimalRNN at publication time. All five models – three FROG variants, MinimalRNN and AD-Map – were trained on ADNI and tested on the external datasets.

**Results:** L2C-FNN performed the best overall. In the case of predicting cognition and ventricle volume, L2C-FNN and AD-Map were the best. For clinical diagnosis prediction, L2C-FNN was the best, while AD-Map was the worst. L2C-FNN also maintained its edge over other models, regardless of the number of observed timepoints, and regardless of the prediction horizon from 0 to 6 years into the future.

**Conclusions:** L2C-FNN shows strong potential for both short-term and long-term dementia progression prediction. Pretrained ADNI models are available: https://github.com/ThomasYeoLab/CBIG/tree/master/stable_projects/predict_phenotypes/Zhang2025_L2CFNN.

## 1 Introduction

Alzheimer’s disease (AD) is a devastating neurodegenerative disorder (Jack et al., 2018; Hampel et al., 2021), which progresses over many years from a preclinical phase to a fully manifested clinical syndrome (Villemagne et al., 2013). There is no cure for AD, although medications exist to slow down cognitive decline in early AD (Van Dyck et al., 2023). The growing consensus is that early intervention is critical for slowing or stopping disease progression (Dubois et al., 2016; Scheltens et al., 2016; Aisen et al., 2022), and for enabling earlier caregiver planning (de Vugt & Verhey, 2013; Rasmussen & Langerman, 2019; Porsteinsson et al., 2021) as well as more efficient clinical trial enrollment (Burns et al., 2021; Oxtoby et al., 2022). Therefore, predicting AD progression is an important task (Zhang et al., 2017; Venkatraghavan et al., 2019). The Alzheimer’s Disease Prediction Of Longitudinal Evolution (TADPOLE) challenge^1^ (Marinescu et al., 2019, 2021) evaluated 92 algorithms from 33 teams worldwide for predicting AD progression. Given multimodal biomarkers at one or more timepoints, the goal was to predict cognition, ventricular volume and clinical diagnosis of an individual every month into the future. The TADPOLE challenge was run only on the Alzheimer’s Disease Neuroimaging Initiative (ADNI) dataset, so the current study seeks to evaluate the cross-dataset performance of several strong algorithms.

There are three key differences between the TADPOLE setup and most existing AD prediction studies. First, many studies use data from only a fixed number of timepoints (often just one) to predict future outcomes (Hebling Vieira et al., 2022; Hett et al., 2021; Wang et al., 2022). In practice, patients have varying numbers of visits, e.g., Patient A has three visits, while Patient B has only one. Limiting analyses to a single timepoint to accommodate Patient B wastes valuable information from Patient A. In contrast, TADPOLE encourages algorithms to leverage the full visit histories, which vary in length across participants. Second, many studies restrict their analyses to participants with complete multimodal data at all timepoints (Golovanevsky et al., 2022; Wang et al., 2022; Reas et al., 2023). But in clinical settings, not every patient undergoes every assessment (e.g., imaging, cognitive testing) at every visit. Discarding timepoints with incomplete data overlooks useful information. TADPOLE instead promotes using all available time points, even when some modalities are missing. Third, most studies predict a single outcome within a predefined window. For example, a typical mild cognitive impairment (MCI) progression task is to predict whether an MCI patient will develop dementia within three years (Basaia et al., 2019; El-Sappagh et al., 2021; Ocasio & Duong, 2021). The TADPOLE challenge, by contrast, requires forecasting monthly outcomes indefinitely into the future, which could then be flexibly used to answer specific clinical questions, such as the MCI progression task. Overall, unlike most existing studies, TADPOLE accommodates (1) a variable number of observed timepoints per patient, (2) missing data across modalities and visits, and (3) prediction over an open-ended time horizon. These features make it better aligned with the complexities of real-world clinical prediction.

There are two classes of existing algorithms naturally suited for the TADPOLE setup. The first class of models is dynamical state models, such as recurrent neural networks (RNNs; Ghazi et al., 2019; Nguyen et al., 2020; Jung et al., 2021; Liang et al., 2021; Xu et al., 2022; Cheng et al., 2024), where an individual’s latent state is represented by a vector, thus providing a rich encoding of an individual’s “disease state” beyond a single integer (such as in discrete state hidden Markov models). At each timepoint, observations are used to update the latent state of the individual at that timepoint. The latent state is in turn used to predict observations at the next time point. If some (or even all) observations are missing at the next time point, the model predictions can be used to fill in the missing data. Therefore, predicting missing data and future disease progression are unified into a single prediction task. This “model-filling” strategy was introduced by the MinimalRNN (Nguyen et al., 2020) and adopted by several subsequent studies (Jung et al., 2021; Liang et al., 2021; Xu et al., 2022; Cheng et al., 2024). MinimalRNN was evaluated by the TADPOLE organizers on their non-public ADNI test set, and was found to rank second at publication time (Nguyen et al., 2020). As such, the current benchmarking study included MinimalRNN because it is an influential algorithm whose performance has been evaluated in truly unseen data.

The second class of models is inspired by theoretical models of sigmoidal evolution of AD biomarkers (Jack et al., 2010; Villemagne et al., 2013; Selkoe & Hardy, 2016). These approaches fit parametric sigmoid-like functions to longitudinal biomarkers. Mixed-effects models capture group-level trends as fixed effects and individual variations as random effects (Iddi et al., 2019; Li, Iddi, et al., 2019; Oxtoby, 2023). Jedynak and colleagues used dynamic time-warping to align individual biomarker data to a group template represented by sigmoid curves (Jedynak et al., 2012). A Bayesian extension incorporated individual-specific latent time shifts (Bilgel et al., 2019). This line of research was extended to fit a constrained generalized sigmoidal function (Ghazi et al., 2021). A disease course mapping framework integrating Riemannian geometry and mixed-effects modeling with time reparameterization, known as AD course map (AD-Map; Koval et al., 2021) was shown to outperform 56 TADPOLE algorithms, including MinimalRNN, for predicting cognition (Maheux et al., 2023). Given the promise of the AD-Map algorithm, we also included AD-Map in the current benchmarking study.

In contrast to previous algorithms, the TADPOLE winner FROG utilized a longitudinal-to-cross-sectional (L2C) transformation (Nanopoulos et al., 2001; Deng et al., 2013; Barandas et al., 2020) to convert the longitudinal data into a cross-sectional format. Although the number of input features was initially different across participants (because of varying number of visits), the number of features was the same for all participants after L2C transformation (i.e., same-length input features). An important benefit of L2C transformation is that many powerful algorithms (e.g., random forest, support vector machines, etc) cannot be applied to variable-length input features, but could now be applied to the same-length L2C features. In the case of FROG, XGBoost (eXtreme Gradient Boost; Chen & Guestrin, 2016) was used to predict disease progression with the L2C features. The L2C transformation is relatively unique in the medical imaging community. There have been studies that converted longitudinal MRI histories into brain atrophy rates (Jack et al., 2004; Chincarini et al., 2016), but FROG’s L2C transformation captured a richer set of temporal statistics, such as past maximum and past minimum, in addition to rate of change (see Section 2.6). A final benefit of the L2C transformation was the amount of missing data was greatly reduced (see Section 2.6). In the TADPOLE challenge, FROG was the best for predicting clinical diagnosis and the overall winner. However, FROG has only been tested in the ADNI dataset, so it remains unclear how well it generalizes to new datasets.

In the current study, we trained the FROG algorithm on the ADNI dataset and evaluated its performance in three external datasets comprising 2312 participants with 13200 timepoints from the United States, Australia and Singapore. In the original FROG algorithm, separate XGBoost models were trained for different forecast windows and target variables. Here we considered a FROG variant that unified all XGBoost models with a single XGBoost model. Another FROG variant unified all XGBoost models with a single feedforward neural network (FNN) model, which we refer to as L2C-FNN. We also compared the FROG variants with a strong parametric modeling algorithm AD-Map, and a strong dynamic state algorithm MinimalRNN.

## 2 Methods

### 2.1 Problem overview

The problem setup followed the TADPOLE challenge (Marinescu et al., 2019, 2021). Given multimodal biomarkers and diagnostic history (Table 1) at one or more timepoints of an individual, we aimed to predict the cognitive state, ventricle volume normalized by intracranial volume (ICV), and clinical diagnosis of the individual for every subsequent month beyond the last observed timepoint up to 120 months into the future. Cognitive state was measured with the Alzheimer’s Disease Assessment Scale Cognitive Subdomain (ADAS-Cog13) in the original TADPOLE challenge. However, not all the external datasets had ADAS-Cog13, so we switched to predicting mini mental state examination (MMSE) in this study.

**Table 1.** Features used in the current study. ICV: intracranial volume. MMSE: mini mental state examination. CDR: Clinical Dementia Rating scale. CN: cognitively normal. MCI: mild cognitive impairment. DEM: Dementia with various etiologies. Note that FROG variants use all features. AD-Map was not developed to handle categorical variables or covariates, so do not utilize any of the baseline features or clinical diagnosis (see Section 2.5 for more details). MinimalRNN was not developed to handle covariates, so do not utilize any baseline features and age (see Section 2.4 for more details).

|  |  |  |
| --- | --- | --- |
| **Baseline features** | **Sex (male, female), Genetics (number of APOE-ε4 allele), Marital status (married, not married), Education level (number of years of education)** |  |
| **Recurring features** | MRI features | Hippocampus, Fusiform, Middle Temporal, Ventricle, Whole Brain, ICV |
|  | Cognitive features | MMSE (0-30), CDR GLOBAL (0, 0.5, 1, 2, 3) |
|  | Diagnostic features | Clinical diagnosis (CN, MCI, DEM) |
|  | Demographics | Age |

We used four datasets: the Alzheimer’s Disease Neuroimaging Initiative (ADNI) dataset, the Australian Imaging Biomarkers and Lifestyle Study of Ageing (AIBL) dataset, the Memory, Ageing and Cognition Centre (MACC) Harmonization Cohort, and the Open Access Series of Imaging Studies (OASIS) dataset. These datasets consisted of longitudinal multimodal data, such as T1-weighted structural MRI data, cognitive measurements and clinical diagnosis, as well as baseline demographics. The diagnostic categories corresponded to cognitively normal (CN), mild cognitive impairment (MCI), as well as dementia with various etiologies (DEM). Data collection was approved by the Institutional Review Board (IRB) at each corresponding institution. The analysis in the current study is approved by the National University of Singapore IRB.

### 2.2 Datasets, preprocessing and participant selection

#### 2.2.1 Datasets

The ADNI dataset (Jack et al., 2008; Petersen et al., 2010) is a comprehensive multicenter research initiative in the United States with three phases: ADNI1 (2004-2009), ADNI-GO/2 (2010-2016), and ADNI3 (2017-2023), with a primary focus on advancing the understanding of Alzheimer’s disease dementia. Each phase incorporates newly enrolled participants and individuals transitioning from earlier phases. Notably, there are variations in MRI scanner models and protocols across the phases, with ADNI1 primarily employing 1.5T scanners and subsequent phases adopting 3T scanners (see Table S1 for details). T1 images were downloaded from the USC Laboratory of Neuro Imaging’s Image and Data Archive (IDA). The ADNIMERGE spreadsheet (the ADNI team, 2023) containing various phenotypic data (e.g., demographics, clinical diagnoses, cognitive measurements) was also downloaded.

The AIBL study (Fowler et al., 2021) is an Australian flagship initiative that shares a similar goal and technical infrastructure with ADNI. MRI scans were acquired using both 1.5T and 3T (Avanto, Tim Trio and Verio) scanners (see Table S2 for details). MRI scans and phenotypic data were obtained from the USC Laboratory of Neuro Imaging’s Image and Data Archive (IDA).

The MACC Harmonization dataset (Hilal et al., 2020) focuses on a memory clinic population in Singapore. T1 images in this dataset were acquired exclusively using 3T (Tim Trio and Prisma) scanners (see Table S3 for details). Note that this dataset contained participants with vascular dementia and/or Alzheimer’s Disease dementia, which we have grouped together as Vascular/Alzheimer’s disease dementia (DEM). The mixed pathology allowed us to evaluate the generalizability of these models beyond AD dementia. However, for completeness, we will also report results for only AD dementia.

The OASIS dataset (LaMontagne et al., 2019) serves as a multimodal resource for studying normal aging and cognitive decline. It consists of four releases: OASIS-1 (cross-sectional) and OASIS-2 (longitudinal) as smaller-scale studies, OASIS-3 as the primary large dataset that includes OASIS-1 and OASIS-2 subjects, and OASIS-4, which encompasses a separate clinical cohort. For this study, we utilized OASIS-3 data. Note that this dataset included both AD and non-AD dementia, which we have grouped together as DEM (similar to MACC). However, for completeness, we will also report results for only AD dementia. The imaging data in OASIS were acquired using both 1.5T and 3T scanners (see Table S4 for details). MRI scans and phenotypic data were downloaded from the XNAT Central (Herrick et al., 2016).

#### 2.2.2 Preprocessing

All T1 images were de-obliqued and reoriented to the RPI orientation. Subsequently, we used the FreeSurfer 6.0 recon-all pipeline (Fischl et al., 2002; Desikan et al., 2006) to derive the volumes of various regions of interest (ROIs). Following the FROG algorithm, we derived five brain ROI volumetric features associated with AD-dementia, namely Hippocampal volume, Fusiform volume, Middle Temporal (MidTemp) volume, Ventricle volume, and Whole Brain volume (see Table S5 for details). We also incorporated ICV as an additional feature and standardized the five brain ROI features with respect to ICV (Table 1).

The generated brain ROI features were then merged with downloaded phenotypic data (e.g., demographics, clinical diagnoses, cognitive measurements). Notably, non-MRI phenotypes (e.g., clinical diagnoses and cognitive measurements) and MRI scans might not have been performed on the same day. Following the ADNIMERGE convention, if the non-MRI and MRI dates were within 6 months of each other, then the non-MRI and MRI phenotypes were merged into one timepoint corresponding to the non-MRI date.

We systematically eliminated empty or duplicate entries, along with those displaying outliers or errors. Certain datasets used inconsistent coding for missing values, such as NaN or special integers (e.g., –1, –4, 999). To ensure consistency, we replaced all special integers with NaN. Consequently, we obtained a clean longitudinal data table where each row represents one timepoint of a participant, containing MRI features and/or cognitive features and/or diagnostic features (Table 1).

#### 2.2.3 Participant selection and characteristics

Our objective was to predict the longitudinal progression of dementia, so across all four datasets, we only included participants with recurring features (Table 1) at two or more timepoints. We note that under this criterion, the recurring features did not need to all occur in the same timepoints. For example, a participant with only MRI features in timepoint 1 and only cognitive features in timepoint 2 was considered acceptable.

With the above selection criterion, the final ADNI dataset comprised 2111 participants with a total of 15791 timepoints, including 9668 timepoints with MRI features. In the case of AIBL, the final dataset comprised 402 participants with a total of 1220 timepoints, including 940 timepoints with MRI features. In the case of MACC, the final dataset comprised 650 participants with a total of 3067 timepoints, including 1453 timepoints with MRI features. In the case of OASIS, the final dataset comprised 1260 participants with a total of 8913 timepoints, including 2519 timepoints with MRI scans.

The demographics, disease severity and number of timepoints vary significantly between ADNI and the three external datasets (Table 2). Actual distributions are plotted in Figures S1 and S2. Compared with ADNI, the AIBL participants were younger and had higher MMSE scores. There were also proportionally more female and CN participants in the AIBL dataset than the ADNI dataset. Compared with ADNI, the MACC participants had lower MMSE scores. Furthermore, there were proportionally more female and participants with DEM diagnosis in MACC than ADNI. Finally, compared with ADNI, the OASIS participants were younger and had higher MMSE scores. There were also proportionally more female and CN participants in the OASIS dataset than the ADNI dataset.

**Table 2.** Participant characteristics in the four datasets. Statistical tests were performed to compare ADNI and each external dataset. For continuous variables (e.g., age and MMSE), a permutation test was used. For discrete variables (e.g., sex and diagnosis), the chi square test was used. Bolded p values indicate statistical significance after correcting for multiple comparisons with false discovery rate (FDR) q < 0.05. Note that not all OASIS participants have clinical diagnosis at baseline, so the percentages of participants with baseline diagnosis do not add up to 100%.

|  | **ADNI**<br>**(N = 2111)** | **AIBL**<br>**(N = 402)** |  | **MACC**<br>**(N = 650)** |  | **OASIS**<br>**(N = 1260)** |  |
| --- | --- | --- | --- | --- | --- | --- | --- |
|  | **mean ± std** | **mean ± std** | **p** | **mean ± std** | **p** | **mean ± std** | **p** |
| **Baseline age (y)** | 73.3 ± 7.2 | 72.4 ± 6.7 | **2.9e-2** | 72.7 ± 7.9 | 7.8e-2 | 69.0 ± 9.0 | **1.0e-4** |
| **Baseline MMSE** | 27.4 ± 2.6 | 28.0 ± 2.8 | **1.0e-4** | 21.6 ± 6.0 | **1.0e-4** | 28.3 ± 2.5 | **1.0e-4** |
| **Sex (M/F)** | 1135 / 976<br>(54% / 46%) | 157 / 176<br>(39% / 44%) | **2.9e-2** | 286 / 364<br>(44% / 56%) | **1.6e-5** | 560 / 700<br>(44% / 56%) | **2.0e-7** |
| **Baseline diagnosis**<br>**(CN/MCI/DEM)** | 745 / 995 / 371<br>(35%/47%/18%) | 319 / 48 / 35<br>(79%/12%/9%) | **8.4e-60** | 131 / 272 / 247<br>(20%/42%/38%) | **2.7e-29** | 741 / 27 / 187<br>(59%/2%/15%) | **7.6e-138** |
| **Baseline DEM**<br>**(AD/Other dementias)** | - | - | - | 194 / 53<br>(79% / 21%) | - | 53 / 134<br>(28% / 72%) | - |
| **Cognitively Normal (CN)** |  |  |  |  |  |  |  |
| Baseline Age (y) | 73.0 ± 6.2 | 72.1 ± 6.4 | **2.5e-2** | 68.4 ± 7.5 | **1.0e-4** | 68.3 ± 8.4 | **1.0e-4** |
| Sex (M/F) | 333 / 412<br>(45% / 55%) | 125 / 148<br>(39% / 46%) | 8.1e-1 | 58 / 73<br>(44% / 56%) | 1.0 | 319 / 422<br>(43% / 57%) | 5.6e-1 |
| **Mild Cognitive Impairment (MCI)** |  |  |  |  |  |  |  |
| Baseline age (y) | 73.0 ± 7.5 | 75.2 ± 6.5 | 4.2e-2 | 72.9 ± 7.6 | 9.2e-1 | 71.8 ± 6.2 | 4.3e-1 |
| Sex (M/F) | 592 / 403<br>(59% / 41%) | 22 / 14<br>(46% / 29%) | 9.8e-1 | 130 / 142<br>(48% / 52%) | **7.1e-4** | 15 / 12<br>(56% / 44%) | 8.3e-1 |
| **Dementia (DEM)** |  |  |  |  |  |  |  |
| Baseline age (y) | 74.6 ± 7.8 | 71.8 ± 8.8 | 5.2e-2 | 74.7 ± 7.7 | 8.4e-1 | 74.2 ± 7.3 | 6.1e-1 |
| Sex (M/F) | 210 / 161<br>(57% / 43%) | 10 / 14<br>(29% / 40%) | 2.2e-1 | 98 / 149<br>(40% / 60%) | **5.3e-5** | 96 / 91<br>(51% / 49%) | 2.8e-1 |
| **Number of visits** | 7.5 ± 4.5 | 3.0 ± 1.3 | **1.0e-4** | 4.7 ± 1.4 | **1.0e-4** | 7.1 ± 4.7 | **1.7e-2** |
| **Follow-up duration (y)** | $4.5 \pm 3.4$ | $3.3 \pm 2.0$ | **1.0e-4** | $3.9 \pm 1.5$ | **1.0e-4** | $7.8 \pm 5.6$ | **1.0e-4** |
| **Percentage of timepoints with missing data for recurring features** |  |  |  |  |  |  |  |
| CDR Global | 28.6% | 0.5% | **2.7e-101** | 1.6% | **5.8e-224** | 5.1% | **0.0** |
| MMSE | 30.1% | 0.4% | **1.4e-109** | 1.7% | **7.6e-240** | 16.6% | **2.3e-122** |
| MRI features | 38.8% | 23.0% | **5.9e-28** | 52.6% | **4.6e-46** | 71.8% | **0.0** |
| Diagnosis | 30.2% | 0.4% | **5.8e-110** | 1.2% | **1.7e-249** | 19.2% | **5.3e-80** |

Furthermore, both the AIBL and MACC datasets typically have fewer than 7 timepoints collected within a span of 7 years. In contrast, some participants in the ADNI and OASIS datasets have up to 20 to 30 timepoints, covering a tracking period of over 15 years. The percentage of timepoints with missing data was also highly different across the datasets (Table 2).

### 2.3 Training, validation and test procedure

We compared five different models: MinimalRNN, AD-Map, original FROG (L2C-XGBw), FROG variant 1 (L2C-XGBnw) and FROG variant 2 (L2C-FNN). We trained different models using ADNI and evaluated their performance within the ADNI dataset (within-dataset evaluation) and in AIBL, MACC and OASIS datasets (cross-dataset evaluation). More specifically, we randomly divided the ADNI participants into 20 partitions. Since each participant contributed data from multiple timepoints, care was taken to assign all timepoints from the same participant to a single partition to prevent test set leakage. In other words, no participant’s data was split across multiple partitions.

To train a given model, 18 partitions were used for training, while 1 partition was used as a validation set to tune the hyperparameters. The remaining partition was used as test set to evaluate the within-cohort performance of the model. To ensure stability of results (Kong et al., 2019; Li, Kong, et al., 2019; Varoquaux, 2018), this procedure was repeated 20 times with a different partition being the test set (e.g., partition 5) and the partition next to it being the validation set (e.g., partition 6). Therefore, we ended up with 20 sets of trained models together with 20 sets of within-cohort evaluation results. The 20 sets of trained models were applied to all participants in AIBL, MACC, and OASIS for cross-cohort evaluation (Figure 1).

**Figure 1.**
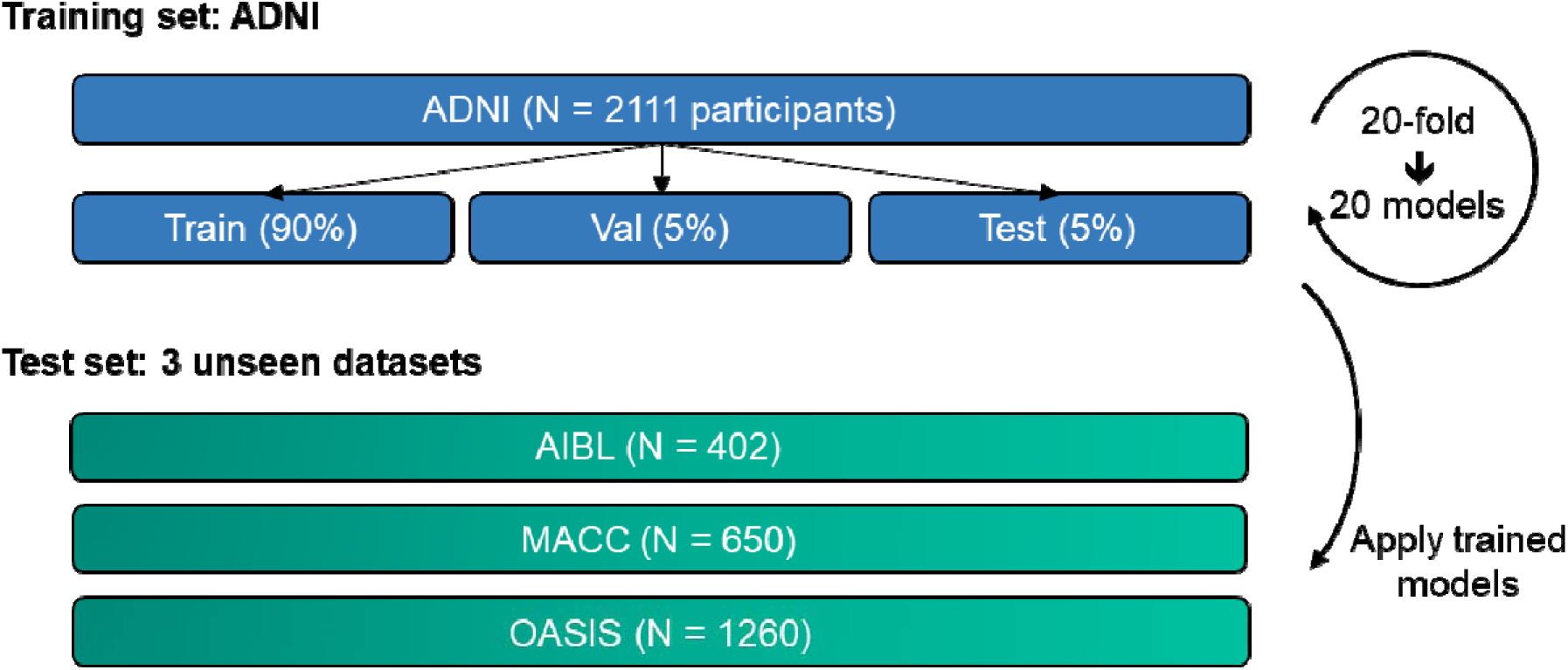
Training and testing procedure. All models were trained on the ADNI dataset and subsequently applied to three unseen test datasets to assess generalizability. ADNI participants were randomly divided into training, validation, and test sets (ratio of 18:1:1) for hyperparameter tuning and within-cohort evaluation. This procedure was repeated 20 times to ensure result stability. Care was taken to ensure non-overlapping test sets, covering the entirety of the ADNI dataset across the 20 data splits. Trained models were then evaluated on participants from the three unseen test datasets (AIBL, MACC, and OASIS) for cross-cohort evaluation.

Following TADPOLE convention, for participants in the ADNI validation and test sets, as well as all participants in AIBL, MACC, and OASIS, the first half of each participant’s timepoints were used to predict the second half of the same participant’s timepoints. For example, if a participant had 10 timepoints, then the first 5 timepoints were used as input (observed) timepoints, and we sought to predict the second 5 timepoints (unobserved). On the other hand, the entire longitudinal time series of training participants were used during training to increase data efficiency.

The Optuna library (Akiba et al., 2019) was utilized to find the best hyperparameters by maximizing model performance on the validation set. We note that this optimization was performed independently for each training/validation/test split of the dataset. The hyperparameter search spaces for each algorithm are described in their respective method sections.

### 2.4 MinimalRNN

MinimalRNN is a recurrent neural network (RNN) with less parameters than LSTM (Long short-term memory) to mitigate overfitting. In our previous study (Nguyen et al., 2020), we found that MinimalRNN performed better than the more complex LSTM, as well as a simpler linear state space model in the TADPOLE challenge. As such, the MinimalRNN struck the perfect complexity balance, yielding the best prediction performance among the RNN models we tested.

In RNNs, the same computational unit is repeated at each time step, where the output at the current step becomes the input at the next step. Therefore, the longitudinal data of a participant is analyzed sequentially, where the input features at a particular timepoint is used to update the internal “disease” state of the participant. This internal state is then used to predict the input features at the next time point.

For our experiments, we utilized the publicly available code from https://github.com/ThomasYeoLab/CBIG/tree/master/stable_projects/predict_phenotypes/Nguyen2020_RNNAD. However, we replaced the HORD hyperparameter search algorithm (Eriksson et al., 2019; Ilievski et al., 2017) employed in our previous study (Nguyen et al., 2020) with Optuna (Akiba et al., 2019) because of its ease of use.

Table 3 summarizes the hyperparameters optimized by Optuna and their corresponding search range. Consistent with the original study (Nguyen et al., 2020), MinimalRNN only utilized the recurring MRI features, cognitive features and clinical diagnosis (Table 1), but not any baseline features (sex, education, marital status and number of APOE-ε4) and age. Our previous experiments (not shown) found that these additional information did not improve prediction performance.

**Table 3.**
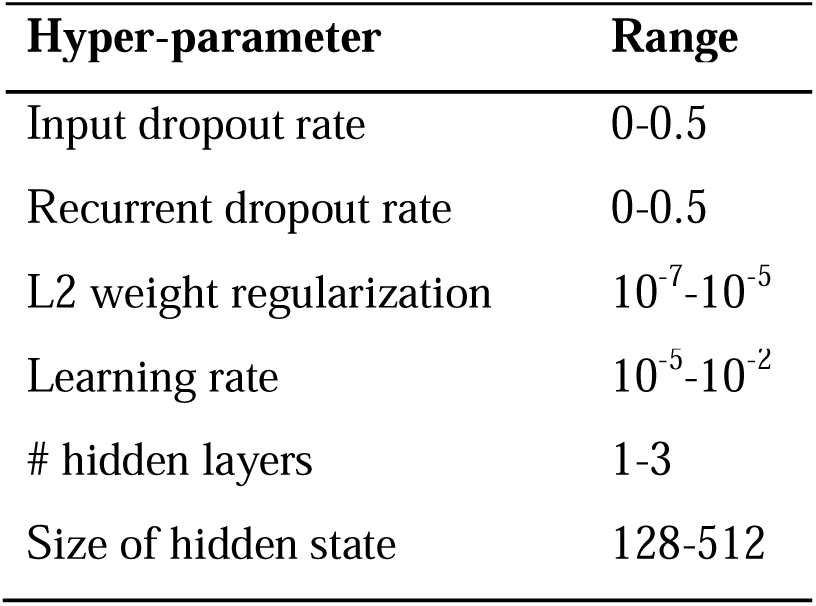
Hyper-parameters and corresponding search ranges for MinimalRNN estimated from the validation sets using Optuna.

### 2.5 AD Course Map (AD-Map)

AD-Map is a parametric Bayesian non-linear mixed-effects model designed to predict cognition and brain atrophy. It was shown to outperform MinimalRNN (Maheux et al., 2023). AD-Map assumes that each biomarker follows a logistic curve, with different biomarkers exhibiting distinct progression rates and ages at inflexion point. The model adjusts these curves for each individual by learning individual-specific shifts in disease onset, progression rates, and the timing/ordering of biomarker progression. As a result, the model predicts an individual-specific set of logistic curves, which show the value of each biomarker at any age of the participant.

For our experiments, we used the Leaspy software (https://gitlab.com/icm-institute/aramislab/leaspy) and optimized hyperparameters (Table 4) via Optuna (Akiba et al., 2019). We note that ICV was not included as a feature since AD-Map requires time-dependent features, and ICV shows minimal change with time (Courchesne et al., 2000; Jenkins et al., 2000). However, we remind the reader that the other MRI volumetric features were normalized with respect to ICV, consistent with other algorithms.

**Table 4.** Hyper-parameters and corresponding search ranges for AD-Map estimated from the validation sets using Optuna.

| Hyper-parameter | Range |
| --- | --- |
| Source dimension | 1-5 |
| # iterations | (1-10) * 500 |

Because AD-Map assumes that each biomarker follows a sigmoidal-like curve, categorical variables (e.g., clinical diagnosis) cannot be used as an input or be predicted with AD-Map (Maheux et al., 2023). To compare with models that predict clinical diagnosis, we converted AD-Map’s predicted CDR score into probabilities for CN, MCI, and DEM using standard cut-off points (O’Bryant et al., 2010; Tariot et al., 2024): CDR 0 was mapped to CN, CDR 0.5 was mapped to MCI, and CDR 1, 2, and 3 were mapped to DEM. For predicted CDR scores between 0 and 0.5, we used linear interpolation, so for instance, a CDR score of 0.1 resulted in 80% probability for CN, 20% probability for MCI, and 0% probability for DEM. For scores between 0.5 and 1, we again used linear interpolation, so for instance, a CDR score of 0.6 resulted in 0% probability for CN, and 80% probability for MCI, and 20% probability for DEM. CDR scores of 1 or higher are fully assigned to DEM.

We also explored the possibility of treating clinical diagnosis as a continuous variable by setting CN to be 0, MCI to be 1 and DEM to be 2. This allowed clinical diagnosis to be included as an input to AD-Map and also allowed diagnosis to be directly predicted. We found that this approach improved clinical diagnosis prediction but led to very poor MMSE and ventricular volume prediction. Therefore, consistent with the original study (Maheux et al., 2023), we did not include clinical diagnosis in the AD-Map algorithm. Instead, we used the posthoc CDR-to-Diagnosis conversion approach described above, which maintained AD-Map’s original focus on continuous biomarkers while allowing for comparisons of clinical diagnosis prediction.

Furthermore, the AD-Map package does not take in any baseline features (sex, education, marital status and number of APOE-ε4), so in summary, the AD-Map only used recurring MRI features (excluding ICV), cognitive features and age.

### 2.6 Original FROG: Longitudinal-to-Cross-sectional XGBoost with Windows (L2C-XGBw)

#### 2.6.1 Longitudinal-to-Cross-sectional (L2C) transformation

The TADPOLE problem set up is challenging for standard machine learning algorithms (e.g., support vector machine) because of the variable length of observed timepoints. The winner of the TADPOLE challenge FROG used a feature engineering technique (Nanopoulos et al., 2001; Deng et al., 2013; Barandas et al., 2020) that transformed the longitudinal visit history of participants into a cross-sectional format, which we will refer to L2C (Longitudinal-to-Cross-sectional) transformation.

More specifically, suppose for a given participant, we observed data at m timepoints t_l_, t_2_, t_3_, t_4_, …, t_m_, and we would like to predict clinical diagnosis (or MMSE or ventricular volume) at a future timepoint t_f_. Note that these timepoints might not be equally spaced in time. To convert the variable length input features, FROG proposed the following L2C transformation, in which each continuous input modality (i.e., MMSE, CDR_GLOBAL, six anatomical ROI volumes) is converted into seven features (Table 5) and clinical diagnosis is converted into eight features (Table 6), resulting in 8 x 7 + 8 = 64 features.

**Table 5.**
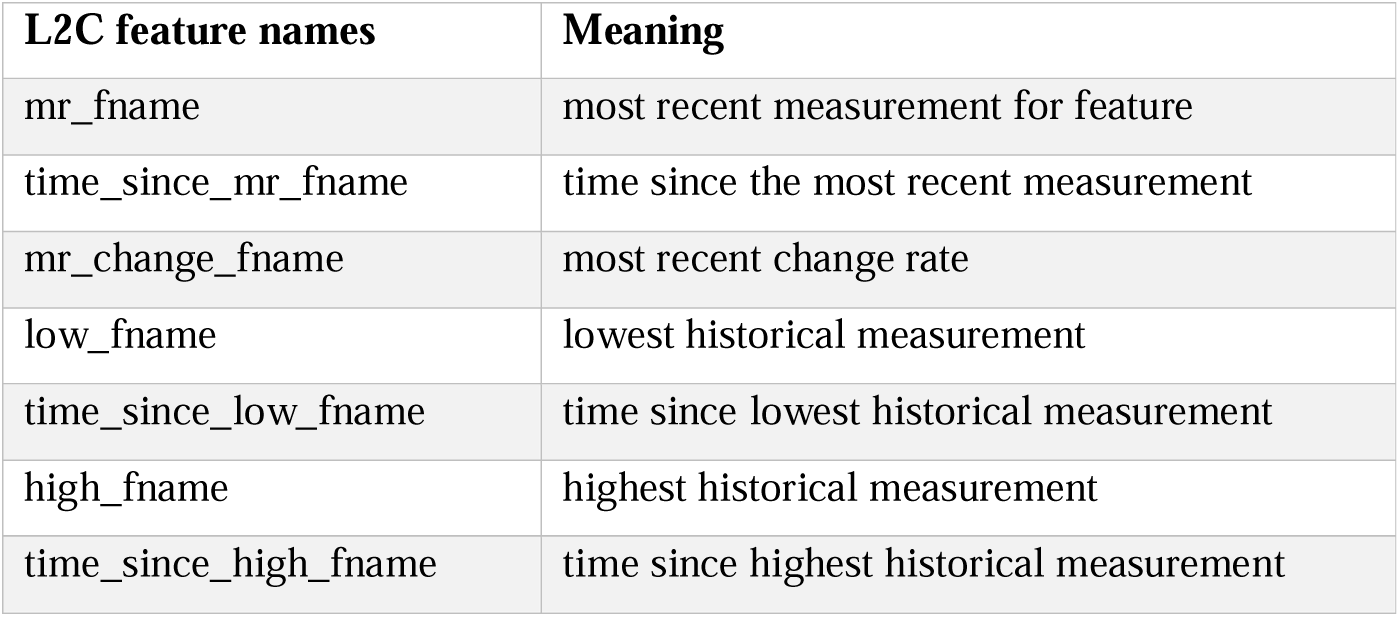
L2C feature names and their corresponding meaning for continuous input modalities.

| L2C feature names | Meaning |
| --- | --- |
| mr_fname | most recent measurement for feature |
| time_since_mr_fname | time since the most recent measurement |
| mr_change_fname | most recent change rate |
| low_fname | lowest historical measurement |
| time_since_low_fname | time since lowest historical measurement |
| high_fname | highest historical measurement |
| time_since_high_fname | time since highest historical measurement |

**Table 6.** L2C feature names and their corresponding meaning for clinical diagnosis.

| L2C feature names | Meaning |
| --- | --- |
| mr_dx | most recent non-missing diagnosis |
| time_since_mr_dx | time since the most recent non-missing diagnosis |
| best_dx | best historical diagnosis |
| time_since_best_dx | time since best historical diagnosis |
| worst_dx | worst historical diagnosis |
| time_since_worst_dx | time since worst historical diagnosis |
| milder | 1 if a milder diagnosis occurred in history |
| time_since_milder | time since the milder diagnosis and 999 if no milder diagnosis |

The 64 L2C features were additionally augmented by age at future timepoint t_f_, baseline sex, baseline education level, baseline marital status, APOE status and number of months between future timepoint t_f_ and the first (baseline) visit. In total, there were 64 + 6 = 70 features. Please refer to Supplementary Table S6 for the complete set of L2C features used by FROG.

#### 2.6.2 Theoretical benefits of the L2C transformation

We note that it is not the case that the L2C transformation necessarily reduced the number of features. For a participant with the full set of multimodal data on two visits, the L2C transformation would generate 70 features, while MinimalRNN and AD-Map would only have 9 × 2 = 18 and 8 × 2 = 16 features, respectively. For a participant with three complete visits, the L2C transformation would again generate 70 features, while MinimalRNN and AD-Map would only have 9 × 3 = 27 and 8 × 3 = 24 features, respectively.

In the above example, MinimalRNN had to deal with 18 features for a participant with two complete visits and 27 features for a participant with three complete visits. On the other hand, the L2C transformation always generates 70 features, regardless of whether a participant has one past visit or ten past visits. This is a key benefit of L2C transformation because a wide range of powerful machine learning models (e.g., random forests, support vector machines, etc) require the same number of features for each participant. In the case of FROG, the powerful XGBoost model was used (Section 2.6.4), while L2C-FNN utilized a fully connected feedforward network (Section 2.8). In other words, the L2C transformation gives the user great flexibility in choosing the algorithm to be applied to the L2C features.

The L2C features also captured key aspects of a time series, including temporal trends (i.e., most recent change rate) and extreme values (lowest and highest historical measurements), which are well-established summaries in time series feature extraction (Nanopoulos et al., 2001; Deng et al., 2013; Barandas et al., 2020). For example, a rapid decline in hippocampal volume or extremely low hippocampal volume might be predictive of progression to dementia. The inclusion of time-varying covariates (e.g., “time since the most recent measurement” or “time since lowest historical measurement”) are necessary to compensate for the loss of temporal information when collapsing the longitudinal history into cross-sectional features. For example, the lowest measured hippocampal volume would be more useful if we also knew how far in the past the measurement occurred.

While a broader set of transformations (e.g., standard deviation) could be considered, these were not explored in the TADPOLE challenge, when FROG was first developed. To maintain consistency with the original FROG algorithm and given the strong performance we already observed, we did not introduce additional transformations in the current study. However, we suspect that any sufficiently rich set of the time series summaries might be equally effective.

Another advantage of the L2C transformation is the reduction in missing data. For example, the average ADNI participant has 7.5 visits. Let us consider a participant with eight visits, and the first four visits were used to predict MMSE, ventricular volume and clinical diagnosis in the last four visits. Suppose MMSE was only measured in two of the four input timepoints. In the case of MinimalRNN, the missing MMSE values needed to be filled in by the MinimalRNN model (i.e., model filling). However, in the case of L2C transformation, all seven features associated with MMSE (Table 5) could be computed with just two MMSE measurements in the visit history. Even if there was only one MMSE measurement, there would only be one missing value out of the seven features associated with MMSE (“most recent change rate” in Table 5).

#### 2.6.3 Data augmentation

In addition to the L2C transformation, FROG proposed the following data augmentation strategy during training. Suppose we observed data at m timepoints t_l_, t_2_, t_3_, t_4_, …, t_m_ for a particular training participant. FROG then generated m-1 training samples by using t_l_ to predict t_2_, or t_3_ or t_4_ or t_S_ etc. FROG also generated another m-2 training samples by using t_l_ and t_2_ to predict t_3_ or t_4_ or t_S_ etc. In total, given m timepoints, FROG generated m * (m – 1)/2 training samples.

#### 2.6.4 L2C eXtreme Gradient Boost with separate windows (L2C-XGBw)

The L2C transformation converted variable length input features into same-length input features for each participant (Section 2.6.1), while the data augmentation procedure generated more training samples (Section 2.6.3). The original FROG team used eXtreme Gradient Boost (XGBoost; Chen & Guestrin, 2016) to predict the target variables from the L2C features. Gradient boosting is a model ensemble of individual decision trees that are trained sequentially such that a new tree improves the error of the previous tree ensemble. XGBoost is an optimized distributed gradient boosting library. The original FROG submission used the XGBoost R library, while we reimplemented the FROG algorithm in python. We performed 5 repetitions of train/validation/test split in the ADNI dataset to ensure our python implementation yields numerically the same results as the R code.

Furthermore, consistent with the original FROG submission to the TADPOLE challenge, we trained separate XGBoost models for each target variable (clinical diagnosis, MMSE, ventricle volume). Following the original FROG submission, we also trained separate models based on specific forecast interval ranges, with the assumption that certain models may excel in short-term predictions while others in long-term forecasts. The forecast interval ranges (i.e., forecast windows) for each target variable (measured in months) adhere to the FROG team’s settings (Table 7). Hence, we referred to this algorithm as L2C eXtreme Gradient Boost with separate windows (L2C-XGBw).

**Table 7.** L2C-XGBw (FROG) trained a separate XGBoost model for each forecast window and each target variable.

| Target variables | Forecast Windows (months) |
| --- | --- |
| MMSE | 0-9, 9-15, 15-27, 27-39, >54 |
| Clinical diagnosis | 0-8, 8-15, 15-27, 27-39, 39-60, >60 |
| Ventricle volume | 0-9, 9-15, 15-30, >30 |

Three important hyperparameters were tuned in the ADNI validation sets using Optuna (Akiba et al., 2019). The three hyperparameters and search ranges are detailed in Table 8. We note that there is no extra feature normalization or missing data imputation since the XGBoost package handles such issues internally.

**Table 8.**
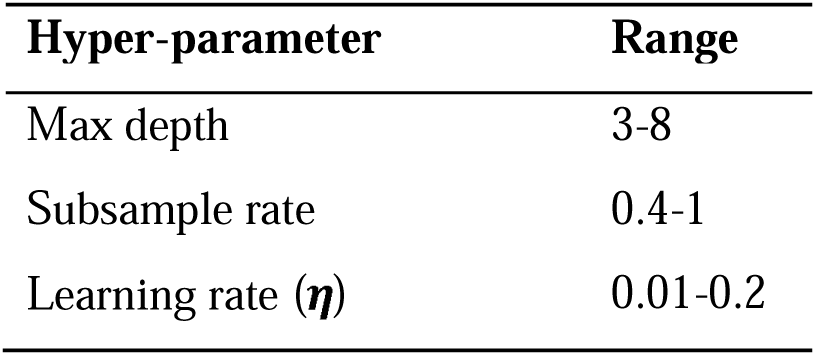
Hyper-parameters and corresponding search ranges for L2C-XGBw estimated from the validation sets using Optuna.

### 2.7 FROG variant 1: L2C eXtreme Gradient Boost with no window (L2C-XGBnw)

L2C-XGBw (FROG) involved training a separate XGBoost model for each forecast window. This is not ideal because the forecast windows are themselves hyperparameters, which might be hard to pick for new target variables. We hypothesized that the multiple forecast windows might not be necessary because L2C features like “time since baseline” and “time since most recent measurement” already encode the necessary temporal information for the model. Therefore, we considered a variant of FROG, where a single XGBoost model was trained for all future timepoints, as opposed to a separate model for each time window. We refer to this baseline as L2C eXtreme Gradient Boost with no window (L2C-XGBnw). All other implementation details remain consistent with those of L2C-XGBw.

### 2.8 FROG variant 2: L2C Fully-Connected Feedward Neural Network (L2C-FNN)

L2C-XGBnw trained a separate XGBoost model for each target variable. Previous studies have suggested that predicting multiple target variables can potentially improve prediction performance. By learning shared representations to capture common patterns among related tasks, these shared representations might enhance data efficiency, accelerate learning, and mitigate overfitting issues (Rahim et al., 2017; Crawshaw, 2020).

A natural choice to incorporate multi-task learning is to replace XGBoost with a fully connected feedforward neural network (FNN) model, with the output layer predicting all target variables jointly. Adding new target variables only increases the dimension of the output layer, which eliminates the need for separate models and simplifies the coding and hyperparameter tuning. Similar to L2C-XGBnw, we will train a single FNN model to predict all future timepoints, instead of the original FROG implementation which trained a separate XGBoost model for each forecast window. We will refer to this model as the L2C Fully-Connected Feedward Neural Network (L2C-FNN).

Figure 2 illustrates the L2C-FNN architecture. LeakyReLU (Maas, 2013) was chosen as the activation function, and dropout (Srivastava et al., 2014) was applied after each activation function to enhance model generalizability. The FNN output is a 5-dimensional vector: the first three elements represent the individual’s probabilities of being diagnosed as CN, MCI, or DEM at a future timepoint, computed using a SoftMax function, while the fourth and fifth elements correspond to MMSE and ventricle volume predictions.

**Figure 2.**
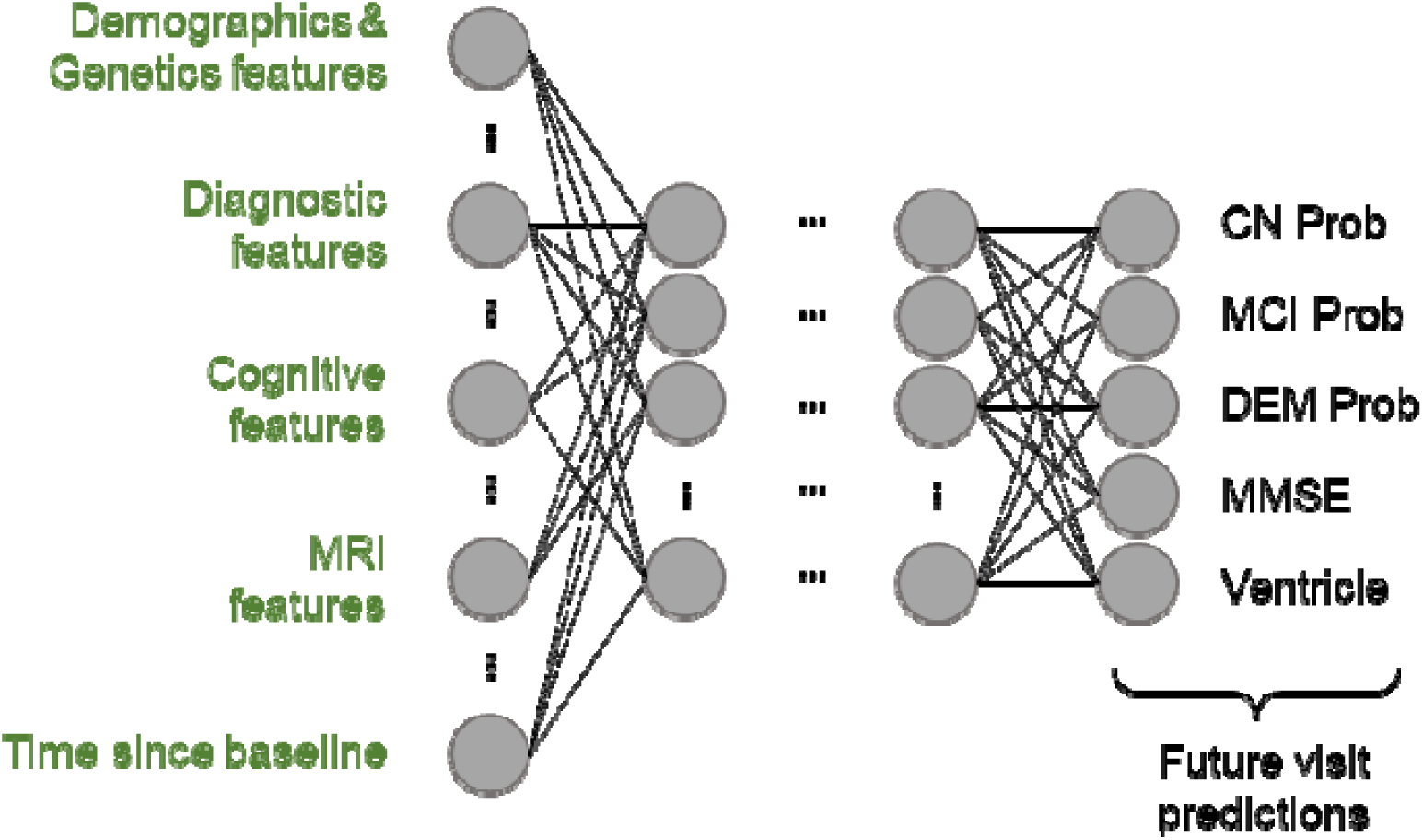
Architecture of the L2C Fully Connected Feedforward Neural Network (L2C-FNN). FNN incorporates leaky rectified linear units (LeakyReLU) between layers. The input layer comprises multimodal L2C features, among which “time since baseline” denotes the duration from the baseline visit to the timepoint we want to predict, aiding in longitudinal prediction. The final layer simultaneously outputs clinical diagnosis probabilities (calculated with a soft-max function), MMSE score, and ventricle volume for multi-task learning.

Input features are similar to L2C-XGBw (FROG), with additional preprocessing steps. Due to FNN sensitivity to input scale and missing data compared to tree-based models such as XGBoost, we performed Gauss Rank normalization, a special form of quantile normalization (Zhao et al., 2020), with a Gaussian reference distribution. The transformation was performed using the Scikit-learn quantile transform function (Pedregosa et al., 2011). Discrete features, such as APOE, sex, and most recent diagnosis (mr_dx) were encoded using one-hot encoding. To handle missing data, an “unknown” class was introduced for all discrete features, and missing values were assigned to this class. Numeric feature imputation involved replacing missing values with the median of the training set before Gauss Rank transformation. During inference, the learned Gauss Rank transformations and statistics from the training set were used to impute and transform validation and test data. Notably, the “time since milder clinical diagnosis” feature was removed due to its high proportion of NaN values (91.57%) which primarily stems from two causes: either because there is no milder clinical diagnosis in the patient’s history or the clinical diagnosis data is missing. Overall, this yields a 101-dimensional input vector for L2C-FNN (see Supplementary Table S7 for details).

The loss function is computed by comparing the predictions with the ground truth. Similar to MinimalRNN, cross-entropy loss was used for clinical diagnosis prediction, while mean absolute error (MAE) loss was employed for MMSE and ventricle volume prediction (based on the Gauss Rank values). Because MAE was based on Gauss Rank values, the three losses were of similar magnitude, and so the three losses were added together with equal weighting. Changing the relative weights of the three terms could potentially influence the model performance. However, this would increase the number of hyperparameters, so we did not experiment with different weights in this study.

Finally, stochastic gradient descent (SGD) with momentum (Qian, 1999; Sutskever et al., 2013) was chosen as the optimizer, with Optuna utilized to search for optimal hyperparameters. Table 9 shows all the hyperparameters considered and their corresponding search range. The ExponentialLR scheduler was employed to regulate learning rate behavior.

**Table 9.** Hyper-parameters and corresponding search ranges for L2C-FNN estimated from the validation sets using Optuna.

| Hyper-parameter | Range |
| --- | --- |
| Dropout rate | 0-0.5 |
| LeakyReLU slope | 0.01-0.1 |
| L2 weight regularization | $10^{-7}$-$10^{-4}$ |
| SGD momentum | 0-0.9 |
| Learning rate | $10^{-5}$-$10^{-1}$ |
| ExponentialLR gamma | 0.1-0.9 |
| Num of hidden layers | 2-5 |
| Size of hidden state | 128-512 |

### 2.9 Further analyses

We performed five additional analyses to study the effectiveness of all five models (MinimalRNN, AD-Map and the three FROG variants).

#### 2.9.1 Impact of the number of observed timepoints on cross-cohort prediction accuracy

For a disease progression model to be effective in early detection of AD-dementia risk, it should ideally perform well with a small number of input timepoints. We evaluated the performance of all four models on external test datasets using only 1, 2, 3, or 4 input timepoints. This contrasts with the main benchmarking analysis (Section 3.2), where half the total number of timepoints (for each participant) were used to predict the remaining timepoints.

For the OASIS dataset, test subjects with fewer than 4 input timepoints were discarded so that the same test subjects were evaluated across the four conditions (i.e., 1, 2, 3, or 4 input timepoints). In contrast, the maximum number of input timepoints for each subject is less than 4 in the AIBL and MACC datasets (2 for AIBL and 3 for MACC). Consequently, we discarded test subjects with fewer than 2 input timepoints for AIBL and fewer than 3 input timepoints for MACC. Because we excluded some test subjects, the results of this analysis are not directly comparable to those of the main benchmarking analysis (Section 3.2).

#### 2.9.2 Breakdown of cross-cohort prediction in yearly intervals

We extended our investigation of cross-cohort prediction performance by breaking down the prediction results into yearly intervals up to 6 years into the future. Each participant’s future timepoints were categorized into yearly intervals based on the duration between the last input timepoint and the target future timepoint for prediction. For instance, considering a participant with 10 timepoints, if the last input timepoint (5th timepoint) was at month 60 and the 6th timepoint was at month 70, the prediction at the 6th timepoint would be classified as 1 year into the future due to the 10-month duration.

We anticipated that all tested algorithms would experience a decline in performance as the prediction horizon extended further into the future. Nevertheless, a robust algorithm was expected to maintain relatively high performance in both early and later years in the prediction horizon.

#### 2.9.3 Subgroup analysis of cross-cohort prediction performance

We conducted an analysis on the external test datasets to evaluate model performance across different diagnostic groups. Participants in each test set were divided into three subgroups (CN, MCI and DEM) based on their last observed clinical diagnosis from the input timepoints. We emphasize that clinical diagnoses from the output (test) timepoints were not used to divide participants into subgroups. Trained models from ADNI were applied and evaluated for each subgroup. A robust algorithm was expected to maintain relatively high performance across various diagnostic groups, ensuring effectiveness in different stages of disease progression.

#### 2.9.4 Effects of missing modalities on cross-cohort prediction accuracy

We also evaluated model performance under real-world conditions where certain biomarker modalities may be unavailable. For each external test dataset, we selected a subset of participants who had at least one observed value for every feature across all input timepoints, which we refer to as the “full feature set”. Participants missing an entire modality across all input timepoints were excluded from this analysis. Because we excluded some test participants, the results of this analysis are not directly comparable to the main benchmarking results (Section 3.2).

We generated three modified versions of the full feature set by ablating one modality at a time: MRI features, cognitive features, or diagnostic features (Table 1). In each version, all features belonging to the ablated modality were set to missing (NaN), while the remaining features were left unaltered. Trained models from ADNI were then applied and evaluated under each scenario. While all algorithms were expected to experience performance decline, a robust algorithm would maintain reasonable performance even under the missing-modality scenarios.

#### 2.9.5 Feature importance analysis for L2C-FNN

Given that L2C-FNN performed the best, we analyzed the L2C-FNN results from the previous ablation analysis (Section 2.9.4). More specifically, we examined the performance decline of L2C-FNN under each modality ablation scenario, by comparing with its performance on the full feature set. A substantial drop in performance following the removal of a modality indicated greater reliance on that modality, highlighting its importance to the model.

### 2.10 Deep neural network implementation

MinimalRNN and L2C-FNN were implemented using PyTorch (Paszke et al., 2019) and computed on NVIDIA RTX 3090 GPUs with CUDA 11.0.

### 2.11 Performance evaluation and statistical tests

In the preceding sections, we utilized a 20-fold cross-validation procedure to train models (MinimalRNN. AD-Map and three FROG variants) in ADNI, and then applied the models to predict clinical diagnosis, MMSE score, and ventricle volume in ADNI test sets (within-dataset evaluation), as well as in the three external datasets (cross-dataset evaluation). This section provides a detailed description of evaluation metrics and statistical tests.

Diagnosis classification accuracy was evaluated using the multiclass area under the operating curve (mAUC; Hand & Till, 2001) following the TADPOLE challenge. The mAUC was computed as the average of three two-class AUC (DEM vs not DEM, MCI vs. not MCI, and CN vs not CN). For mAUC, higher values indicate better performance. The mAUC is a group-level metric whereby the predictions were first pooled over all test participants across their entire forecast horizon into a vector of length # total future timepoints, before calculating the mAUC, resulting one value per test set.

MMSE and ventricles prediction accuracy was evaluated using mean absolute error (MAE), following the TADPOLE challenge (Marinescu et al., 2021), as well as MinimalRNN (Nguyen et al., 2020), and AD-Map (Maheux et al., 2023) studies. Lower MAE indicates better performance. The MAEs were averaged across all forecast timepoints within each participant, resulting in a vector of length #test_participant values per test set. For the main within-ADNI (Section 3.1) and cross-dataset (Section 3.2) analyses, root mean square error (RMSE) was also reported in the supplementary results, confirming that both MAE and RMSE exhibit similar trends.

For within-cohort evaluation, because of the 20-fold cross-validation, there were 20 mAUC values, 20 MAE values for MMSE and 20 MAE values for ventricle volumes. Although the test sets do not overlap, the participants used for training do overlap across the test sets. Therefore, the prediction metrics were not independent across the 20 test sets. To account for the non-independence, we utilized the corrected resampled t-test (Nadeau & Bengio, 2003) to assess performance differences between algorithms. Separate tests were performed for mAUC, MMSE and ventricle volume.

For cross-cohort evaluation, the final performance was computed by averaging the performance metrics across 20 trained models of each algorithm. To assess performance differences between algorithms, since each participant in the external datasets were independent, we performed paired sample t-test (Cohen, 1988) for MMSE MAE and ventricle volume MAE, as well as a permutation test (Good, 2000) for group-level metrics (mAUC). Supplementary Figures S3 and S4 illustrate the t-test and permutation test respectively.

Multiple comparisons were corrected with a false discovery rate (FDR) of q < 0.05 (Benjamini & Hochberg, 1995) for both within and cross-cohort evaluations.

## 3 Results

### 3.1 FROG variants perform the best for within-cohort clinical diagnosis prediction

Figure 3 and Table 10 compare the prediction performance of MinimalRNN, AD-Map and three FROG variants (L2C-XGBw, L2C-XGBnw and L2C-FNN) for within-cohort (ADNI) clinical diagnosis, MMSE and ventricle volume prediction. All models exhibited similar performance for predicting MMSE and ventricle volume. However, for predicting future clinical diagnosis, the three FROG variants were better than MinimalRNN, which was in turn better than AD-Map.

**Figure 3.**
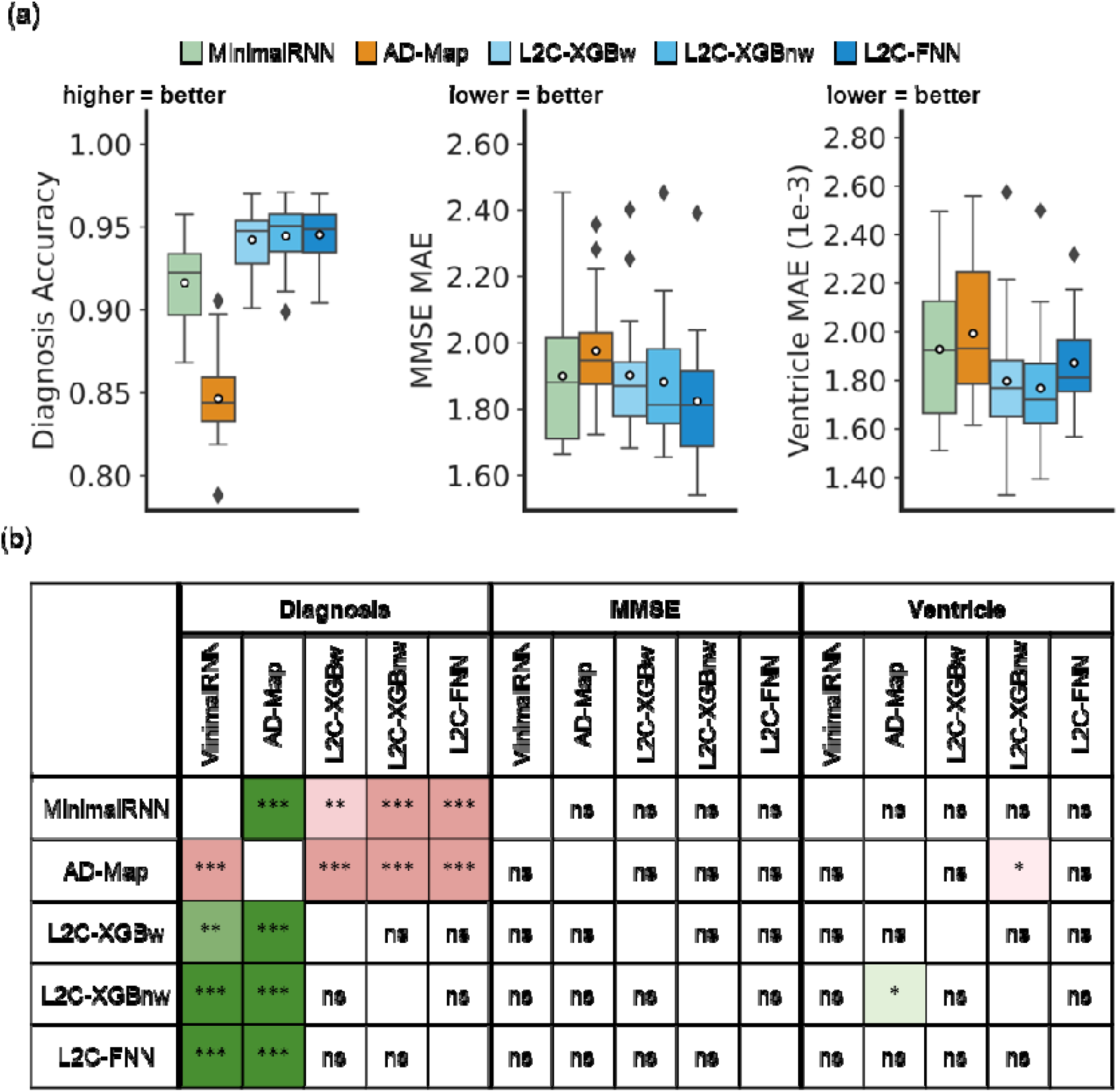
Within-cohort (ADNI) prediction performance. (a) Boxplots represent variability in prediction performance across 20 test folds for (left) clinical diagnosis prediction measured, (middle) MMSE prediction and (right) ventricular volume prediction. (b) Statistical difference between all models. “***” indicates p < 0.00001 and statistical significance after multiple comparison correction (FDR q < 0.05). “**” indicates p < 0.001 and statistical significance after multiple comparison correction (FDR q < 0.05). “ns” indicates no statistical significance (p ≥ 0.05) or did not survive FDR correction.

**Table 10.** Within-cohort (ADNI) prediction performance averaged across 20 folds. For clinical diagnosis, ↑ implies that higher mAUC indicates better performance. For MMSE and ventricular volume, ↓ indicates that lower MAE indicates better performance. The best result for each performance metric was bolded. Similar conclusions were obtained with root mean square error (RMSE) instead of MAE (Supplementary Table S8).

| | Diagnostic mAUC ↑ | MMSE MAE ↓ | Ventricle MAE (1e-3) ↓ |
| --- | --- | --- | --- |
| **MinimalRNN** | $0.916 \pm 0.027$ | $1.899 \pm 0.212$ | $1.928 \pm 0.279$ |
| **AD-Map** | $0.847 \pm 0.027$ | $1.976 \pm 0.171$ | $1.993 \pm 0.278$ |
| **L2C-XGBw** | $0.942 \pm 0.018$ | $1.896 \pm 0.185$ | $1.796 \pm 0.279$ |
| **L2C-XGBnw** | $0.945 \pm 0.019$ | $1.875 \pm 0.192$ | **$1.768 \pm 0.247$** |
| **L2C-FNN** | **$0.946 \pm 0.018$** | **$1.824 \pm 0.201$** | $1.884 \pm 0.193$ |

### 3.2 L2C-FNN performed the best for cross-cohort prediction

Figures 4 and 5 compare the prediction performance of MinimalRNN, AD-Map and three FROG variants (L2C-XGBw, L2C-XGBnw and L2C-FNN) for cross-cohort MMSE, ventricle volume and clinical diagnosis prediction in three external datasets (AIBL, MACC, and OASIS). Numerical values are reported in Table 11.

**Figure 4.**
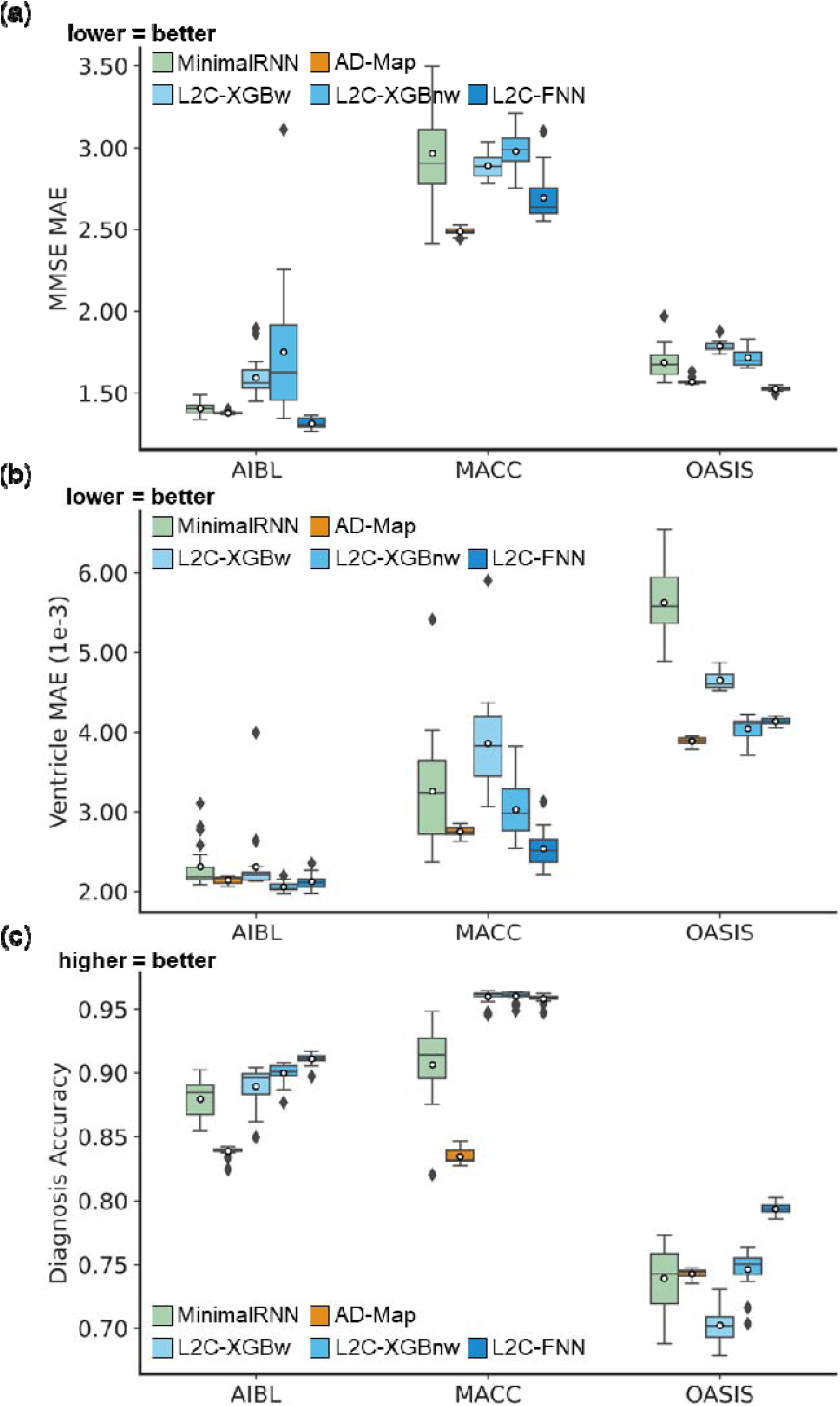
Cross-cohort prediction performance on three external test datasets. (a) Boxplots illustrate the variability across 20 trained models (from ADNI) for MMSE prediction assessed using MAE. Lower MAE indicates better performance. The x-axis denotes the test dataset used for evaluation. (b) Same as (a) but for ventricular volume prediction error assessed using MAE. Lower MAE indicates better performance. (c) Same as (a) but for diagnosis accuracy (mAUC). Higher mAUC indicates better performance.

**Figure 5.**
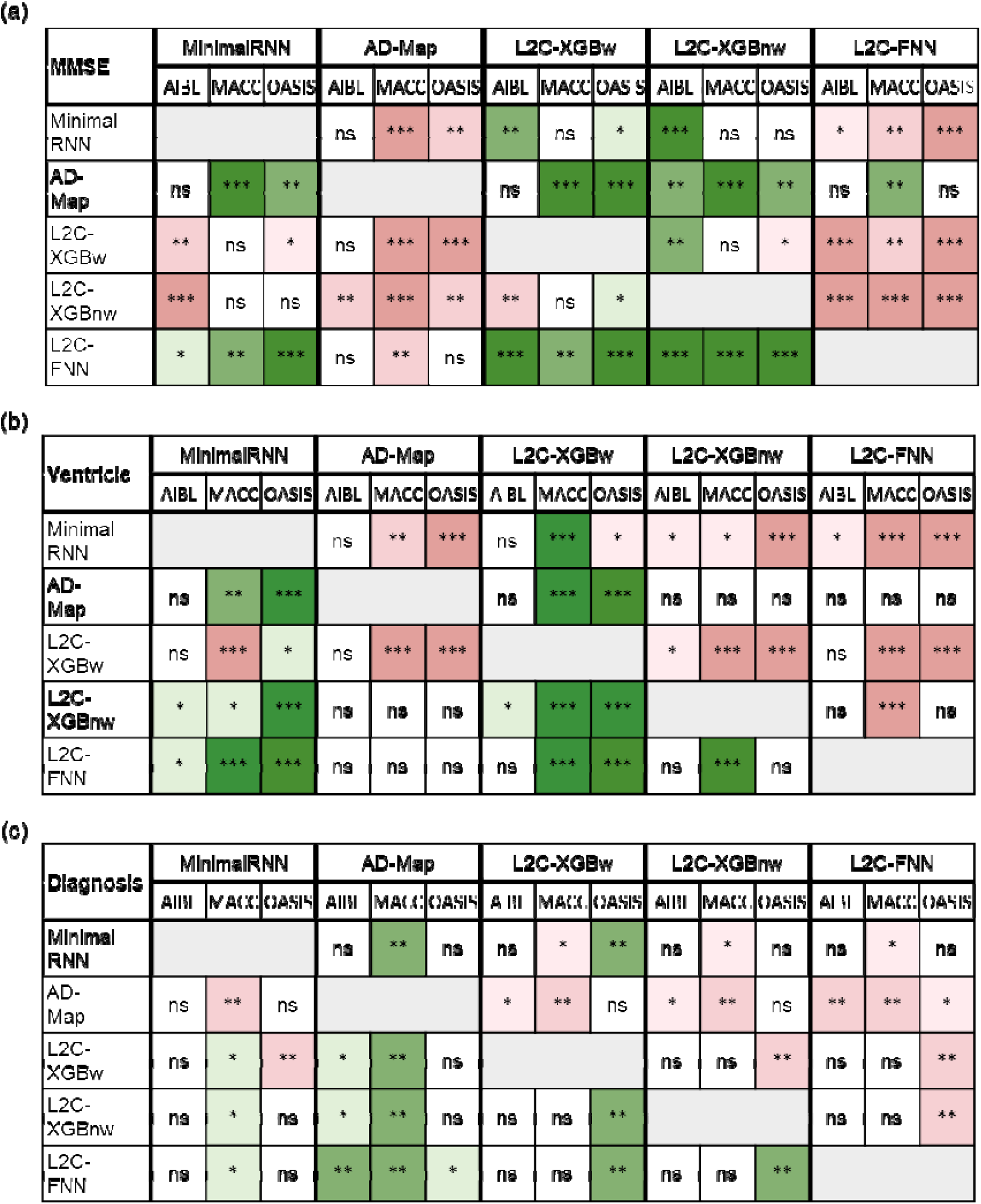
Statistical significance in the prediction performance between all models across the three external datasets. (a) Statistical significance between algorithms for MMSE prediction. Each row shows the statistical difference between a model and all other models across the three test datasets. The columns are grouped into five major columns corresponding to the five approaches. Within each major column, the three sub-columns represent results on AIBL, MACC, and OASIS datasets, respectively. For example, the first row corresponds to the statistical difference between MinimalRNN and the other models – green indicates that MinimalRNN performs better, while red indicates that MinimalRNN performs worse. “*” indicates p < 0.05 and statistical significance after multiple comparisons correction (FDR q < 0.05). “**” indicates p < 0.001 and statistical significance after multiple comparisons correction (FDR q < 0.05). “***” indicates p < 0.00001 and statistical significance after multiple comparisons correction (FDR q < 0.05). “ns” indicates no statistical significance (p ≥ 0.05) or did not survive FDR correction. (b) Same as (a) but for ventricular volume prediction. (c) Same as (a) but for clinical diagnosis prediction.

**Table 11.** Cross-cohort prediction performance averaged across 20 trained models (from ADNI). For clinical diagnosis, higher mAUC indicates better performance. For MMSE and ventricles, lower MAE indicates better performance. The best result for each performance metric on each test dataset was bolded. Similar conclusions were obtained with root mean square error (RMSE) instead of MAE (Supplementary Tables S9 and S10).

|  | MMSE MAE ↓ |  |  | Ventricle MAE (1e-3) ↓ |  |  | Diagnostic mAUC ↑ |  |  |
| --- | --- | --- | --- | --- | --- | --- | --- | --- | --- |
|  | AIBL | MACC | OASIS | AIBL | MACC | OASIS | AIBL | MACC | OASIS |
| MinimalRNN | 1.406 ± 0.040 | 2.965 ± 0.298 | 1.688 ± 0.095 | 2.311 ± 0.284 | 3.260 ± 0.709 | 5.625 ± 0.431 | 0.880 ± 0.016 | 0.906 ± 0.029 | 0.739 ± 0.024 |
| AD-Map | 1.379 ± 0.008 | **2.489 ± 0.022** | 1.569 ± 0.017 | 2.144 ± 0.044 | 2.749 ± 0.062 | **3.887 ± 0.045** | 0.839 ± 0.004 | 0.835 ± 0.006 | 0.743 ± 0.004 |
| L2C-XGBw | 1.595 ± 0.119 | 2.891 ± 0.075 | 1.787 ± 0.030 | 2.310 ± 0.412 | 3.858 ± 0.636 | 4.652 ± 0.118 | 0.889 ± 0.015 | **0.960 ± 0.005** | 0.702 ± 0.013 |
| L2C-XGBnw | 1.752 ± 0.415 | 2.979 ± 0.116 | 1.718 ± 0.057 | **2.057 ± 0.060** | 3.029 ± 0.350 | 4.044 ± 0.150 | 0.900 ± 0.008 | **0.960 ± 0.004** | 0.746 ± 0.015 |
| L2C-FNN | **1.314 ± 0.033** | 2.696 ± 0.142 | **1.526 ± 0.013** | 2.124 ± 0.090 | **2.541 ± 0.223** | 4.133 ± 0.039 | **0.911 ± 0.004** | 0.958 ± 0.003 | **0.794 ± 0.004** |

To simplify the reported statistical tests (Figure 5) into an easier-to-understand format, Table 12 shows the ranking of all models for the three prediction tasks (MMSE, ventricle, diagnosis) across the three external datasets (AIBL, MACC, OASIS). For example, in the case of MMSE prediction in the MACC dataset, L2C-FNN was statistically better than three algorithms and was statistically worse than one algorithm (Figure 5a), so we assigned L2C-FNN a score of 3 – 1 = 2. On the other hand, AD-Map was statistically better than four algorithms and statistically worse than no algorithm for MMSE prediction in the MACC dataset (Figure 5a), so we assigned AD-Map a score of 4 – 0 = 4. By comparing the scores of different algorithms, we concluded that for MMSE prediction in the MACC dataset, AD-Map was ranked first, L2C-FNN was ranked second, while MinimalRNN, L2C-XGBw and L2C-XGBnw were tied for third with a score of –2 (Table 12).

**Table 12.**
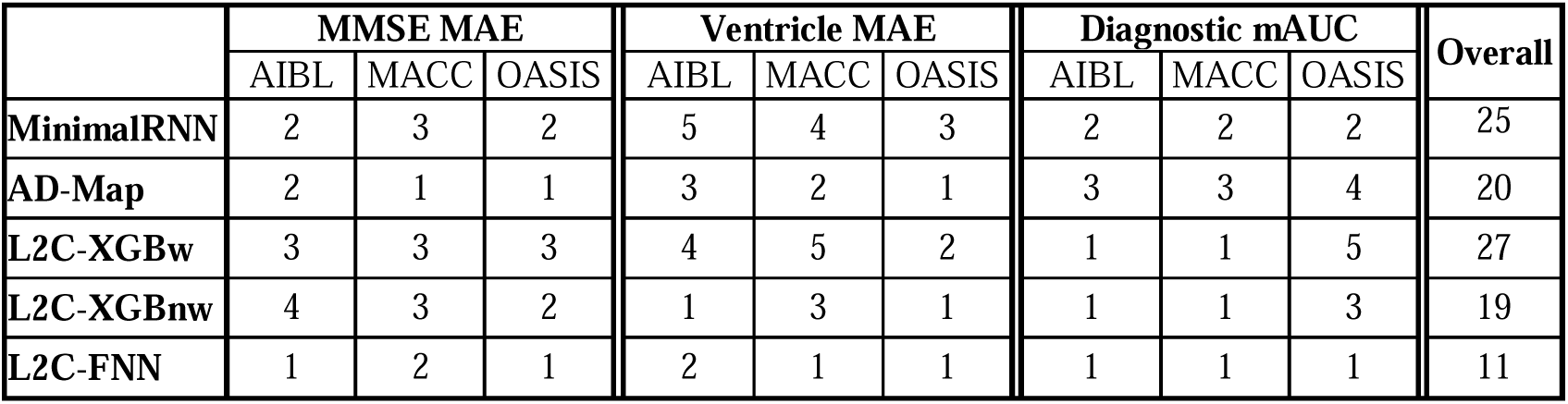
Cross-cohort prediction performance rankings of all models. Each row shows the rank of a model for three evaluation metrics: MMSE MAE, ventricle MAE, and diagnostic mAUC across AIBL, MACC, and OASIS datasets (1 = best, 5 = worst; ties allowed). Rankings were derived by summing wins (+1) and losses (–1) for each model, as indicated by green (win) and red (loss) in the statistical significance tables (Figure 5). The model with the highest total score receives rank 1 for that metric on each dataset. More details can be found in Section 3.2. The “overall” column adds up all the rankings, so a smaller overall ranking was better. The best possible overall ranking was nine, which could be achieved by being the top ranked algorithm for every prediction task in every dataset. With an overall ranking of 11, L2C-FNN performed the best.

Based on Table 12 (and Figures 4 and 5), in the case of MMSE prediction in the AIBL dataset, L2C-FNN was the best, followed by AD-Map and MinimalRNN. In the MACC dataset, AD-Map was the best followed by L2C-FNN. In the OASIS dataset, L2C-FNN and AD-Map were the best. Overall, for MMSE prediction, L2C-FNN and AD-Map were the best (Table 12). Similar conclusions were obtained if we only considered AD dementia, with non-AD dementia set to NaN (Figure S5).

In the case of ventricle volume prediction in the AIBL dataset, all approaches had similar performance, although L2C-XGBnw was the best, followed closely by L2C-FNN (Table 12). In the MACC dataset, L2C-FNN was the best, followed by AD-Map. In the OASIS dataset, AD-Map performed the best, followed by L2C-XGBnw. Overall, for ventricle volume prediction, L2C-FNN had the best ranking (Table 12). The original FROG algorithm (L2C-XGBw) and MinimalRNN performed the worst. Similar conclusions were obtained if we only considered AD dementia, with non-AD dementia set to NaN (Figure S5).

In the case of clinical diagnosis prediction in the AIBL dataset, L2C-FNN was numerically the best, but there was no statistical difference among the FROG variants and MinimalRNN. However, AD-Map was statistically worse than the three FROG variants. In the MACC dataset, the three FROG variants performed similarly well and were all statistically better than MinimalRNN, which was in turn better than AD-Map. Finally, in the OASIS dataset, L2C-FNN was the best, while the original FROG algorithm (L2C-XGBw) was the worst. Overall, for clinical diagnosis prediction, L2C-FNN performed the best. Similar conclusions were obtained if we only considered AD dementia, with non-AD dementia set to NaN (Figure S5).

The “overall” column in Table 12 shows the overall ranking by adding the rankings across all three prediction tasks (MMSE, ventricle, diagnosis) in the three external datasets. The best possible overall ranking was nine, which could be achieved by being the top ranked algorithm for every prediction task in every dataset. With an overall ranking of 11, L2C-FNN performed the best, followed by L2C-XGBnw (19) and AD-Map (20).

### 3.3 Further analysis 1: cross-dataset prediction with varying input timepoints

Figures S6 to S8 show the cross-dataset prediction performance of MinimalRNN, AD-Map and three FROG variants (L2C-XGBw, L2C-XGBnw and L2C-FNN) with varying number of input time points. Actual numerical values are reported in Tables S11 to S13. Due to the constraints of the datasets, the maximum number of input timepoints for each participant is only 2 for AIBL and 3 for MACC. Therefore, results for AIBL with 3 and 4 timepoints and for MACC with 4 timepoints are marked as “N.A.” Figure 6 shows the results of statistical tests comparing L2C-FNN and other approaches.

**Figure 6.**
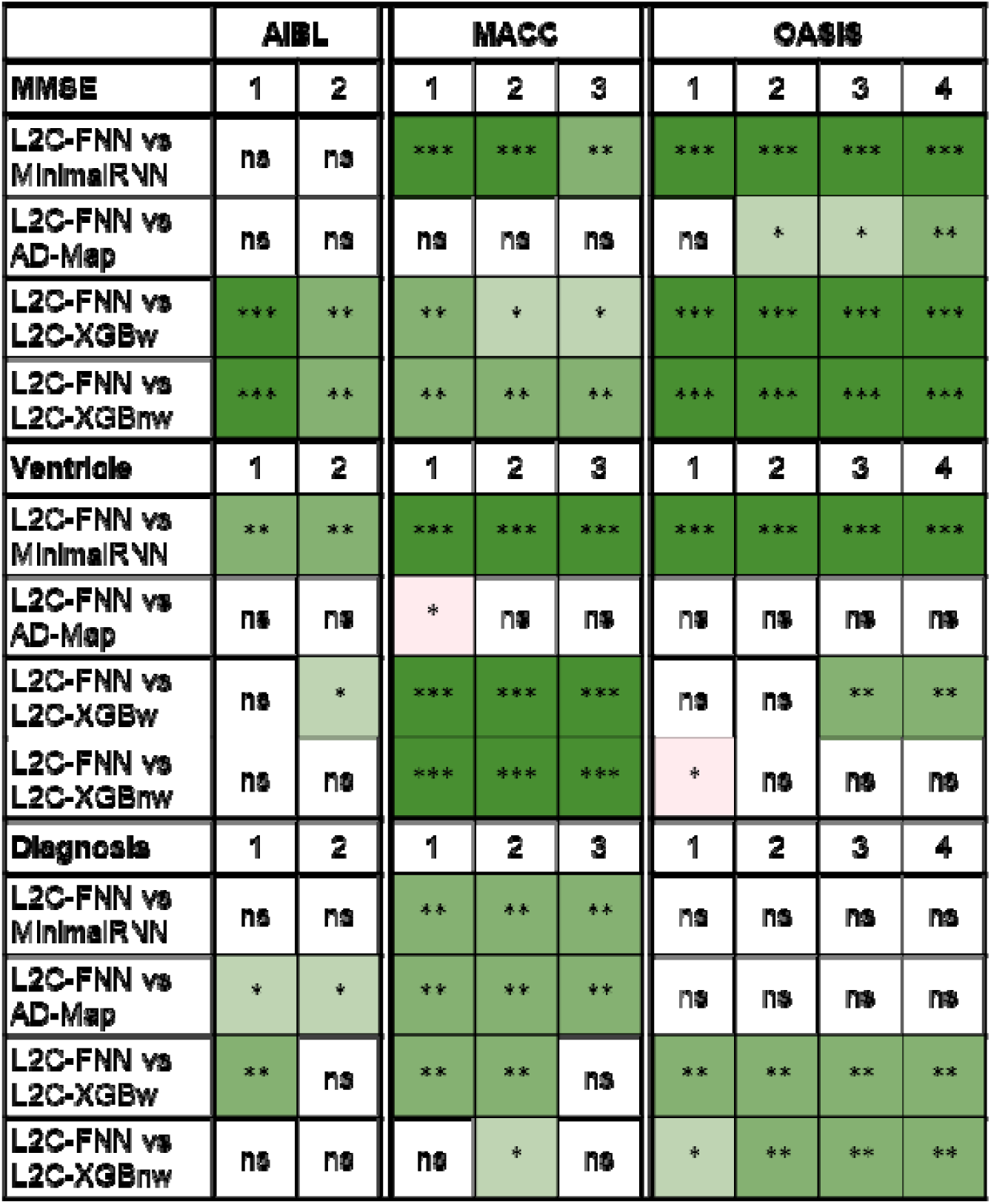
Statistical significance between L2C-FNN and other models for cross-cohort MMSE, ventricle volume, and clinical diagnosis prediction performance, using different numbers of input timepoints (after training with all timepoints in ADNI). “*” indicates p < 0.05 and significance after multiple comparisons correction (FDR q < 0.05). “**” indicates p < 0.001 and significance after multiple comparisons correction (FDR q < 0.05). “***” indicates p < 0.00001 and significance after multiple comparisons correction (FDR q < 0.05). “ns” indicates no significance (p ≥ 0.05) or did not survive FDR correction. Green indicates that L2C-FNN was statistically better than other approaches compared, while red indicates that it was statistically worse.

As anticipated, prediction performance generally improved with more input timepoints (Figures S6 to S8). Overall, L2C-FNN consistently matched or outperformed other approaches across all datasets and different number of observed timepoints, with only two exceptions (Figure 6). The first exception was that AD-Map was statistically better than L2C-FNN when predicting ventricle volume with 1 input timepoint in the MACC dataset (Figure 6). The second exception was that L2C-XGBnw was statistically better than L2C-FNN when predicting ventricle volume with 1 input timepoint in the OASIS dataset (Figure 6).

### 3.4 Further analysis 2: cross-dataset prediction across varying forecast windows

Supplementary Figures S9 to S11 show the yearly breakdown in prediction performance up to 6 years into the future (from Figure 4). As anticipated, the prediction performance for all algorithms declines as the prediction horizon increases. Numerical values are reported in Supplementary Tables S14 to S16. Figure 7 shows the results of statistical tests comparing L2C-FNN with other approaches. L2C-FNN consistently matched or outperformed other methods from year 1 to year 6 across all datasets with two exceptions (Figure 7). The exceptions were that AD-Map was statistically better than L2C-FNN when predicting MMSE in year 0-1 and year 1-2 in the MACC dataset.

**Figure 7.**
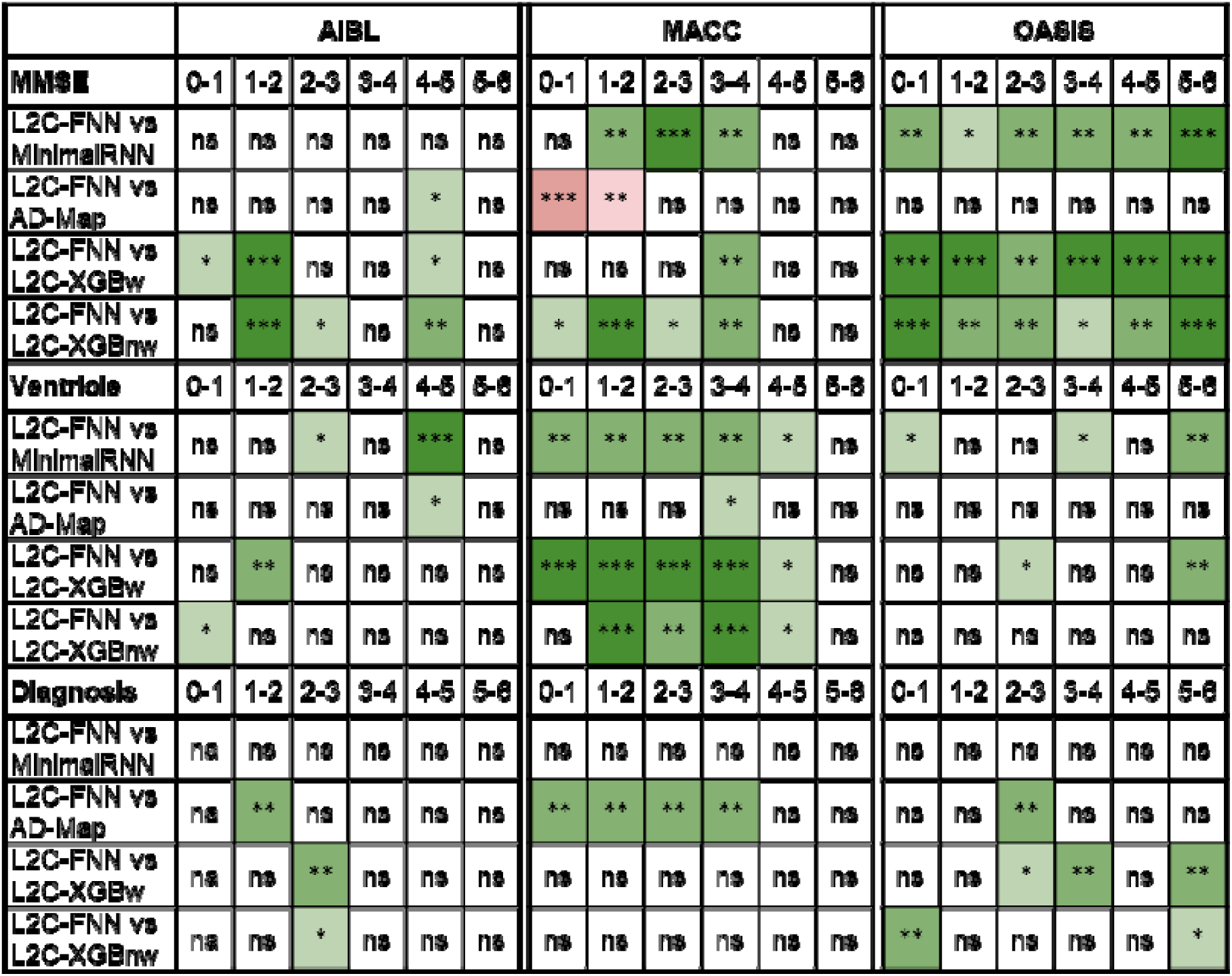
Statistical significance between L2C-FNN and other models for cross-cohort MMSE, ventricle volume, and clinical diagnosis prediction performance (from Figure 4) broken down into yearly intervals up to 6 years into the future. “*” indicates p < 0.05 and significance after multiple comparisons correction (FDR q < 0.05). “**” indicates p < 0.001 and significance after multiple comparisons correction (FDR q < 0.05). “***” indicates p < 0.00001 and significance after multiple comparisons correction (FDR q < 0.05). “ns” indicates no significance (p ≥ 0.05) or did not survive FDR correction. Green indicates L2C-FNN was statistically better than other approaches compared, while red indicated that it was statistically worse.

### 3.5 Further analysis 3: Initial diagnostic group

Tables S17 to S19 report the prediction performance (from Figure 4) broken down by last observed diagnostic group (CN, MCI or DEM). Figure 8 shows the results of statistical tests comparing L2C-FNN with other approaches. Overall, L2C-FNN demonstrated robust performance across clinical groups, showing comparable or better performance than other models (Figure 8). The one exception was that AD-Map was statistically better than L2C-FNN in predicting MMSE within the dementia group in the MACC dataset.

**Figure 8.**
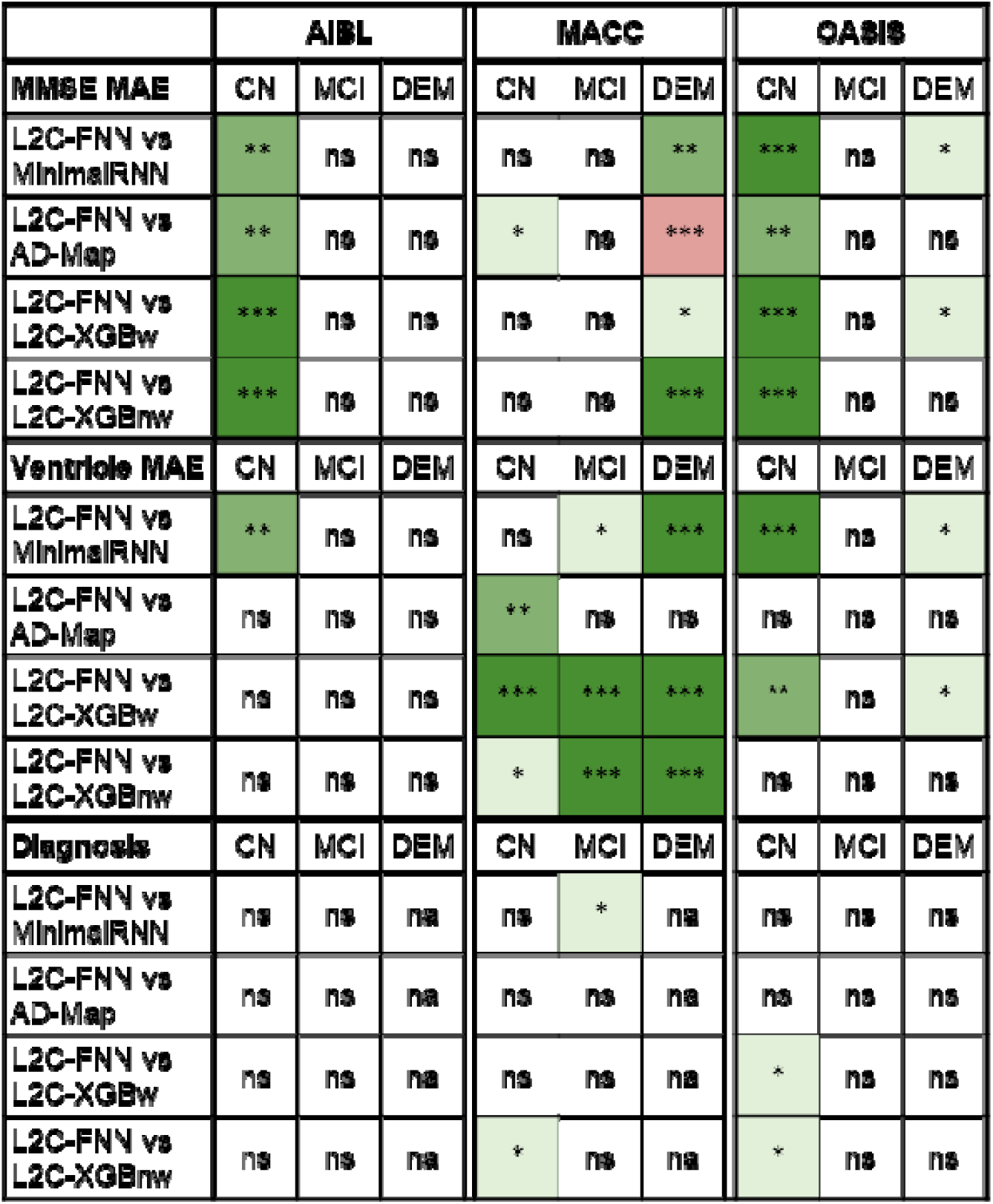
Statistical significance between L2C-FNN and other models for cross-cohort MMSE, ventricle volume, and clinical diagnosis prediction performance (from Figure 4) broken down by last observed diagnostic group (CN, MCI or DEM). “*” indicates p < 0.05 and significance after multiple comparisons correction (FDR q < 0.05). “**” indicates p < 0.001 and significance after multiple comparisons correction (FDR q < 0.05). “***” indicates p < 0.00001 and significance after multiple comparisons correction (FDR q < 0.05). “ns” indicates no significance (p ≥ 0.05) or did not survive FDR correction. Green indicates L2C-FNN was statistically better than other approaches compared, while red indicated that it was statistically worse. “na” indicates not applicable.

### 3.6 Further analysis 4: cross-dataset prediction with missing modalities

Supplementary Tables S20 to S22 report the prediction performance for different missing-modality scenarios. As expected, prediction accuracy declined for all algorithms when an entire data modality is missing in a participant. Figure 9 reports the statistical comparisons between L2C-FNN and other approaches. L2C-FNN consistently outperformed MinimalRNN and L2C-XGBw with the exception of statistically worse performance than MinimalRNN when predicting MMSE when cognitive features were ablated in the MACC dataset. Compared to AD-Map, L2C-FNN exhibited better performance in diagnosis and ventricular volume prediction. However, in the case of MMSE prediction, L2C-FNN was statistically better in two scenarios, and statistically worse in three scenarios, compared with AD-Map. Compared to L2C-XGBnw, L2C-FNN exhibited better performance in diagnosis and MMSE prediction. However, in the case of ventricular volume prediction, L2C-FNN was statistically better in five scenarios, and statistically worse in four scenarios (Figure 9). Therefore, overall, L2C-FNN compared favorably with other models.

**Figure 9.**
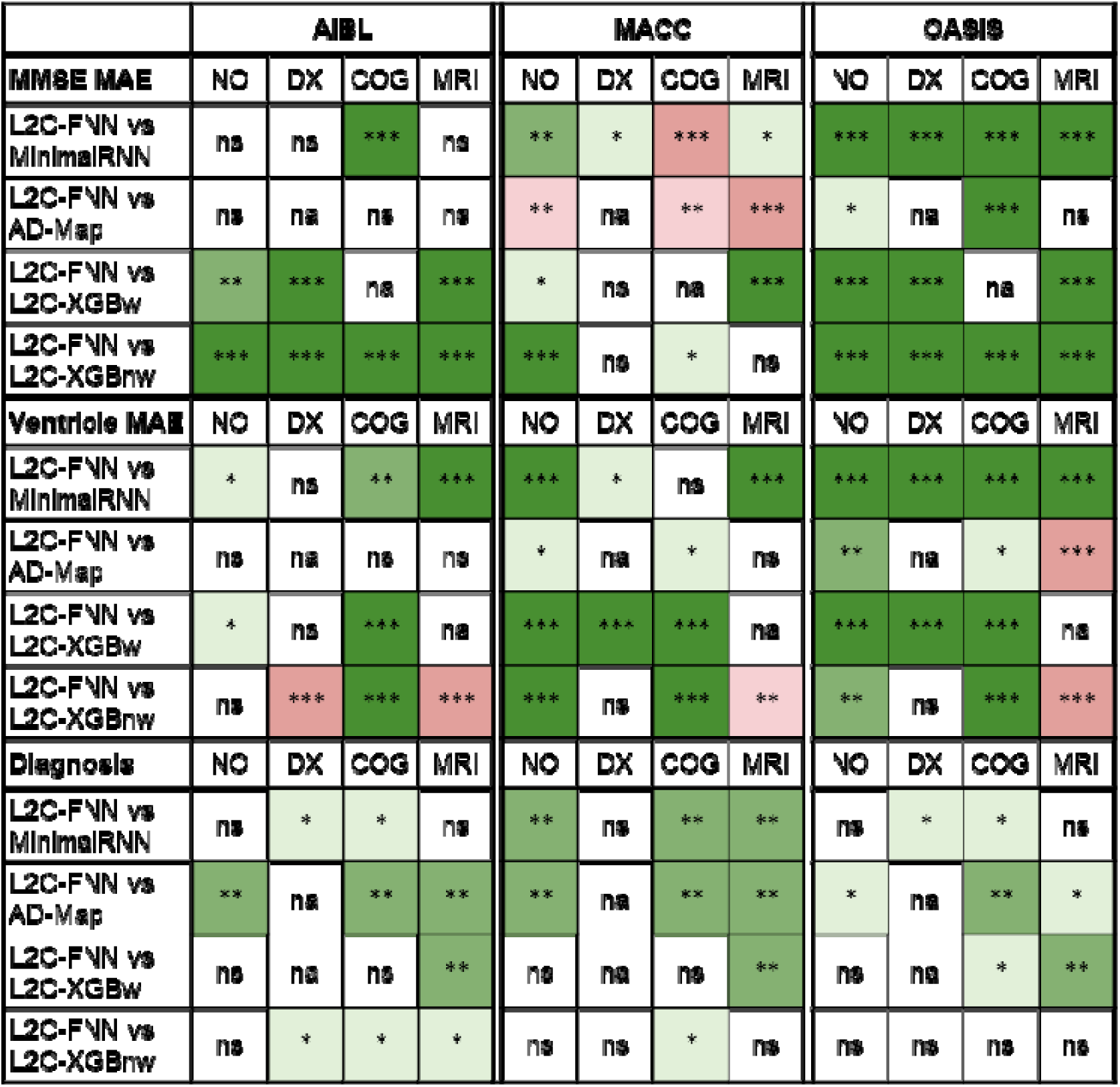
Statistical significance between L2C-FNN and other models for cross-cohort MMSE, ventricle volume, and clinical diagnosis prediction performance under different modality ablation scenarios (“NO”: no ablation; “DX”: ablate diagnosis; “COG”: ablate cognition; “MRI”: ablate MRI). “*” indicates p < 0.05 and significance after multiple comparisons correction (FDR q < 0.05). “**” indicates p < 0.001 and significance after multiple comparisons correction (FDR q < 0.05). “***” indicates p < 0.00001 and significance after multiple comparisons correction (FDR q < 0.05). “ns” indicates no significance (p ≥ 0.05) or did not survive FDR correction. Green indicates L2C-FNN was statistically better than other approaches compared, while red indicated that it was statistically worse. Due to model design, AD-Map did not utilize clinical diagnosis as input features (Section 2.5) and thus did not have results under the “DX” condition. These cases are marked as “na” (not applicable). On the other hand, L2C-XGBw relies on “time since most recent measurement” (MMSE, ventricle volume, or diagnosis) to select the appropriate XGBoost model (Section 2.6.4), so no prediction could be made for a modality if it is completely missing from a participant. Therefore, these cases are also marked as “na” (not applicable).

### 3.7 Further analysis 5: feature importance for L2C-FNN

Figure 10 quantifies feature importance for the L2C-FNN model by comparing prediction performance without ablation (dark color) and prediction performance with ablation (light color). Actual numerical results are reported in Supplementary Table S23.

**Figure 10.**
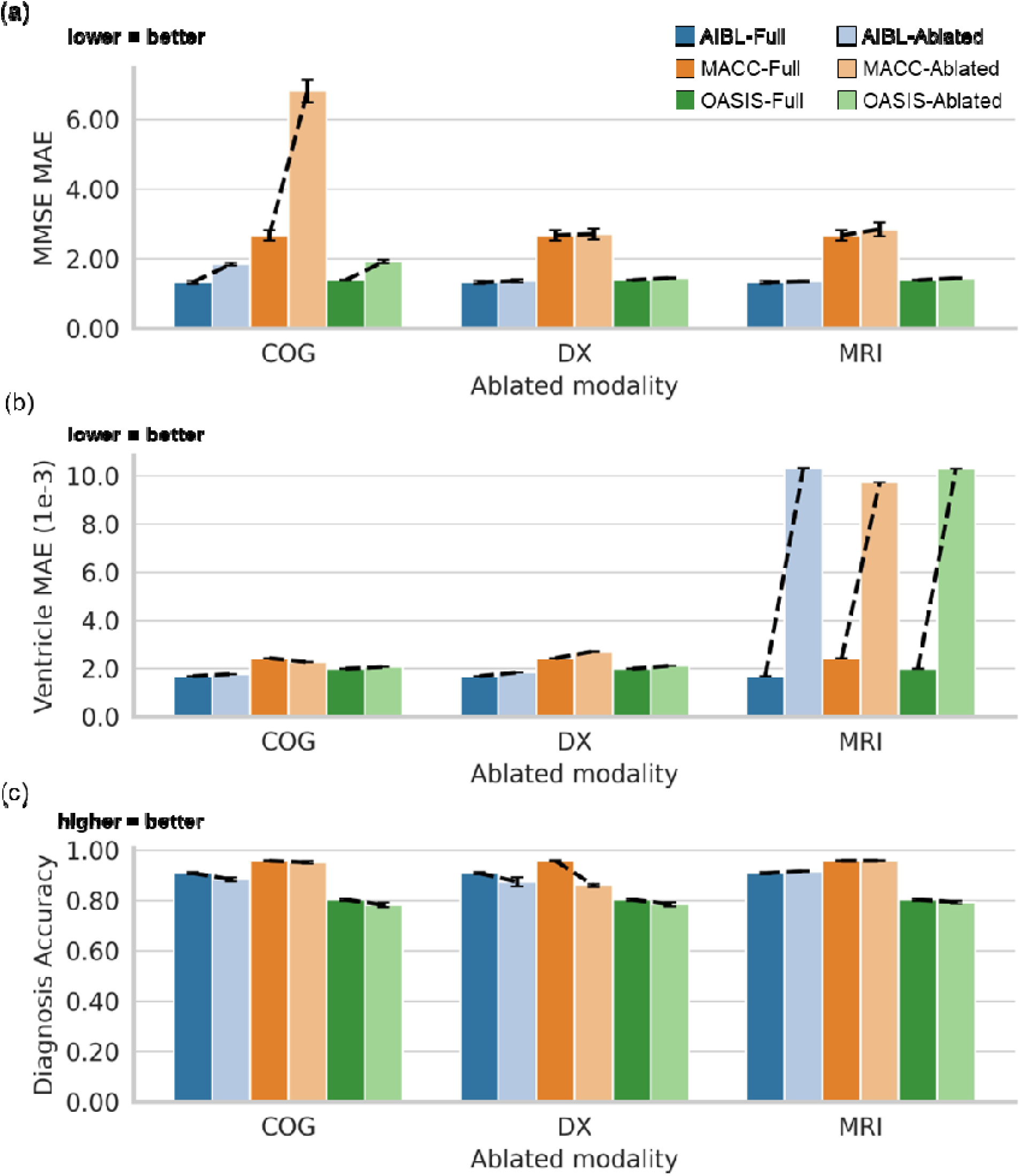
Cross-cohort feature importance analysis for L2C-FNN. (a) Bar charts show the average MMSE prediction error (MAE) across 20 trained models (from ADNI), with error bars indicating standard deviation. The x-axis denotes the ablated modality. For each ablated modality, six bars are shown: three datasets (AIBL, MACC, OASIS) each represented in two shades—dark for the full feature set (i.e., no ablation) and light for the ablated modality. Dotted lines connect each pair of bars within a dataset, highlighting the performance decline caused by modality ablation. (b) Same as (a) but for ventricle prediction error (MAE). (c) Same as (a) but for clinical diagnosis accuracy (mAUC).

When predicting MMSE, ablating MMSE from visit histories led to a massive increase in MMSE prediction error (Table S23; Figure 10a). This was especially pronounced in the MACC dataset, where MAE increased by 250% from 2.672 to 6.809 (Table S23). The second most important features were clinical diagnoses, followed by MRI, which impacted MMSE prediction accuracy only by 2% to 6% (Table S23).

When predicting ventricular volume, ablating MRI from visit histories led to an even larger increase in prediction error of around 400% to 600% (Table S23; Figure 10b). For example, MAE increased from 1.985 to 10.288 in the OASIS dataset (Table S23). The second most important features were clinical diagnoses, followed by cognitive features (Table S23).

Finally, excluding clinical diagnosis led to the largest drop in diagnosis prediction accuracy. However, accuracy degradation was quite modest (unlike MMSE and ventricle volume prediction), ranging from 3% to 10% (Table S23). Ablating cognitive features led to only 1% to 3% drop in prediction accuracy (Table S23). In the OASIS, ablating cognitive features led to a larger drop in accuracy than ablating clinical diagnoses. Finally, ablating MRI has little or no effect on clinical diagnosis prediction.

Overall, not surprisingly, cognitive features, MRI and clinical diagnoses were the most important for predicting MMSE, ventricle volume and clinical diagnoses respectively. What is interesting is that ablating the most important feature for clinical diagnosis prediction led to only a small drop in prediction performance unlike predicting MMSE or ventricular volume.

Furthermore, given the cost of MRI, if the goal is to predict future cognition or clinical diagnoses, our analyses suggested limited additional value of MRI beyond neuropsychological assessments. However, we note that we have only utilized T1 MRI in the current study. It is possible that other MRI modality might be more predictive of future cognitive decline or clinical diagnoses.

## 4 Discussion

In this study, we evaluated the winning algorithm of the TADPOLE challenge FROG, two FROG variants, MinimalRNN and AD-Map in the ADNI dataset, as well as three external datasets. In the ADNI dataset, all three FROG variants performed similarly well and outperformed MinimalRNN and AD-Map for clinical diagnosis. The excellent performance of FROG in clinical diagnosis was consistent with the outcome of the TADPOLE challenge, where FROG was ranked 1st in clinical diagnosis prediction (Marinescu et al., 2021).

In the three external datasets, the FROG variant L2C-FNN outperformed the other two FROG variants, MinimalRNN and AD-map, achieving top rank in seven of nine scenarios and second in the remaining two scenarios (Table 12). Furthermore, L2C-FNN mostly outperformed other models, regardless of the number of observed input timepoints (many more green entries than red entries in Figure 6), and regardless of the prediction horizon from 0 to 6 years into the future (many more green entries than red entries in Figure 7). L2C-FNN also mostly outperformed other models regardless of the last observed diagnoses (many more green entries than red entries in Figure 8), and regardless of missing input modality (many more green entries than red entries in Figure 9).

### 4.1 Algorithmic considerations

An inherent challenge in the TADPOLE problem set up is the pervasive missing data in each participant. Missing data occurs when participants fail to show up or fail to complete certain tests or scans during visits. In most longitudinal datasets, not all data is collected at all timepoints by design. For example, one visit might only involve MRI scans, while another visit might only involve detailed neuropsychological exams. Therefore, the implication is that in every participant, there is missing data at almost every observed time point.

State-based models, such as MinimalRNN, require specialized techniques to handle the missing data. By contrast, the L2C feature transformation significantly reduces the ratio of missing data. As a result, the input to models, comprising L2C features, contains substantially fewer missing data. Beyond mitigating missing data issues, this transformation also offers greater flexibility in model selection. By converting longitudinal data into a cross-sectional format, L2C removes the constraint of using time series models, allowing for a broader range of predictive algorithms. This flexibility enables the use of established machine learning models that may not naturally handle sequential inputs, potentially improving predictive performance and interpretability. These properties of L2C transformation might explain the advantage of FROG variants over MinimalRNN.

Another potential drawback of MinimalRNN is that each future prediction is based on previous predictions, which might lead to error accumulation (Fan et al., 2019). This error accumulation becomes particularly pronounced in longer-term predictions (Figures S9 to S11). Indeed, we observed that L2C-FNN’s improvements over MinimalRNN progressively improved further into the future (Figure S12a). This trend was not obvious when comparing L2C-FNN and other approaches (Figures S12b to S12d).

The original FROG algorithm (L2C-XGBw) constructs separate XGBoost models for each target variable and different forecast windows, resulting in a total of 15 models. However, the optimal window ranges might vary significantly between datasets. Furthermore, dividing training samples into different bins based on the forecast interval range reduces the available training samples in each bin. We hypothesized that eliminating forecast window stratification and enabling the model to implicitly leverage temporal information within L2C features (e.g., time since baseline, time since most recent measurements) could enhance model generalizability. To test this hypothesis, we trained a single FNN for all training samples with varying forecast intervals. Experimentally, L2C-FNN outperformed L2C-XGBw in every prediction task (i.e., clinical diagnosis, MMSE, ventricle volume) across all external datasets (Table 12).

Another feature of L2C-FNN is the use of multi-task learning, such that a single FNN is used to predict all three target variables (ventricle volume, cognition, clinical diagnosis) simultaneously. Multi-task learning leverages shared representations to capture common patterns among related tasks, which might enhance data efficiency, accelerate learning, and mitigate overfitting (Crawshaw, 2020). The FROG variant (L2C-XGBnw) helped to dissociate the effects of multi-task learning and the elimination of forecast window stratification by training a separate XGBoost model for each target variable. L2C-FNN again outperformed L2C-XGBnw in nearly every prediction task (i.e., clinical diagnosis, MMSE, ventricle volume) across all external datasets (Table 12). Therefore, these results suggest the potential advantage of multi-task learning.

In our evaluation, AD-Map was highly competitive in terms of predicting MMSE and ventricle volume but performed poorly for clinical diagnosis. Because AD-Map utilized a sigmoid-like parameterization, it might predict continuous variables (e.g., MMSE and ventricle volume) better than categorical variables (e.g., clinical diagnosis). Consistent with the original study (Maheux et al., 2023), we did not directly model clinical diagnosis in the AD-Map algorithm. We have also experimented with including clinical diagnosis in the AD-Map model, which improved clinical diagnosis prediction, but resulted in much worse MMSE and ventricle prediction (not shown).

### 4.2 Dataset variability and limitations

We observed significant variability in prediction performance across datasets (Figure 4) that was consistent across algorithms. For example, MMSE prediction error was the highest in the MACC dataset (compared with the AIBL or OASIS dataset) for all algorithms (Figure 4). The reason might be because the MMSE scores were significantly lower in the MACC dataset compared with the ADNI dataset, which was used to train the models. More specifically, the average baseline MMSE scores were 27.4, 28.0, 28.3 and 21.6 in ADNI, AIBL, OASIS and MACC datasets respectively (Table 2). The large shift in MMSE distributions between the ADNI and MACC datasets might have contributed to the relatively poor MMSE prediction in the MACC dataset.

On the other hand, ventricle volume and clinical diagnosis predictions were the worst in the OASIS dataset (compared with the MACC and AIBL datasets) for all algorithms (Figure 4). The reason for the poor ventricle volume prediction might be because MRI was missing in most OASIS timepoints. More specifically, 39%, 23%, 53% and 72% of timepoints did not have any MRI features in the ADNI, AIBL, MACC and OASIS datasets respectively (Table 2). The sparse MRI availability in OASIS required models to make predictions with few or even no imaging input features.

Furthermore, the average follow-up durations were 4.5, 3.3, 3.9 and 7.8 years in the ADNI, AIBL, MACC and OASIS datasets respectively (Table 2). The longer follow-up durations in OASIS required stronger long-term prediction capabilities, which may also explain the overall lower ventricle volume and diagnosis prediction performance in the OASIS dataset.

A limitation of the current study is that the models are trained from a single dataset (ADNI). We expect that models trained from multiple datasets might lead to better generalization to new populations and scanners (Dou et al., 2019; Liu et al., 2020; Chen et al., 2024). Therefore, a potential future work is to collate multiple datasets and train a single L2C-FNN model for future usage. Another limitation is that the current study only considered biomarkers that existed in all four datasets, so blood and PET biomarkers were excluded. Blood and PET biomarkers have been shown to be important markers of AD dementia (Nordberg et al., 2010; Mattsson et al., 2017; Chételat et al., 2020; Chong et al., 2021), therefore future studies could likely benefit from the incorporation of these additional biomarkers.

Another limitation of the current study is that there are in general more early follow-up data than late follow-up data. For example, a participant with a 5-year follow-up will almost certainly have an earlier follow-up (e.g., 1-year or 2-year follow-up) before the 5-year follow-up. On the other hand, participants with 1-year follow-up might never return for future follow-up. As a result, the main cross-cohort results (Figure 4) are naturally weighted towards earlier prediction horizons. We ameliorated this limitation by examining yearly breakdown in prediction performance up to 6 years into the future (Figure 7), but could not extend further into the future due to insufficient data.

We note that both short-term and long-term predictions can be clinically useful. On the one hand, there is growing consensus that interventions should be as early as possible before neuronal damage becomes irreversible (Dubois et al., 2016; Scheltens et al., 2016; Porsteinsson et al., 2021; Aisen et al., 2022). Therefore, the ability to forecast disease progression many years in advance can be highly beneficial. On the other hand, short-term predictions may be more actionable in certain cases, particularly when considering a patient’s age and comorbidities. For example, predicting that a 90-year-old patient is at high risk of developing dementia in ten years may have limited clinical utility, given the average life expectancy at that age is approximately another 3 to 5 years in the United States (U.S. Social Security Administration, 2020).

Moreover, prognosis can guide treatment preferences. A patient facing a poor short-term outlook might be more willing to accept an intensive or burdensome treatment, such as frequent infusions with serious side effects, for the chance of near-term benefit. Conversely, a patient with a favorable short-term prognosis but poor long-term outlook might prefer a simpler regimen with fewer side effects, prioritizing quality of life over aggressive intervention.

## 5 Conclusion

In this study, we evaluated three FROG variants, MinimalRNN and AD-Map in predicting future dementia progression in the ADNI dataset and three external datasets. We found that a FROG variant (L2C-FNN) performed the best in the three external datasets. L2C-FNN maintained better prediction performance regardless of the number of observed timepoints in a participant. L2C-FNN also consistently matched or outperformed other approaches from year 1 to year 6 across all external datasets, underscoring its potential for reliable long-term prediction in dementia progression.

## 6 Acknowledgment

We would like to thank Christina Rabe, and Paul Manser from Team FROG for sharing their code with us, which significantly facilitated the current study. This research is supported by the NUS Yong Loo Lin School of Medicine (NUHSRO/2020/124/TMR/LOA), the Singapore National Medical Research Council (NMRC) LCG (OFLCG19May-0035), NMRC CTG-IIT (CTGIIT23jan-0001), NMRC STaR (STaR20nov-0003), NMRC OF-IRG (OFIRG24jan-0006; OFIRG24jul-0049), Singapore Ministry of Health (MOH) Centre Grant (CG21APR1009), the Temasek Foundation (TF2223-IMH-01), and the United States National Institutes of Health (R01MH120080 & R01MH133334). Any opinions, findings and conclusions or recommendations expressed in this material are those of the authors and do not reflect the views of the funders.

Data collection and sharing for this project was funded by the Alzheimer’s Disease Neuroimaging Initiative (ADNI) (National Institutes of Health Grant U01 AG024904) and DOD ADNI (Department of Defense award number W81XWH-12-2-0012). ADNI is funded by the National Institute on Aging, the National Institute of Biomedical Imaging and Bioengineering, and through generous contributions from the following: AbbVie, Alzheimer’s Association; Alzheimer’s Drug Discovery Foundation; Araclon Biotech; BioClinica, Inc.; Biogen; Bristol-Myers Squibb Company; CereSpir, Inc.; Cogstate; Eisai Inc.; Elan Pharmaceuticals, Inc.; Eli Lilly and Company; EuroImmun; F. Hoffmann-La Roche Ltd and its affiliated company Genentech, Inc.; Fujirebio; GE Healthcare; IXICO Ltd.;Janssen Alzheimer Immunotherapy Research & Development, LLC.; Johnson & Johnson Pharmaceutical Research & Development LLC.; Lumosity; Lundbeck; Merck & Co., Inc.;Meso Scale Diagnostics, LLC.; NeuroRx Research; Neurotrack Technologies; Novartis Pharmaceuticals Corporation; Pfizer Inc.; Piramal Imaging; Servier; Takeda Pharmaceutical Company; and Transition Therapeutics. The Canadian Institutes of Health Research is providing funds to support ADNI clinical sites in Canada. Private sector contributions are facilitated by the Foundation for the National Institutes of Health (www.fnih.org). The grantee organization is the Northern California Institute for Research and Education, and the study is coordinated by the Alzheimer’s Therapeutic Research Institute at the University of Southern California. ADNI data are disseminated by the Laboratory for Neuro Imaging at the University of Southern California.

Data were provided in part by OASIS-3: Longitudinal Multimodal Neuroimaging: Principal Investigators: T. Benzinger, D. Marcus, J. Morris; NIH P30 AG066444, P50 AG00561, P30 NS09857781, P01 AG026276, P01 AG003991, R01 AG043434, UL1 TR000448, R01 EB009352. AV-45 doses were provided by Avid Radiopharmaceuticals, a wholly owned subsidiary of Eli Lilly.

## 7 Data availability statement

The ADNI and the AIBL datasets can be accessed via the Image & Data Archive (https://ida.loni.usc.edu/). The MACC dataset can be obtained via a data-transfer agreement with the MACC (http://www.macc.sg/). The OASIS dataset can be requested from (https://www.oasis-brains.org/).

## 8 Code availability statement

Code for all five models can be found here (https://github.com/ThomasYeoLab/CBIG/tree/master/stable_projects/predict_phenotypes/Zhang2025_L2CFNN). Two co-authors (L.A. and N.W.) reviewed the code before merging it into the GitHub repository to reduce the chance of coding errors.

## 9 Author contribution statement

**C.Z.**: Conceptualization; Data curation; Formal analysis; Investigation; Methodology; Project administration; Software; Visualization; Writing – original draft; Writing – review & editing. **L.A.**: Software; Validation; Visualization; Writing – review & editing. **N.W.**: Software; Validation; Visualization; Writing – review & editing. **K.N.**: Investigation; Software; Data curation; Visualization; Writing – review and editing. **C.O.**: Visualization; Writing – review and editing. **P.C.**: Visualization; Writing – review & editing. **C.C.**: Resource; Writing – review & editing. **J.H.Z.**: Resource; Writing – review & editing. **K.L.**: Methodology; Software; Writing – review & editing. **B.T.T.Y.**: Conceptualization; Formal analysis; Funding acquisition; Investigation; Methodology; Resource; Supervision; Visualization; Writing – original draft; Writing – review & editing.

## 10 Conflict of interest statement

Associate Editor is co-author – Dr. Juan Helen Zhou is a handling editor of *Human Brain Mapping* and a co-author of this article. To minimize bias, they were excluded from all editorial decision-making related to the acceptance of this article for publication.

## Footnotes

1 The TADPOLE challenge website: https://tadpole.grand-challenge.org/. Note that the website may not be actively updated. In addition, even though the goal is to predict indefinitely into the future, evaluation can only be performed on the finite number of timepoints available in a dataset.

## Notes

### Competing Interest Statement

The authors have declared no competing interest.

### Author Declarations

The Institutional Review Board (IRB) of the National University of Singapore gave ethical approval for this work.

### Summary of Updates

Revised manuscript following peer review. GitHub repository is now publicly available.

