## Supplemental materials for "Cross-dataset Evaluation of Dementia Longitudinal Progression Prediction Models"

### Supplementary Materials

**Table S1**. Scanner information for 9668 scans in ADNI dataset.

| Vendor | Scanner Model | Field Strength | Number of Scans |
| --- | --- | --- | --- |
| GE | Discovery MR750 | 3.0T | 886 |
|  | Discovery MR750w | 3.0T | 123 |
|  | Genesis Signa | 1.5T | 248 |
|  |  | 3.0T | 6 |
|  | Signa Excite | 1.5T | 892 |
|  |  | 3.0T | 6 |
|  | Signa HDx | 1.5T | 463 |
|  |  | 3.0T | 36 |
|  | Signa HDxt | 1.5T | 208 |
|  |  | 3.0T | 419 |
|  | Signa Premier | 3.0T | 33 |
|  | Signa UHP | 3.0T | 1 |
| Philips | Achieva dStream | 3.0T | 88 |
|  | Ingenia | 3.0T | 180 |
|  | Ingenia Elition X | 3.0T | 8 |
|  | Achieva | 1.5T | 73 |
|  |  | 3.0T | 541 |
|  | Gemini | 3.0T | 32 |
|  | Gyroscan Intera | 1.5T | 12 |
|  | Gyroscan NT | 1.5T | 2 |
|  | Ingenuity | 3.0T | 18 |
|  | Intera | 1.5T | 333 |
|  |  | 3.0T | 217 |
|  | Intera Achieva | 1.5T | 6 |
| SIEMENS | Allegra | 3.0T | 32 |
|  | Avanto | 1.5T | 391 |
|  | Biograph_mMR | 3.0T | 19 |
|  | Espree | 1.5T | 24 |
|  | Numaris4 | 1.5T | 2 |
|  | Prisma | 3.0T | 191 |
|  | Prisma_fit/Magnetom Prisma_fit | 3.0T | 466 |
|  | Skyra | 3.0T | 406 |
|  | Skyra_fit | 3.0T | 17 |
|  | Sonata | 1.5T | 379 |
|  | SonataVision | 1.5T | 31 |
|  | Symphony/SymphonyTim | 1.5T | 671 |
|  | Trio/TrioTim | 3.0T | 1487 |
|  | Verio | 3.0T | 718 |
|  | Skyra\|DicomCleaner | 3.0T | 3 |

**Table S2**. Scanner information for 940 scans in AIBL dataset.

| Vendor | Scanner Model | Field Strength | Number of Scans |
| --- | --- | --- | --- |
| SIEMENS | Avanto | 1.5T | 245 |
|  | Trio/TrioTim | 3.0T | 622 |
|  | Verio | 3.0T | 73 |

**Table S3**. Scanner information for 1453 scans in MACC dataset.

| Vendor | Scanner Model | Field Strength | Number of Scans |
| --- | --- | --- | --- |
| SIEMENS | Prisma | 3.0T | 85 |
|  | Trio/TrioTim | 3.0T | 1368 |

**Table S4**. Scanner information for 2519 scans in OASIS dataset.

| Vendor | Scanner Model | Field Strength | Number of Scans |
| --- | --- | --- | --- |
| SIEMENS | Avanto | 1.5T | 2 |
|  | Biograph_mMR | 3.0T | 812 |
|  | Magnetom Vida | 3.0T | 305 |
|  | Prisma_fit/Magnetom Prisma_fit | 3.0T | 1 |
|  | Sonata | 1.5T | 40 |
|  | Trio/TrioTim | 3.0T | 1359 |

**Table S5.** Our study used five regional brain volumes (Hippocampus, Fusiform, MidTemp, Ventricles, WholeBrain), computed by summing the volumes of various FreeSurfer regions of interest. Note that the 6^th^ anatomical volume (intracranial volume) was directly provided by FreeSurfer.

| **Regional volumetric feature** | **Brain regions used to compute regional volumes** |
| --- | --- |
| Hippocampus | Left-Hippocampus, Right-Hippocampus |
| Fusiform | lh_fusiform_volume, rh_fusiform_volume |
| MidTemp | lh_middletemporal_volume, rh_middletemporal_volume |
| Ventricles | Left-Inf-Lat-Vent, Left-Lateral-Ventricle,  Right-Inf-Lat-Vent, Right-Lateral-Ventricle |
| WholeBrain | WM-hypointensities, Left-Cerebellum-Cortex,  Left-Cerebellum-White-Matter, Left-Thalamus-Proper,  Left-Caudate, Left-Putamen,  Left-Pallidum, Left-Hippocampus,  Left-Amygdala, Left-Accumbens-area,  Left-VentralDC, Right-Cerebellum-Cortex,  Right-Cerebellum-White-Matter, Right-Thalamus-Proper, Right-Caudate, Right-Putamen,  Right-Pallidum, Right-Hippocampus,  Right-Amygdala, Right-Accumbens-area,  Right-VentralDC, lhCortexVol,  rhCortexVol, lhCerebralWhiteMatterVol, rhCerebralWhiteMatterVol |

**Table S6**. Complete set of L2C-XGBw and L2C-XGBnw features and their corresponding original features.

| **Original feature** | **L2C features** |
| --- | --- |
| Demographic and time-related features | APOE4, is_male, educ, marital_status, current_age, month_since_baseline, |
| Clinical diagnosis (categorical: CN, MCI, DEM) | mr_dx, time_since_mr_dx, best_dx, time_since_best_dx, worst_dx, time_since_worst_dx, milder, time_since_milder, |
| MMSE score (numeric, ordinal: 0-30) | mr_MMSE, time_since_mr_MMSE, mr_change_MMSE, low_MMSE, time_since_low_MMSE, high_MMSE, time_since_high_MMSE, |
| CDR_GLOBAL score (numeric, ordinal: 0, 0.5, 1, 2, 3) | mr_CDR, time_since_mr_CDR, mr_change_CDR, low_CDR, time_since_low_CDR, high_CDR, time_since_high_CDR, |
| Ventricle volume (numeric, continuous) | mr_Ventricles, time_since_mr_Ventricles, mr_change_Ventricles, low_Ventricles, time_since_low_Ventricles, high_Ventricles, time_since_high_Ventricles, |
| Fusiform volume (numeric, continuous) | mr_Fusiform, time_since_mr_Fusiform, mr_change_Fusiform, low_Fusiform, time_since_low_Fusiform, high_Fusiform, time_since_high_Fusiform, |
| WholeBrain volume (numeric, continuous) | mr_WholeBrain, time_since_mr_WholeBrain, mr_change_WholeBrain, low_WholeBrain, time_since_low_WholeBrain, high_WholeBrain, time_since_high_WholeBrain, |
| Hippocampus volume (numeric, continuous) | mr_Hippocampus, time_since_mr_Hippocampus, mr_change_Hippocampus, low_Hippocampus, time_since_low_Hippocampus, high_Hippocampus, time_since_high_Hippocampus, |
| Middle temporal volume (numeric, continuous) | mr_MidTemp, time_since_mr_MidTemp, mr_change_MidTemp, low_MidTemp, time_since_low_MidTemp, high_MidTemp, time_since_high_MidTemp, |
| Intracranial volume (numeric, continuous) | mr_ICV, time_since_mr_ICV, mr_change_ICV, low_ICV, time_since_low_ICV, high_ICV, time_since_high_ICV |

**Table S7.** L2C-FNN input vector dimensionality. For discrete features, the calculation is performed after one-hot encoding, including an additional class for missing data or unknown.

| **Category** | **L2C features** | **Feature dimension** |
| --- | --- | --- |
| Discrete features (dimensions are after one-hot encoding, including “unknown” class) | apoe | 4 |
|  | is_male | 3 |
|  | marital_status | 3 |
|  | mr_CDR | 6 |
|  | high_CDR | 6 |
|  | low_CDR | 6 |
|  | mr_dx | 4 |
|  | best_dx | 4 |
|  | worst_dx | 4 |
|  | milder | 3 |
| MRI features | mr_Ventricles, time_since_mr_Ventricles, mr_change_Ventricles, low_Ventricles, time_since_low_Ventricles, high_Ventricles, time_since_high_Ventricles, mr_Fusiform, time_since_mr_Fusiform, mr_change_Fusiform, low_Fusiform, time_since_low_Fusiform, high_Fusiform, time_since_high_Fusiform, mr_WholeBrain, time_since_mr_WholeBrain, mr_change_WholeBrain, low_WholeBrain, time_since_low_WholeBrain, high_WholeBrain, time_since_high_WholeBrain, mr_Hippocampus, time_since_mr_Hippocampus, mr_change_Hippocampus, low_Hippocampus, time_since_low_Hippocampus, high_Hippocampus, time_since_high_Hippocampus, mr_MidTemp, time_since_mr_MidTemp, mr_change_MidTemp, low_MidTemp, time_since_low_MidTemp, high_MidTemp, time_since_high_MidTemp, mr_ICV, time_since_mr_ICV, mr_change_ICV, low_ICV, time_since_low_ICV, high_ICV, time_since_high_ICV | 42 |
| Cognitive features | mr_MMSE, time_since_mr_MMSE, mr_change_MMSE, low_MMSE, time_since_low_MMSE, high_MMSE, time_since_high_MMSE, time_since_mr_CDR, time_since_low_CDR, time_since_high_CDR, | 10 |
| Diagnostic features | time_since_mr_dx, time_since_best_dx, time_since_worst_dx | 3 |
| Demographic and time-dependent features | baseline education level, current_age, month_since_baseline | 3 |

Table S8. Within-cohort (ADNI) prediction performance averaged across 20 test folds, comparing MAE and RMSE (root mean square error) for MMSE and ventricular volume predictions. Lower values indicate better performance. The best result for each metric was bolded. MAE and RMSE exhibit consistent trends across models, suggesting the robustness of the evaluation.

|  | MMSE MAE ↓ | MMSE RMSE ↓ | Ventricle MAE ↓ | Ventricle RMSE ↓ |
| --- | --- | --- | --- | --- |
| MinimalRNN | 1.899 ± 0.212 | 2.136 ± 0.227 | 1.928 ± 0.279 | 2.126 ± 0.297 |
| AD-Map | 1.976 ± 0.171 | 2.198 ± 0.183 | 1.993 ± 0.278 | 2.174 ± 0.310 |
| L2C-XGBw | 1.896 ± 0.185 | 2.127 ± 0.190 | 1.796 ± 0.279 | 1.993 ± 0.310 |
| L2C-XGBnw | 1.875 ± 0.192 | 2.087 ± 0.195 | **1.768 ± 0.247** | **1.948 ± 0.273** |
| L2C-FNN | **1.824 ± 0.201** | **2.083 ± 0.200** | 1.884 ± 0.193 | 2.066 ± 0.216 |

Table S9. Cross-cohort prediction performance averaged across 20 trained models (from ADNI), comparing MAE and RMSE for MMSE prediction. Lower values indicate better performance. The best result for each metric in each test dataset was bolded. MAE and RMSE exhibit consistent trends across models and datasets, ensuring the robustness of the evaluation.

|  | MMSE MAE ↓ | | | MMSE RMSE ↓ | | |
| --- | --- | --- | --- | --- | --- | --- |
|  | AIBL | MACC | OASIS | AIBL | MACC | OASIS |
| MinimalRNN | 1.406 ± 0.040 | 2.965 ± 0.298 | 1.688 ± 0.095 | 1.494 ± 0.041 | 3.250 ± 0.312 | 1.894 ± 0.101 |
| AD-Map | 1.379 ± 0.008 | **2.489 ± 0.022** | 1.569 ± 0.017 | 1.473 ± 0.010 | **2.744 ± 0.022** | **1.753 ± 0.018** |
| L2C-XGBw | 1.595 ± 0.119 | 2.891 ± 0.075 | 1.787 ± 0.030 | 1.681 ± 0.121 | 3.220 ± 0.086 | 2.007 ± 0.032 |
| L2C-XGBnw | 1.752 ± 0.415 | 2.979 ± 0.116 | 1.718 ± 0.057 | 1.827 ± 0.417 | 3.228 ± 0.114 | 1.908 ± 0.053 |
| L2C-FNN | **1.314 ± 0.033** | 2.696 ± 0.142 | **1.526 ± 0.013** | **1.415 ± 0.037** | 2.976 ± 0.133 | 1.764 ± 0.014 |

Table S10. Cross-cohort prediction performance averaged across 20 trained models (from ADNI), comparing MAE and RMSE for ventricular volume prediction. Lower values indicate better performance. The best result for each metric on each test dataset was bolded. MAE and RMSE exhibit consistent trends across models and datasets, ensuring the robustness of the evaluation.

|  | Ventricle MAE (1e-3) ↓ | | | Ventricle RMSE (1e-3) ↓ | | |
| --- | --- | --- | --- | --- | --- | --- |
|  | AIBL | MACC | OASIS | AIBL | MACC | OASIS |
| MinimalRNN | 2.311 ± 0.284 | 3.260 ± 0.709 | 5.625 ± 0.431 | 2.473 ± 0.289 | 3.314 ± 0.726 | 5.699 ± 0.438 |
| AD-Map | 2.144 ± 0.044 | 2.749 ± 0.062 | **3.887 ± 0.045** | 2.276 ± 0.045 | 2.788 ± 0.063 | **3.949 ± 0.046** |
| L2C-XGBw | 2.310 ± 0.412 | 3.858 ± 0.636 | 4.652 ± 0.118 | 2.465 ± 0.430 | 3.892 ± 0.636 | 4.727 ± 0.122 |
| L2C-XGBnw | **2.057 ± 0.060** | 3.029 ± 0.350 | 4.044 ± 0.150 | **2.200 ± 0.058** | 3.067 ± 0.350 | 4.120 ± 0.151 |
| L2C-FNN | 2.124 ± 0.090 | **2.541 ± 0.223** | 4.133 ± 0.039 | 2.269 ± 0.088 | **2.564 ± 0.223** | 4.200 ± 0.041 |

Table S11. Cross-cohort MMSE prediction performance using different numbers of input timepoints averaged across 20 trained models (training with all timepoints in ADNI). Lower MAE indicates better performance. The best result for each input horizon (i.e., number of input timepoints) on each test dataset was bolded. Due to dataset constraints, the maximum number of input timepoints for each participant is only 2 for AIBL and 3 for MACC. Therefore, results for AIBL with 3 and 4 timepoints and MACC with 4 timepoints are marked as “N.A.”

| MMSE MAE ↓ | 1 timepoints | 2 timepoints | 3 timepoints | 4 timepoints |
| --- | --- | --- | --- | --- |
| AIBL | | | | |
| MinimalRNN | 1.556 ± 0.057 | 1.380 ± 0.048 | N.A. | N.A. |
| AD-Map | 1.520 ± 0.007 | 1.442 ± 0.024 | N.A. | N.A. |
| L2C-XGBw | 1.828 ± 0.058 | 1.562 ± 0.074 | N.A. | N.A. |
| L2C-XGBnw | 1.961 ± 0.401 | 1.737 ± 0.382 | N.A. | N.A. |
| L2C-FNN | **1.463 ± 0.018** | **1.325 ± 0.030** | N.A. | N.A. |
| MACC | | | | |
| MinimalRNN | 4.046 ± 0.514 | 3.503 ± 0.447 | 2.777 ± 0.288 | N.A. |
| AD-Map | 3.072 ± 0.033 | **2.683 ± 0.029** | **2.340 ± 0.020** | N.A. |
| L2C-XGBw | 3.570 ± 0.169 | 3.066 ± 0.084 | 2.649 ± 0.054 | N.A. |
| L2C-XGBnw | 3.447 ± 0.208 | 3.081 ± 0.169 | 2.711 ± 0.112 | N.A. |
| L2C-FNN | **3.064 ± 0.205** | 2.750 ± 0.156 | 2.444 ± 0.137 | N.A. |
| OASIS | | | | |
| MinimalRNN | 1.835 ± 0.282 | 1.716 ± 0.221 | 1.623 ± 0.218 | 1.573 ± 0.204 |
| AD-Map | 1.399 ± 0.015 | 1.337 ± 0.018 | 1.338 ± 0.025 | 1.352 ± 0.023 |
| L2C-XGBw | 1.827 ± 0.163 | 1.697 ± 0.088 | 1.587 ± 0.076 | 1.511 ± 0.076 |
| L2C-XGBnw | 2.460 ± 0.153 | 2.159 ± 0.117 | 2.010 ± 0.106 | 1.916 ± 0.107 |
| L2C-FNN | **1.349 ± 0.055** | **1.234 ± 0.026** | **1.198 ± 0.023** | **1.174 ± 0.021** |

Table S12. Cross-cohort ventricular volume prediction performance using different numbers of input timepoints averaged across 20 trained models (training with all timepoints in ADNI). Lower MAE indicates better performance. The best result for each input horizon (i.e., number of input timepoints) on each test dataset was bolded. Due to dataset constraints, the maximum number of input timepoints for each participant is only 2 for AIBL and 3 for MACC. Therefore, results for AIBL with 3 and 4 timepoints and MACC with 4 timepoints are marked as “N.A.”

| Ventricle  MAE (1e-3) ↓ | 1 timepoints | 2 timepoints | 3 timepoints | 4 timepoints |
| --- | --- | --- | --- | --- |
| AIBL | | | | |
| MinimalRNN | 3.910 ± 0.349 | 2.064 ± 0.321 | N.A. | N.A. |
| AD-Map | 3.242 ± 0.039 | 2.122 ± 0.087 | N.A. | N.A. |
| L2C-XGBw | 3.437 ± 0.053 | 1.985 ± 0.294 | N.A. | N.A. |
| L2C-XGBnw | 3.216 ± 0.070 | 1.852 ± 0.063 | N.A. | N.A. |
| L2C-FNN | **3.168 ± 0.056** | **1.803 ± 0.076** | N.A. | N.A. |
| MACC | | | | |
| MinimalRNN | 4.766 ± 1.184 | 4.599 ± 1.206 | 2.836 ± 0.613 | N.A. |
| AD-Map | **2.673 ± 0.023** | **2.827 ± 0.054** | 2.616 ± 0.077 | N.A. |
| L2C-XGBw | 4.146 ± 0.391 | 3.938 ± 0.455 | 3.434 ± 0.557 | N.A. |
| L2C-XGBnw | 4.155 ± 0.269 | 3.746 ± 0.273 | 2.895 ± 0.299 | N.A. |
| L2C-FNN | 3.067 ± 0.183 | 2.921 ± 0.162 | **2.351 ± 0.194** | N.A. |
| OASIS | | | | |
| MinimalRNN | 12.112 ± 1.389 | 9.371 ± 1.082 | 7.321 ± 0.775 | 6.861 ± 0.684 |
| AD-Map | **7.489 ± 0.164** | 6.231 ± 0.124 | 5.309 ± 0.084 | **4.808 ± 0.077** |
| L2C-XGBw | 7.499 ± 0.022 | 6.491 ± 0.057 | 5.923 ± 0.090 | 5.532 ± 0.081 |
| L2C-XGBnw | 7.518 ± 0.302 | **6.117 ± 0.170** | 5.185 ± 0.098 | 4.812 ± 0.099 |
| L2C-FNN | 7.913 ± 0.139 | 6.130 ± 0.080 | **5.132 ± 0.064** | 4.870 ± 0.066 |

Table S13. Cross-cohort diagnosis prediction performance using different numbers of input timepoints averaged across 20 trained models (training with all timepoints in ADNI). Higher mAUC indicates better performance. The best result for each input horizon (i.e., number of input timepoints) on each test dataset was bolded. Due to dataset constraints, the maximum number of input timepoints for each participant is only 2 for AIBL and 3 for MACC. Therefore, results for AIBL with 3 and 4 timepoints and MACC with 4 timepoints are marked as “N.A.”

| mAUC ↑ | 1 timepoints | 2 timepoints | 3 timepoints | 4 timepoints |
| --- | --- | --- | --- | --- |
| AIBL | | | | |
| MinimalRNN | 0.806 ± 0.022 | 0.874 ± 0.019 | N.A. | N.A. |
| AD-Map | 0.719 ± 0.007 | 0.823 ± 0.008 | N.A. | N.A. |
| L2C-XGBw | 0.755 ± 0.039 | 0.890 ± 0.015 | N.A. | N.A. |
| L2C-XGBnw | 0.799 ± 0.021 | 0.887 ± 0.018 | N.A. | N.A. |
| L2C-FNN | **0.834 ± 0.011** | **0.903 ± 0.008** | N.A. | N.A. |
| MACC | | | | |
| MinimalRNN | 0.839 ± 0.029 | 0.852 ± 0.030 | 0.896 ± 0.033 | N.A. |
| AD-Map | 0.834 ± 0.005 | 0.829 ± 0.004 | 0.824 ± 0.008 | N.A. |
| L2C-XGBw | 0.866 ± 0.010 | 0.894 ± 0.008 | **0.960 ± 0.006** | N.A. |
| L2C-XGBnw | 0.885 ± 0.006 | 0.901 ± 0.006 | **0.960 ± 0.005** | N.A. |
| L2C-FNN | **0.895 ± 0.005** | **0.911 ± 0.004** | 0.958 ± 0.003 | N.A. |
| OASIS | | | | |
| MinimalRNN | 0.607 ± 0.027 | 0.627 ± 0.027 | 0.639 ± 0.028 | 0.656 ± 0.029 |
| AD-Map | 0.624 ± 0.004 | 0.640 ± 0.005 | 0.650 ± 0.004 | 0.686 ± 0.005 |
| L2C-XGBw | 0.529 ± 0.005 | 0.530 ± 0.010 | 0.541 ± 0.013 | 0.589 ± 0.011 |
| L2C-XGBnw | 0.589 ± 0.016 | 0.580 ± 0.019 | 0.598 ± 0.021 | 0.643 ± 0.020 |
| L2C-FNN | **0.657 ± 0.007** | **0.677 ± 0.009** | **0.691 ± 0.007** | **0.712 ± 0.007** |

Table S14. Cross-cohort MMSE prediction performance broken down into yearly intervals up to 6 years into the future. Results were averaged across 20 trained models (from ADNI). Lower MAE indicates better performance. The best result for each yearly interval (i.e., prediction horizon) on each test dataset was bolded.

| MMSE MAE ↓ | 0-1 | 1-2 | 2-3 | 3-4 | 4-5 | 5-6 |
| --- | --- | --- | --- | --- | --- | --- |
| AIBL | | | | | | |
| MinimalRNN | 0.771 ± 0.440 | 1.255 ± 0.034 | 1.517 ± 0.058 | 2.121 ± 0.115 | 1.456 ± 0.086 | 3.185 ± 0.279 |
| AD-Map | **0.136 ± 0.034** | 1.237 ± 0.009 | 1.492 ± 0.019 | **2.094 ± 0.035** | 1.677 ± 0.061 | 2.799 ± 0.053 |
| L2C-XGBw | 1.550 ± 0.433 | 1.497 ± 0.160 | 1.487 ± 0.090 | 2.316 ± 0.111 | 1.662 ± 0.092 | 2.721 ± 0.218 |
| L2C-XGBnw | 1.649 ± 0.771 | 1.678 ± 0.426 | 1.784 ± 0.348 | 2.191 ± 0.314 | 1.860 ± 0.417 | 3.113 ± 0.346 |
| L2C-FNN | 0.875 ± 0.222 | **1.209 ± 0.024** | **1.399 ± 0.072** | 2.112 ± 0.084 | **1.301 ± 0.036** | **2.483 ± 0.110** |
| MACC | | | | | | |
| MinimalRNN | 2.238 ± 0.158 | 2.869 ± 0.313 | 3.319 ± 0.431 | 3.942 ± 0.494 | 4.502 ± 0.670 | 4.609 ± 1.215 |
| AD-Map | **2.042 ± 0.015** | **2.343 ± 0.024** | **2.646 ± 0.031** | 3.307 ± 0.064 | **3.180 ± 0.123** | 2.506 ± 0.250 |
| L2C-XGBw | 2.479 ± 0.124 | 2.701 ± 0.090 | 2.920 ± 0.077 | 3.781 ± 0.092 | 4.057 ± 0.222 | 2.694 ± 0.643 |
| L2C-XGBnw | 2.570 ± 0.121 | 2.961 ± 0.147 | 2.997 ± 0.135 | 3.646 ± 0.191 | 3.649 ± 0.208 | 2.895 ± 0.684 |
| L2C-FNN | 2.371 ± 0.137 | 2.606 ± 0.164 | 2.724 ± 0.158 | **3.105 ± 0.151** | 3.550 ± 0.136 | **1.881 ± 0.443** |
| OASIS | | | | | | |
| MinimalRNN | 1.277 ± 0.050 | 1.440 ± 0.069 | 1.620 ± 0.101 | 1.662 ± 0.119 | 1.709 ± 0.166 | 1.942 ± 0.207 |
| AD-Map | 1.222 ± 0.015 | 1.365 ± 0.024 | 1.526 ± 0.015 | 1.521 ± 0.018 | 1.602 ± 0.047 | 1.647 ± 0.021 |
| L2C-XGBw | 1.350 ± 0.041 | 1.523 ± 0.038 | 1.671 ± 0.029 | 1.703 ± 0.041 | 1.824 ± 0.058 | 2.090 ± 0.083 |
| L2C-XGBnw | 1.322 ± 0.054 | 1.543 ± 0.063 | 1.671 ± 0.055 | 1.626 ± 0.062 | 1.785 ± 0.084 | 1.925 ± 0.067 |
| L2C-FNN | **1.171 ± 0.026** | **1.361 ± 0.015** | **1.501 ± 0.016** | **1.480 ± 0.017** | **1.502 ± 0.027** | **1.540 ± 0.027** |

Table S15. Cross-cohort ventricular volume prediction performance broken down into yearly intervals up to 6 years into the future. Results were averaged across 20 trained models (from ADNI). Lower MAE indicates better performance. The best result for each yearly interval (i.e., prediction horizon) on each test dataset was bolded.

| Ventricle  MAE (1e-3) ↓ | 0-1 | 1-2 | 2-3 | 3-4 | 4-5 | 5-6 |
| --- | --- | --- | --- | --- | --- | --- |
| AIBL | | | | | | |
| MinimalRNN | 0.978 ± 0.302 | 1.592 ± 0.221 | 2.895 ± 0.385 | 2.390 ± 0.364 | 3.454 ± 0.428 | 4.246 ± 1.093 |
| AD-Map | 0.958 ± 0.048 | 1.710 ± 0.036 | **2.427 ± 0.084** | 2.417 ± 0.037 | 3.428 ± 0.069 | 4.123 ± 0.138 |
| L2C-XGBw | 1.146 ± 0.170 | 1.907 ± 0.682 | 2.642 ± 0.147 | 2.118 ± 0.076 | 3.064 ± 0.069 | **3.199 ± 0.208** |
| L2C-XGBnw | 1.358 ± 0.134 | **1.576 ± 0.081** | 2.466 ± 0.115 | **2.075 ± 0.124** | 3.089 ± 0.082 | 3.245 ± 0.217 |
| L2C-FNN | **0.947 ± 0.266** | 1.604 ± 0.090 | 2.484 ± 0.103 | 2.147 ± 0.145 | **2.746 ± 0.091** | 3.349 ± 0.201 |
| MACC | | | | | | |
| MinimalRNN | 3.279 ± 0.631 | 2.403 ± 0.540 | 3.893 ± 0.862 | 4.778 ± 1.276 | 5.762 ± 2.062 | 6.377 ± 1.574 |
| AD-Map | **2.372 ± 0.037** | 2.150 ± 0.069 | 3.577 ± 0.063 | 4.391 ± 0.154 | 3.722 ± 0.162 | 5.038 ± 0.676 |
| L2C-XGBw | 4.209 ± 0.649 | 3.318 ± 0.612 | 4.117 ± 0.732 | 4.428 ± 0.683 | 4.372 ± 0.508 | 3.059 ± 0.401 |
| L2C-XGBnw | 2.812 ± 0.325 | 2.438 ± 0.394 | 3.575 ± 0.347 | 4.133 ± 0.313 | 4.548 ± 0.430 | 3.408 ± 0.621 |
| L2C-FNN | 2.598 ± 0.219 | **2.060 ± 0.247** | **3.007 ± 0.251** | **3.076 ± 0.247** | **2.894 ± 0.278** | **2.450 ± 0.387** |
| OASIS | | | | | | |
| MinimalRNN | 3.864 ± 0.155 | 4.132 ± 0.202 | 4.102 ± 0.296 | 5.769 ± 0.435 | 7.007 ± 0.848 | 7.312 ± 0.999 |
| AD-Map | **2.656 ± 0.041** | **3.451 ± 0.037** | 3.399 ± 0.058 | 4.082 ± 0.122 | **4.863 ± 0.071** | **4.630 ± 0.094** |
| L2C-XGBw | 3.291 ± 0.085 | 3.938 ± 0.069 | 3.884 ± 0.116 | 4.529 ± 0.172 | 5.723 ± 0.207 | 6.195 ± 0.369 |
| L2C-XGBnw | 2.774 ± 0.190 | 3.473 ± 0.074 | 3.410 ± 0.110 | **4.023 ± 0.159** | 5.048 ± 0.172 | 5.217 ± 0.322 |
| L2C-FNN | 3.102 ± 0.063 | 3.591 ± 0.065 | **3.360 ± 0.062** | 4.113 ± 0.082 | 4.926 ± 0.093 | 4.969 ± 0.079 |

Table S16. Cross-cohort clinical diagnosis prediction performance broken down into yearly intervals up to 6 years into the future. Results were averaged across 20 trained models (from ADNI). Higher mAUC indicates better performance. The best result for each yearly interval (i.e., prediction horizon) on each test dataset was bolded. Due to dataset constraints, AIBL only had one diagnostic class in year 0-1, making mAUC undefined in this case. Therefore, results for AIBL at year 0-1 is marked as “N.A.”

| mAUC ↑ | 0-1 | 1-2 | 2-3 | 3-4 | 4-5 | 5-6 |
| --- | --- | --- | --- | --- | --- | --- |
| AIBL | | | | | | |
| MinimalRNN | N.A. | 0.904 ± 0.014 | 0.903 ± 0.027 | **0.847 ± 0.025** | 0.810 ± 0.029 | 0.661 ± 0.105 |
| AD-Map | N.A. | 0.848 ± 0.007 | 0.891 ± 0.005 | 0.811 ± 0.005 | 0.763 ± 0.014 | 0.678 ± 0.044 |
| L2C-XGBw | N.A. | 0.926 ± 0.007 | 0.917 ± 0.019 | 0.791 ± 0.025 | 0.819 ± 0.014 | 0.868 ± 0.074 |
| L2C-XGBnw | N.A. | 0.918 ± 0.006 | 0.925 ± 0.010 | 0.823 ± 0.012 | 0.813 ± 0.016 | **0.880 ± 0.065** |
| L2C-FNN | N.A. | **0.928 ± 0.004** | **0.946 ± 0.006** | 0.839 ± 0.009 | **0.832 ± 0.012** | 0.854 ± 0.063 |
| MACC | | | | | | |
| MinimalRNN | 0.947 ± 0.025 | 0.906 ± 0.031 | 0.887 ± 0.028 | 0.860 ± 0.045 | 0.848 ± 0.034 | 0.831 ± 0.090 |
| AD-Map | 0.863 ± 0.003 | 0.842 ± 0.006 | 0.828 ± 0.007 | 0.779 ± 0.010 | 0.817 ± 0.024 | 0.676 ± 0.030 |
| L2C-XGBw | **0.986 ± 0.001** | **0.961 ± 0.004** | 0.930 ± 0.006 | 0.928 ± 0.009 | 0.869 ± 0.028 | 0.829 ± 0.067 |
| L2C-XGBnw | 0.984 ± 0.001 | 0.960 ± 0.004 | 0.932 ± 0.005 | **0.936 ± 0.007** | 0.889 ± 0.016 | 0.933 ± 0.021 |
| L2C-FNN | 0.983 ± 0.002 | 0.956 ± 0.004 | **0.937 ± 0.003** | **0.936 ± 0.004** | **0.909 ± 0.012** | **0.976 ± 0.019** |
| OASIS | | | | | | |
| MinimalRNN | 0.808 ± 0.052 | 0.827 ± 0.028 | 0.793 ± 0.030 | 0.747 ± 0.029 | 0.683 ± 0.041 | 0.649 ± 0.042 |
| AD-Map | 0.866 ± 0.006 | 0.785 ± 0.007 | 0.742 ± 0.004 | 0.749 ± 0.005 | **0.755 ± 0.011** | 0.686 ± 0.005 |
| L2C-XGBw | 0.787 ± 0.020 | 0.811 ± 0.012 | 0.743 ± 0.014 | 0.680 ± 0.027 | 0.676 ± 0.012 | 0.556 ± 0.014 |
| L2C-XGBnw | 0.836 ± 0.017 | 0.835 ± 0.018 | 0.815 ± 0.015 | 0.751 ± 0.026 | 0.694 ± 0.007 | 0.589 ± 0.018 |
| L2C-FNN | **0.887 ± 0.012** | **0.850 ± 0.007** | **0.856 ± 0.006** | **0.798 ± 0.008** | 0.749 ± 0.011 | **0.697 ± 0.012** |

Table S17. Cross-cohort MMSE prediction performance broken down by last observed diagnostic group (CN, MCI or DEM), averaged across 20 trained models (from ADNI). Lower MAE indicates better performance. The best result for each diagnostic group on each test dataset was bolded.

| MMSE MAE ↓ | CN | MCI | DEM |
| --- | --- | --- | --- |
| AIBL | | | |
| MinimalRNN | 0.993 ± 0.035 | 2.673 ± 0.101 | 3.379 ± 0.337 |
| AD-Map | 0.999 ± 0.007 | **2.583 ± 0.036** | **3.156 ± 0.056** |
| L2C-XGBw | 1.204 ± 0.147 | 2.821 ± 0.130 | 3.438 ± 0.275 |
| L2C-XGBnw | 1.396 ± 0.509 | 2.838 ± 0.274 | 3.458 ± 0.352 |
| L2C-FNN | **0.881 ± 0.011** | 2.607 ± 0.057 | 3.418 ± 0.304 |
| MACC | | | |
| MinimalRNN | 1.344 ± 0.053 | 2.321 ± 0.113 | 4.385 ± 0.661 |
| AD-Map | 1.458 ± 0.039 | **2.307 ± 0.032** | **3.200 ± 0.021** |
| L2C-XGBw | 1.370 ± 0.038 | 2.386 ± 0.054 | 4.138 ± 0.164 |
| L2C-XGBnw | 1.321 ± 0.058 | 2.383 ± 0.064 | 4.376 ± 0.280 |
| L2C-FNN | **1.301 ± 0.039** | **2.307 ± 0.094** | 3.777 ± 0.299 |
| OASIS | | | |
| MinimalRNN | 1.118 ± 0.053 | 1.503 ± 0.301 | 3.604 ± 0.283 |
| AD-Map | 1.065 ± 0.020 | **1.469 ± 0.024** | **3.316 ± 0.017** |
| L2C-XGBw | 1.225 ± 0.033 | 1.594 ± 0.064 | 3.675 ± 0.069 |
| L2C-XGBnw | 1.152 ± 0.048 | 1.591 ± 0.138 | 3.526 ± 0.117 |
| L2C-FNN | **0.987 ± 0.011** | 1.483 ± 0.086 | 3.367 ± 0.047 |

Table S18. Cross-cohort ventricular volume prediction performance broken down by last observed diagnostic group (CN, MCI or DEM), averaged across 20 trained models (from ADNI). Lower MAE indicates better performance. The best result for each diagnostic group on each test dataset was bolded.

| Ventricle  MAE (1e-3) ↓ | CN | MCI | DEM |
| --- | --- | --- | --- |
| AIBL | | | |
| MinimalRNN | 1.961 ± 0.352 | 3.617 ± 0.213 | **3.420 ± 0.364** |
| AD-Map | 1.665 ± 0.062 | 3.731 ± 0.074 | 3.887 ± 0.127 |
| L2C-XGBw | 1.763 ± 0.436 | 3.808 ± 0.396 | 4.659 ± 0.312 |
| L2C-XGBnw | **1.650 ± 0.065** | **3.497 ± 0.140** | 3.435 ± 0.207 |
| L2C-FNN | 1.701 ± 0.081 | 3.519 ± 0.178 | 3.671 ± 0.317 |
| MACC | | | |
| MinimalRNN | **1.321 ± 0.214** | 2.596 ± 0.553 | 5.218 ± 1.432 |
| AD-Map | 1.694 ± 0.061 | 2.530 ± 0.070 | 3.684 ± 0.072 |
| L2C-XGBw | 1.781 ± 0.255 | 2.946 ± 0.486 | 6.141 ± 1.203 |
| L2C-XGBnw | 1.568 ± 0.075 | 2.818 ± 0.223 | 4.237 ± 0.700 |
| L2C-FNN | 1.327 ± 0.081 | **2.303 ± 0.134** | **3.602 ± 0.487** |
| OASIS | | | |
| MinimalRNN | 4.002 ± 0.326 | 3.121 ± 0.540 | 8.480 ± 0.668 |
| AD-Map | **2.921 ± 0.055** | 3.732 ± 0.160 | 7.408 ± 0.104 |
| L2C-XGBw | 3.397 ± 0.148 | **1.873 ± 0.235** | 7.921 ± 0.091 |
| L2C-XGBnw | 3.031 ± 0.140 | 2.436 ± 0.626 | 6.834 ± 0.229 |
| L2C-FNN | 3.068 ± 0.041 | 2.103 ± 0.265 | **6.759 ± 0.118** |

Table S19. Cross-cohort diagnosis prediction performance broken down by last observed diagnostic group (CN, MCI or DEM), averaged across 20 trained models (from ADNI). Higher mAUC indicates better performance. The best result for each diagnostic group on each test dataset was bolded. “N.A.” indicates that mAUC could not be computed.

| mAUC ↑ | CN | MCI | DEM |
| --- | --- | --- | --- |
| AIBL | | | |
| MinimalRNN | 0.620 ± 0.063 | 0.609 ± 0.053 | N.A. |
| AD-Map | 0.521 ± 0.026 | 0.697 ± 0.026 | N.A. |
| L2C-XGBw | 0.718 ± 0.028 | **0.722 ± 0.037** | N.A. |
| L2C-XGBnw | 0.714 ± 0.022 | 0.694 ± 0.042 | N.A. |
| L2C-FNN | **0.722 ± 0.008** | 0.714 ± 0.017 | N.A. |
| MACC | | | |
| MinimalRNN | 0.708 ± 0.055 | 0.733 ± 0.030 | N.A. |
| AD-Map | **0.818 ± 0.024** | 0.739 ± 0.004 | N.A. |
| L2C-XGBw | 0.727 ± 0.018 | **0.795 ± 0.010** | N.A. |
| L2C-XGBnw | 0.724 ± 0.015 | 0.785 ± 0.009 | N.A. |
| L2C-FNN | 0.767 ± 0.016 | 0.788 ± 0.007 | N.A. |
| OASIS | | | |
| MinimalRNN | 0.607 ± 0.028 | 0.566 ± 0.078 | 0.704 ± 0.062 |
| AD-Map | 0.641 ± 0.009 | 0.542 ± 0.019 | **0.724 ± 0.007** |
| L2C-XGBw | 0.653 ± 0.014 | 0.580 ± 0.024 | 0.700 ± 0.013 |
| L2C-XGBnw | 0.653 ± 0.013 | **0.600 ± 0.022** | 0.711 ± 0.010 |
| L2C-FNN | **0.682 ± 0.006** | 0.590 ± 0.022 | 0.723 ± 0.016 |

Table S20. Cross-cohort MMSE prediction performance under different modality ablation scenarios. Results were averaged across 20 trained models (obtained from training with all data in ADNI). Lower MAE indicates better performance. The best result for each ablation scenario on each test dataset was bolded. Due to model design, AD-Map does not utilize diagnostic input features and therefore has no results in the “Ablate diagnosis” condition. On the other hand, L2C-XGBw relies on “time since most recent MMSE measurement” to select the appropriate XGBoost model, and this information is unavailable when cognitive features are ablated. Therefore, these cases are marked as “N.A.”

| MMSE MAE ↓ | No ablation | Ablate diagnosis | Ablate cognition | Ablate MRI |
| --- | --- | --- | --- | --- |
| AIBL | | | | |
| MinimalRNN | 1.368 ± 0.037 | 1.446 ± 0.067 | 2.408 ± 0.260 | 1.381 ± 0.043 |
| AD-Map | 1.384 ± 0.012 | N.A. | 2.178 ± 0.060 | **1.331 ± 0.009** |
| L2C-XGBw | 1.531 ± 0.115 | 1.569 ± 0.099 | N.A. | 2.031 ± 0.357 |
| L2C-XGBnw | 1.708 ± 0.408 | 2.187 ± 0.495 | 8.225 ± 1.750 | 1.835 ± 0.561 |
| L2C-FNN | **1.330 ± 0.040** | **1.369 ± 0.048** | **1.849 ± 0.041** | 1.354 ± 0.031 |
| MACC | | | | |
| MinimalRNN | 2.954 ± 0.303 | 2.854 ± 0.255 | **6.166 ± 0.510** | 3.074 ± 0.336 |
| AD-Map | **2.464 ± 0.020** | N.A. | 6.269 ± 0.631 | **2.480 ± 0.025** |
| L2C-XGBw | 2.831 ± 0.076 | 2.777 ± 0.068 | N.A. | 3.273 ± 0.091 |
| L2C-XGBnw | 2.965 ± 0.122 | 2.708 ± 0.083 | 7.638 ± 0.668 | 2.912 ± 0.183 |
| L2C-FNN | 2.672 ± 0.150 | **2.707 ± 0.155** | 6.809 ± 0.317 | 2.843 ± 0.203 |
| OASIS | | | | |
| MinimalRNN | 1.534 ± 0.070 | 1.561 ± 0.078 | 2.713 ± 0.338 | 1.574 ± 0.143 |
| AD-Map | 1.445 ± 0.019 | N.A. | 2.543 ± 0.169 | 1.450 ± 0.006 |
| L2C-XGBw | 1.654 ± 0.032 | 1.690 ± 0.044 | N.A. | 1.753 ± 0.078 |
| L2C-XGBnw | 1.568 ± 0.062 | 1.589 ± 0.048 | 8.205 ± 1.468 | 1.582 ± 0.077 |
| L2C-FNN | **1.387 ± 0.016** | **1.448 ± 0.018** | **1.922 ± 0.053** | **1.442 ± 0.022** |

Table S21. Cross-cohort ventricular volume prediction performance under different modality ablation scenarios. Results were averaged across 20 trained models (obtained from training with all data in ADNI). Lower MAE indicates better performance. The best result for each ablation scenario on each test dataset was bolded. Due to model design, AD-Map does not utilize diagnostic input features and therefore has no results in the “Ablate diagnosis” condition. On the other hand, L2C-XGBw relies on “time since most recent ventricle measurement” to select the appropriate XGBoost model, and this information is unavailable when MRI features are ablated. Therefore, these cases are marked as “N.A.”

| Ventricle  MAE (1e-3) ↓ | No ablation | Ablate diagnosis | Ablate cognition | Ablate MRI |
| --- | --- | --- | --- | --- |
| AIBL | | | | |
| MinimalRNN | 1.822 ± 0.304 | 1.913 ± 0.365 | 1.980 ± 0.403 | 12.772 ± 0.627 |
| AD-Map | 1.782 ± 0.049 | N.A. | 1.883 ± 0.053 | **8.922 ± 0.080** |
| L2C-XGBw | 1.823 ± 0.432 | 1.815 ± 0.445 | 4.803 ± 1.463 | N.A. |
| L2C-XGBnw | **1.656 ± 0.061** | **1.659 ± 0.045** | 2.744 ± 0.869 | 9.544 ± 0.204 |
| L2C-FNN | 1.678 ± 0.097 | 1.833 ± 0.330 | **1.763 ± 0.158** | 10.324 ± 0.130 |
| MACC | | | | |
| MinimalRNN | 3.135 ± 0.714 | 2.898 ± 0.686 | 2.288 ± 0.221 | 12.048 ± 0.645 |
| AD-Map | 2.659 ± 0.060 | N.A. | 2.424 ± 0.084 | 10.375 ± 0.468 |
| L2C-XGBw | 3.752 ± 0.641 | 3.510 ± 0.659 | 6.819 ± 1.814 | N.A. |
| L2C-XGBnw | 2.917 ± 0.354 | 2.731 ± 0.317 | 3.730 ± 1.010 | **9.201 ± 0.357** |
| L2C-FNN | **2.426 ± 0.226** | **2.709 ± 0.544** | **2.270 ± 0.234** | 9.734 ± 0.111 |
| OASIS | | | | |
| MinimalRNN | 2.335 ± 0.343 | 2.315 ± 0.360 | 2.600 ± 0.473 | 16.857 ± 2.141 |
| AD-Map | 2.261 ± 0.065 | N.A. | 2.294 ± 0.066 | **7.547 ± 0.146** |
| L2C-XGBw | 2.433 ± 0.162 | 2.401 ± 0.179 | 4.700 ± 1.272 | N.A. |
| L2C-XGBnw | 2.167 ± 0.092 | **2.113 ± 0.093** | 3.695 ± 0.721 | 8.910 ± 0.489 |
| L2C-FNN | **1.985 ± 0.032** | 2.114 ± 0.120 | **2.069 ± 0.081** | 10.288 ± 0.130 |

Table S22. Cross-cohort clinical diagnosis prediction performance under different modality ablation scenarios. Results were averaged across 20 trained models (obtained from training with all data in ADNI). Higher mAUC indicates better performance. The best result for each ablation scenario on each test dataset was bolded. Due to model design, AD-Map does not utilize diagnostic input features and therefore has no results in the “Ablate diagnosis” condition. On the other hand, L2C-XGBw relies on “time since most recent diagnosis” to select the appropriate XGBoost model, and this information is unavailable when diagnostic features are ablated. Therefore, these cases are marked as “N.A.”

| mAUC ↑ | No ablation | Ablate diagnosis | Ablate cognition | Ablate MRI |
| --- | --- | --- | --- | --- |
| AIBL | | | | |
| MinimalRNN | 0.874 ± 0.017 | 0.794 ± 0.050 | 0.838 ± 0.046 | 0.884 ± 0.017 |
| AD-Map | 0.834 ± 0.004 | N.A. | 0.663 ± 0.024 | 0.847 ± 0.002 |
| L2C-XGBw | 0.890 ± 0.013 | N.A. | 0.862 ± 0.014 | 0.813 ± 0.038 |
| L2C-XGBnw | 0.896 ± 0.010 | 0.817 ± 0.018 | 0.851 ± 0.009 | 0.876 ± 0.023 |
| L2C-FNN | **0.909 ± 0.005** | **0.874 ± 0.018** | **0.884 ± 0.008** | **0.916 ± 0.002** |
| MACC | | | | |
| MinimalRNN | 0.906 ± 0.029 | 0.828 ± 0.037 | 0.904 ± 0.043 | 0.910 ± 0.034 |
| AD-Map | 0.836 ± 0.006 | N.A. | 0.741 ± 0.028 | 0.829 ± 0.003 |
| L2C-XGBw | **0.960 ± 0.004** | N.A. | 0.946 ± 0.005 | 0.938 ± 0.017 |
| L2C-XGBnw | 0.959 ± 0.004 | 0.827 ± 0.013 | 0.942 ± 0.005 | 0.954 ± 0.012 |
| L2C-FNN | 0.958 ± 0.003 | **0.860 ± 0.006** | **0.952 ± 0.005** | **0.958 ± 0.003** |
| OASIS | | | | |
| MinimalRNN | 0.754 ± 0.023 | 0.705 ± 0.035 | 0.693 ± 0.036 | 0.752 ± 0.031 |
| AD-Map | 0.744 ± 0.006 | N.A. | 0.660 ± 0.014 | 0.733 ± 0.002 |
| L2C-XGBw | 0.798 ± 0.004 | N.A. | 0.747 ± 0.010 | 0.721 ± 0.029 |
| L2C-XGBnw | **0.804 ± 0.005** | 0.748 ± 0.009 | 0.758 ± 0.010 | 0.761 ± 0.027 |
| L2C-FNN | 0.803 ± 0.005 | **0.785 ± 0.008** | **0.782 ± 0.009** | **0.793 ± 0.006** |

Table S23. Test performance of L2C-FNN under different modality ablation scenarios. Results were averaged across 20 trained models (training with all modalities in ADNI). For each column, the ablated modality that caused the largest drop in performance was bolded.

|  | MMSE MAE ↓ | | | Ventricle MAE (1e-3) ↓ | | | Diagnostic mAUC ↑ | | |
| --- | --- | --- | --- | --- | --- | --- | --- | --- | --- |
|  | AIBL | MACC | OASIS | AIBL | MACC | OASIS | AIBL | MACC | OASIS |
| No Ablation | 1.330 ± 0.040 | 2.672 ± 0.150 | 1.387 ± 0.016 | 1.678 ± 0.097 | 2.426 ± 0.226 | 1.985 ± 0.032 | 0.909 ± 0.005 | 0.958 ± 0.003 | 0.803 ± 0.005 |
| Ablate Diagnosis | 1.369 ± 0.048 | 2.707 ± 0.155 | 1.448 ± 0.018 | 1.833 ± 0.330 | 2.709 ± 0.544 | 2.114 ± 0.120 | **0.874 ± 0.018** | **0.860 ± 0.006** | 0.785 ± 0.008 |
| Ablate Cognition | **1.849 ± 0.041** | **6.809 ± 0.317** | **1.922 ± 0.053** | 1.763 ± 0.158 | 2.270 ± 0.234 | 2.069 ± 0.081 | 0.884 ± 0.008 | 0.952 ± 0.005 | **0.782 ± 0.009** |
| Ablate MRI | 1.354 ± 0.031 | 2.843 ± 0.203 | 1.442 ± 0.022 | **10.324 ± 0.130** | **9.734 ± 0.111** | **10.288 ± 0.130** | 0.916 ± 0.002 | 0.958 ± 0.003 | 0.793 ± 0.006 |

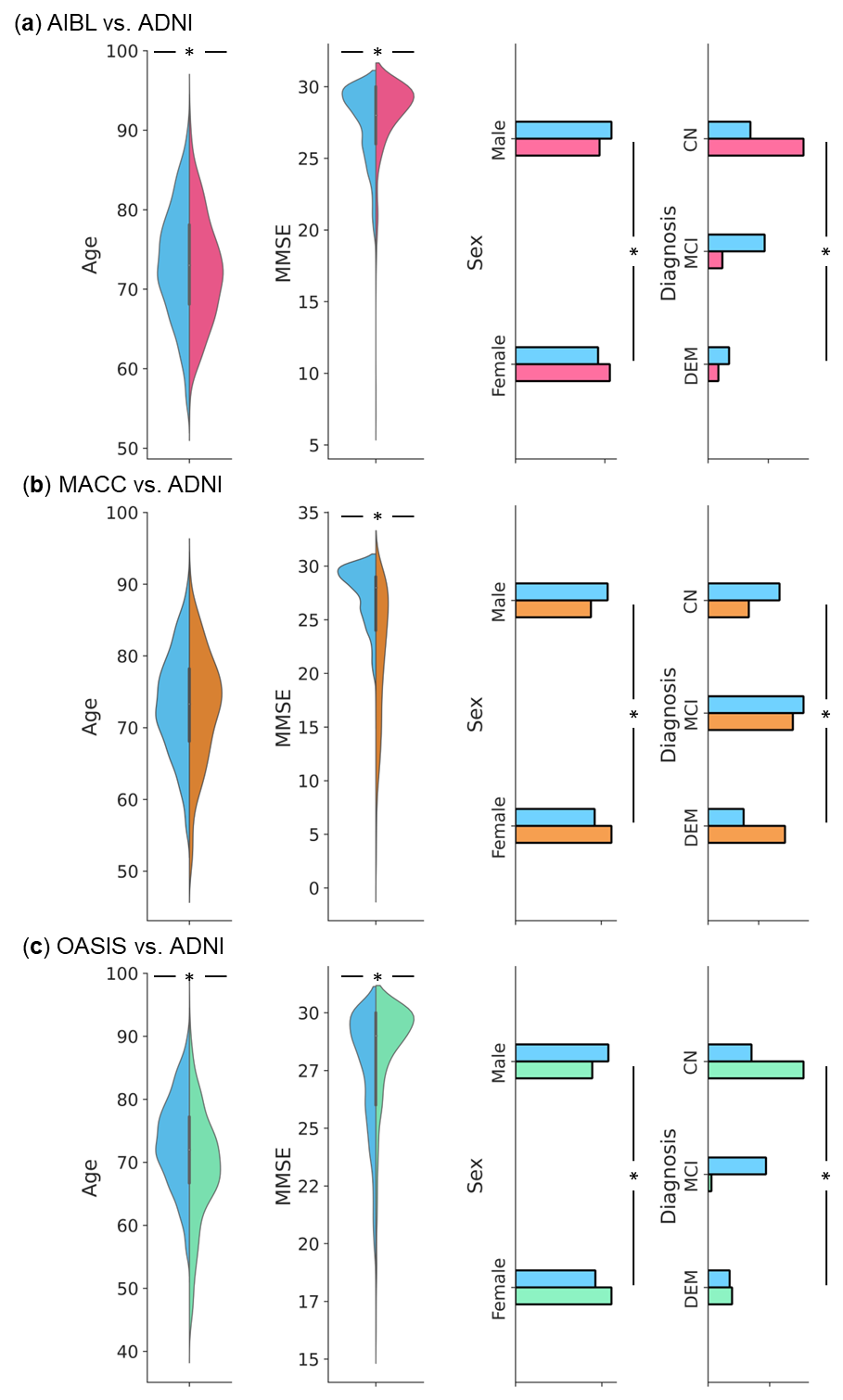

**Figure S1.** Baseline age, baseline MMSE, sex and baseline clinical diagnosis distribution differences between ADNI and external test set. (a) Distributions of age, sex, MMSE and clinical diagnosis for ADNI (blue) and AIBL (pink). (b) Distributions of age, sex, MMSE and clinical diagnosis for ADNI (blue) and MACC (yellow). (c) Distributions of age, sex, MMSE and clinical diagnosis for ADNI (blue) and OASIS (green). To test for differences in mean age and mean MMSE between ADNI and the external dataset, a permutation test was used. To test for differences in distributions of sex and diagnosis between ADNI and the external dataset, the chi square test was used. * indicates statistical significance after correcting for multiple comparisons with false discovery rate (FDR) q < 0.05. See Table 2 in the main text for the full set of statistical tests.

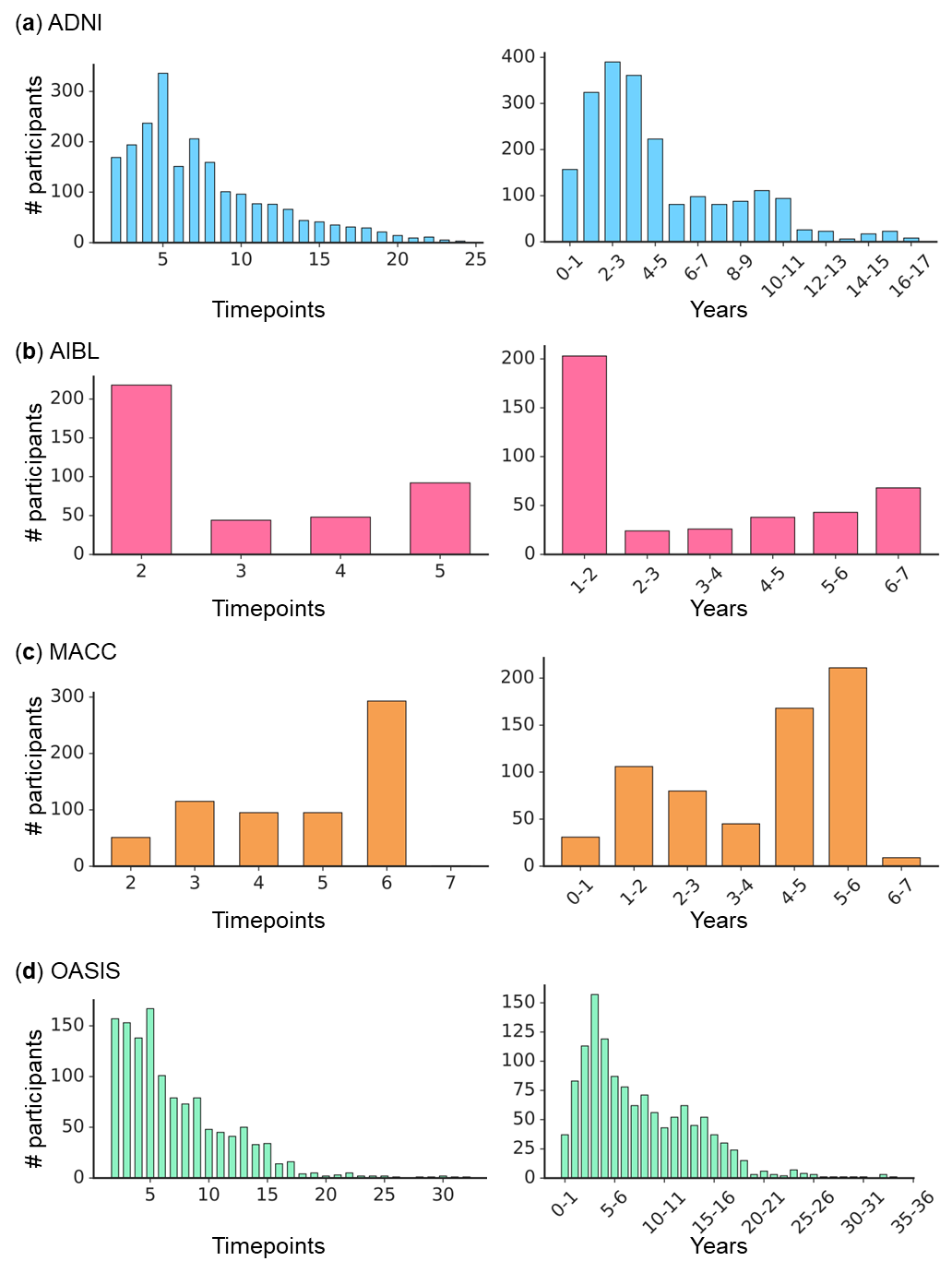

**Figure S2. Left:** Distribution of the number of timepoints for all participants in each dataset. **Right:** Distribution of the number of years between baseline and the last observation for all participants in each dataset. Note that year 0-1 means $0<t\leq12$, where $t$ is the interval between baseline and the last observed timepoint; year 1-2 means $12<t\leq24$, and so on. (a) ADNI. (b) AIBL. (c) MACC. (d) OASIS.

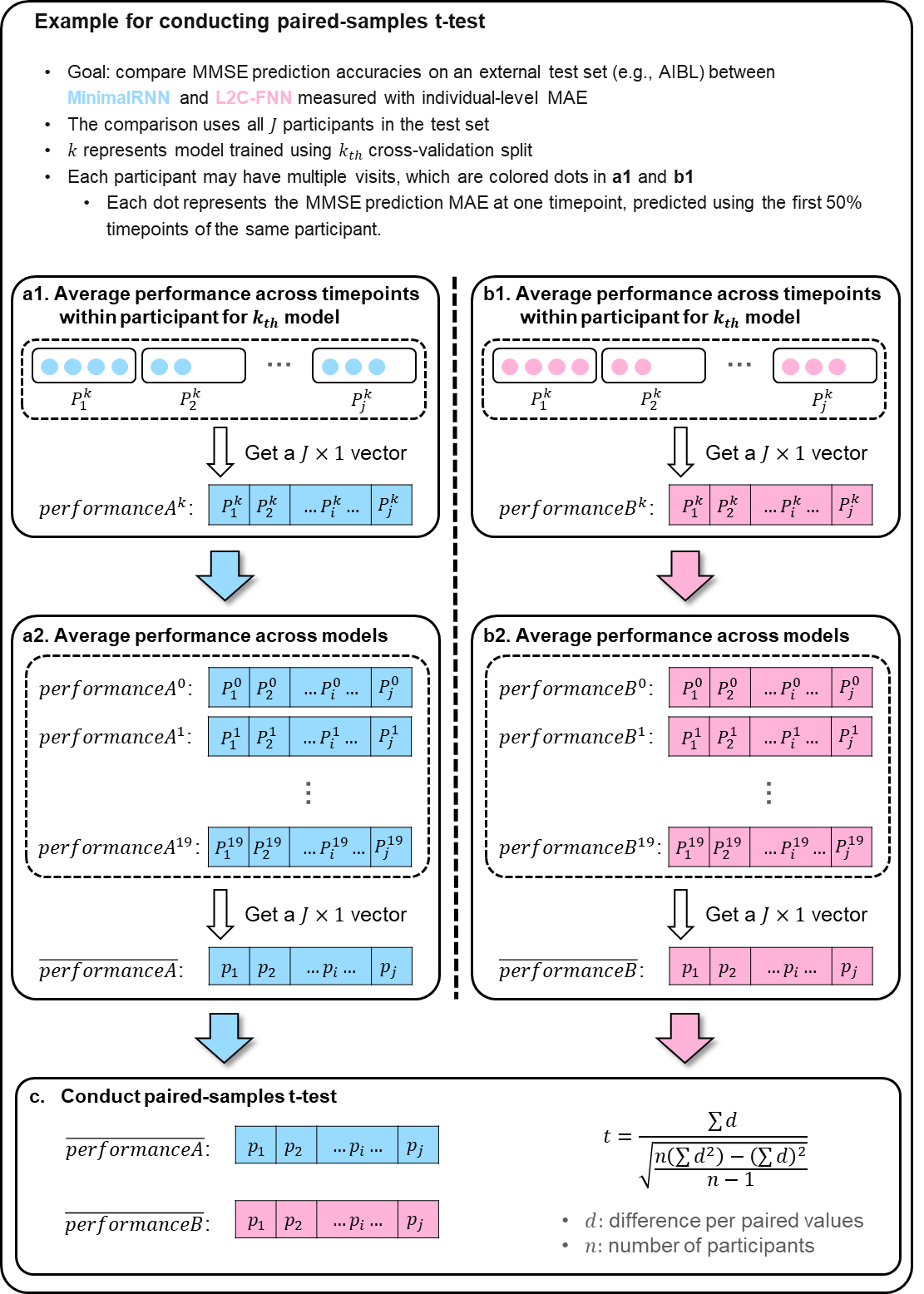

**Figure S3.** Illustration of paired-samples t-test for comparing performance measured with individual-level metrics (e.g., MMSE MAE) of MinimalRNN and L2C-FNN on an external test set. (a1) For a given model, we averaged the MMSE MAE within each participant across all timepoints for MinimalRNN. (a2) Averaging the participant-level MMSE MAE obtained in (a1) across the 20 models. (b1 & b2) Same as a1 and a2 but for L2C-FNN. (c) Conduct paired-samples t-test to obtain p value.

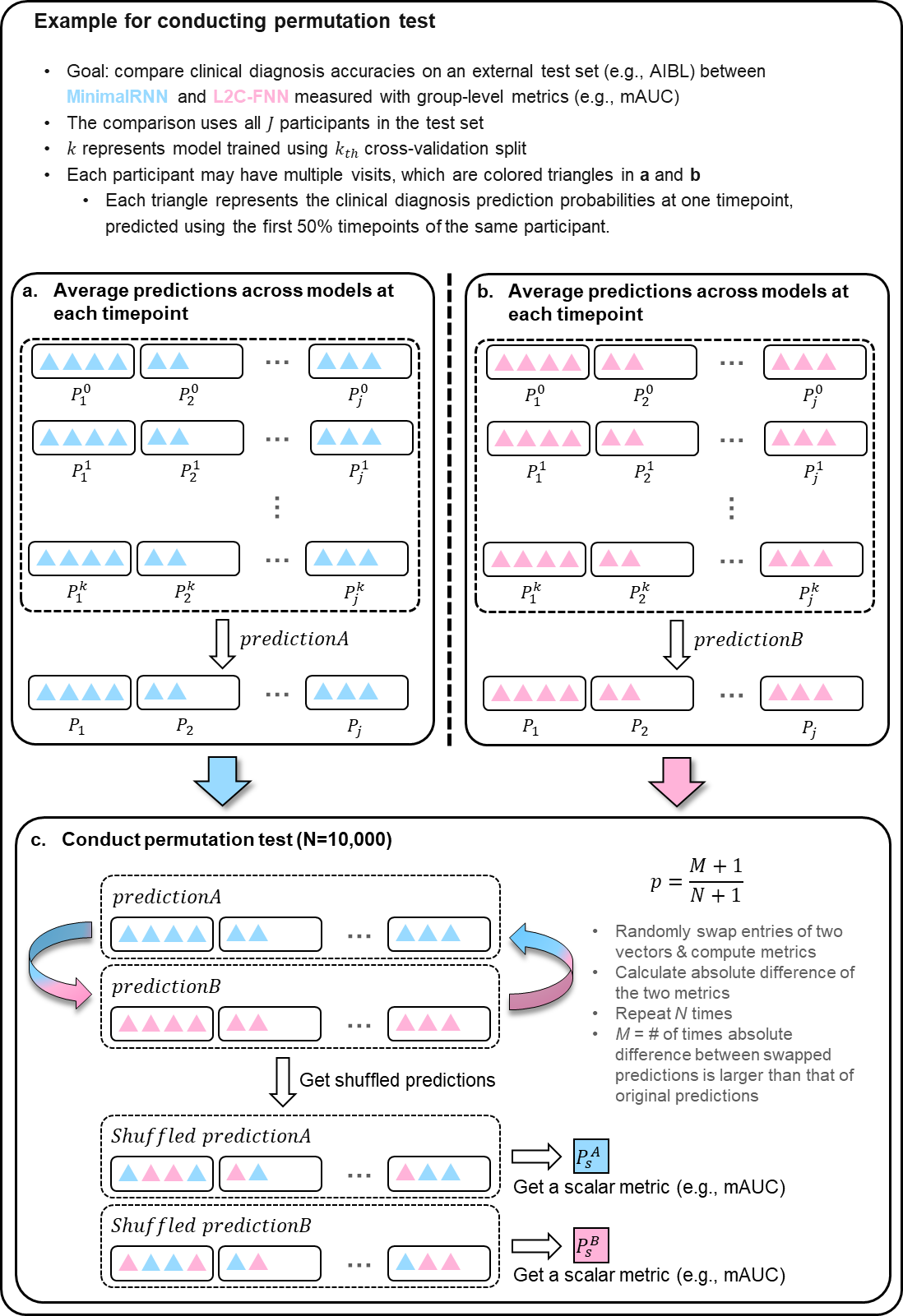

**Figure S4.** Illustration of permutation test for comparing performance measured with group-level metric (e.g., diagnostic prediction mAUC) of MinimalRNN and L2C-FNN on external test set. (a) For each participant, we averaged the predictions at each timepoint across 20 models for MinimalRNN. (b) Same as A but for L2C-FNN. (c) Permute 10,000 times and compute group-level metric (e.g., mAUC) on permuted predictions to obtain p value

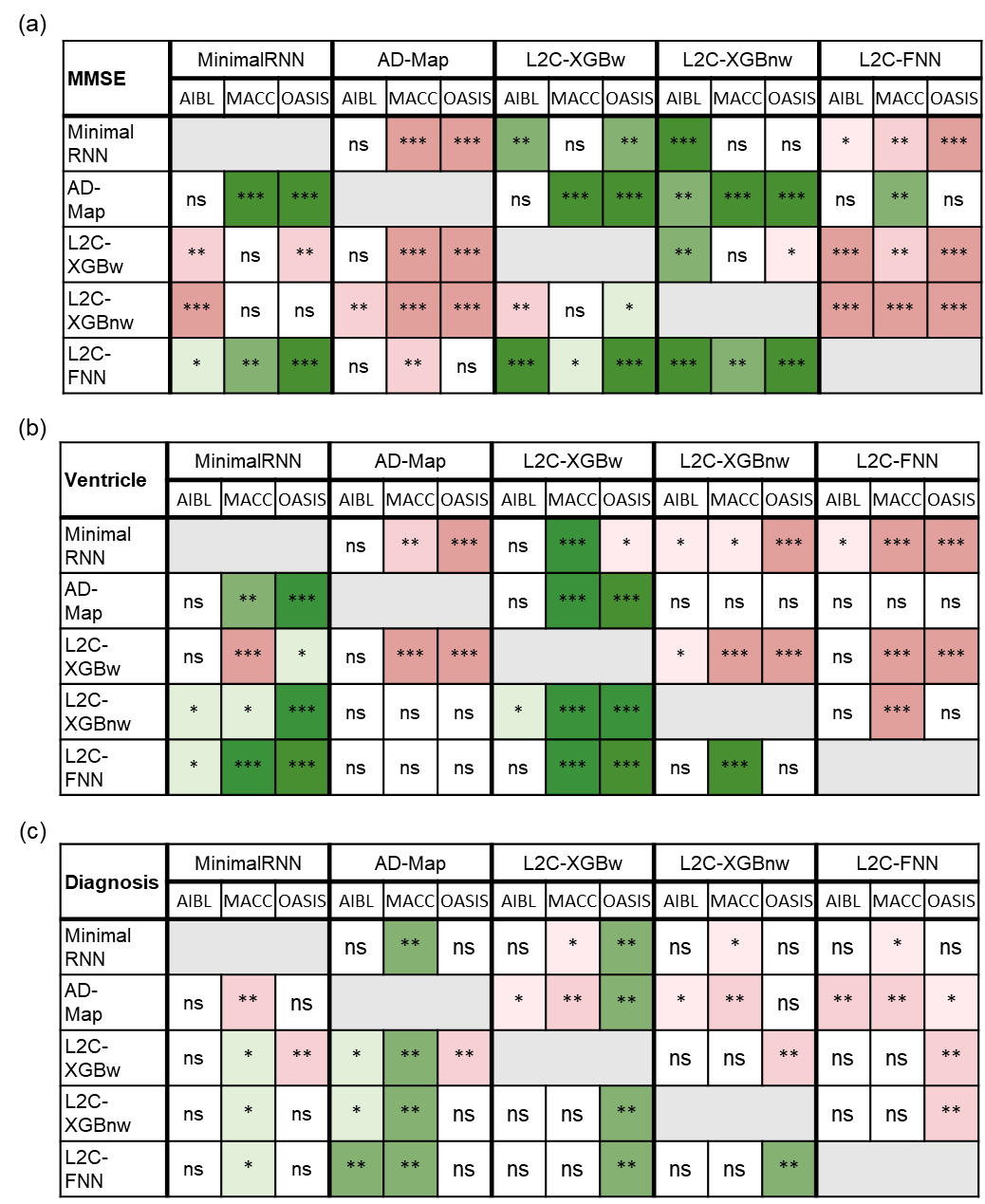

Figure S5. Statistical significance in the prediction performance between all models across the three external datasets for AD-only experiments (setting other dementia diagnosis to NaN). (a) Statistical significance between algorithms for MMSE prediction. Each row shows the statistical difference between a model and all other models across the three test datasets. The columns are grouped into five major columns corresponding to the five approaches. Within each major column, the three sub-columns represent results on AIBL, MACC, and OASIS datasets, respectively. For example, the first row corresponds to the statistical difference between MinimalRNN and the other models – green indicates that MinimalRNN performs better, while red indicates that MinimalRNN performs worse. “*” indicates p < 0.05 and statistical significance after multiple comparisons correction (FDR q < 0.05). “**” indicates p < 0.001 and statistical significance after multiple comparisons correction (FDR q < 0.05). “***” indicates p < 0.00001 and statistical significance after multiple comparisons correction (FDR q < 0.05). “ns” indicates no statistical significance (p ≥ 0.05) or did not survive FDR correction. (b) Same as (a) but for ventricular volume prediction. (c) Same as (a) but for clinical diagnosis prediction.

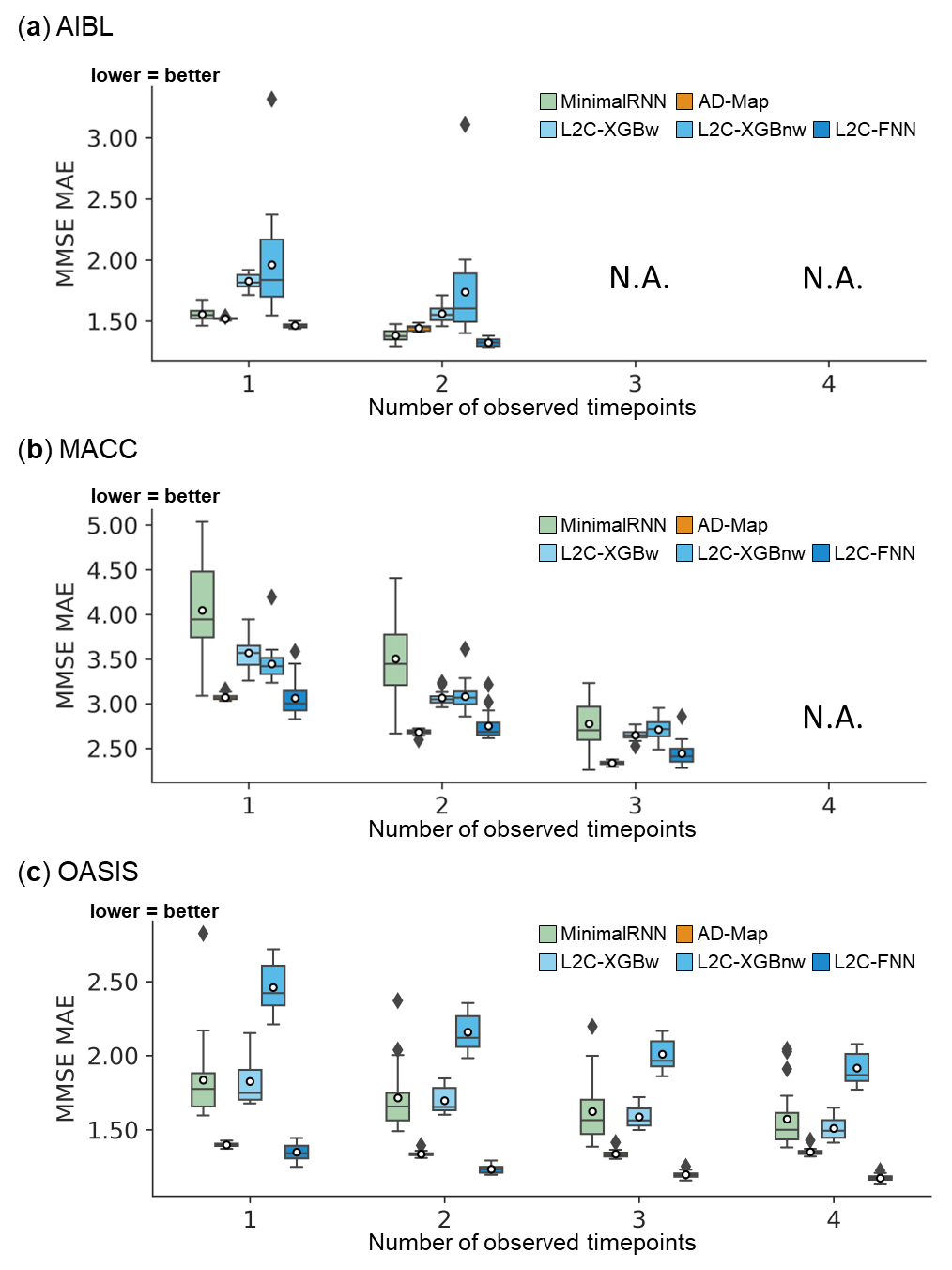

Figure S6. Cross-cohort MMSE prediction performance using different numbers of input timepoints (after training with all timepoints in ADNI). L2C-FNN compared favorably with respect to other approaches across three external test datasets. Results of statistical tests between L2C-FNN and other approaches are reported in Figure 6.

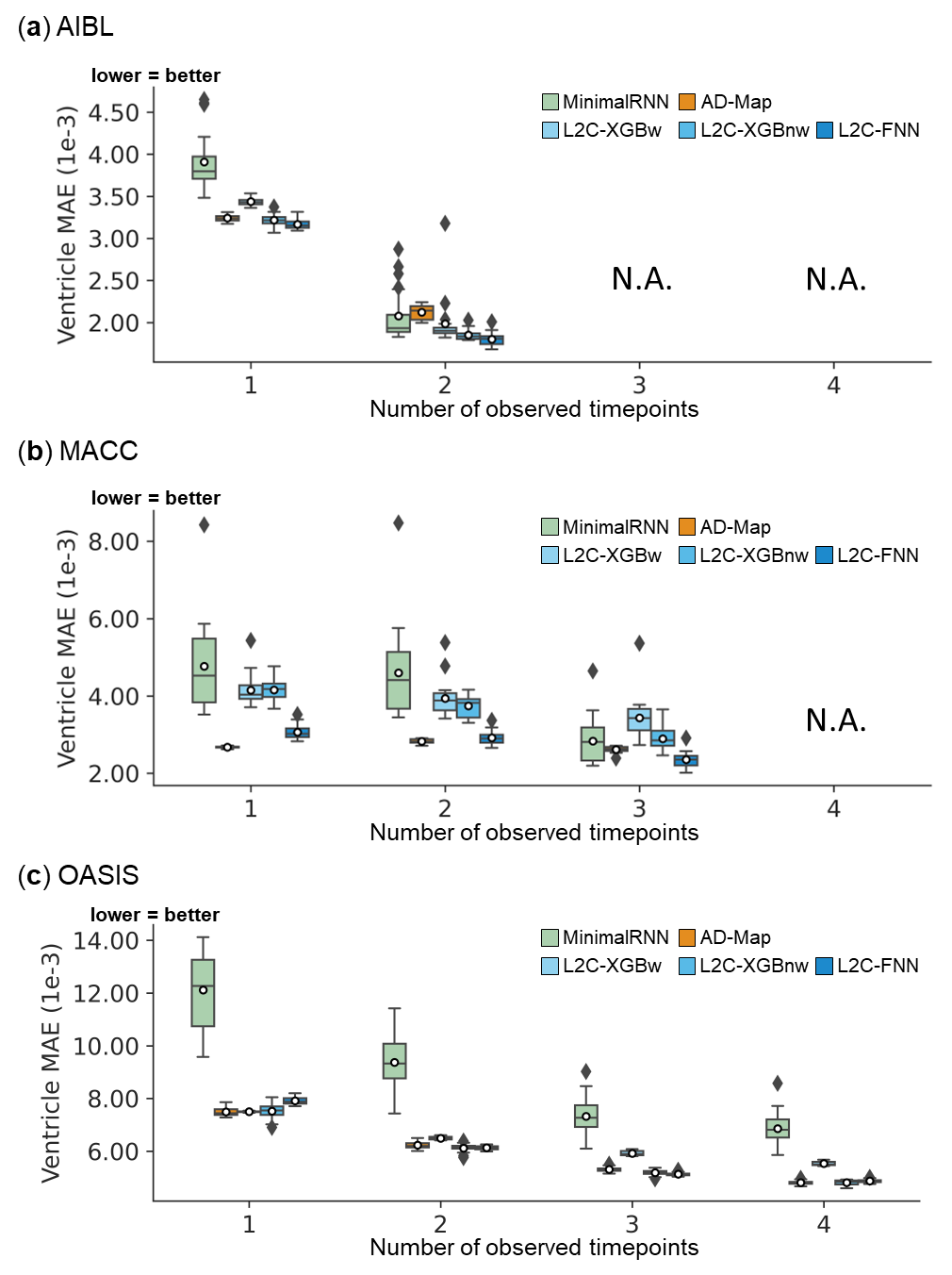

**Figure S7.** Cross-cohort ventricle volume prediction performance using different numbers of input timepoints (after training with all timepoints in ADNI). L2C-FNN compared favorably with respect to other approaches across three external test datasets, except L2C-XGBnw on OASIS using 1 input timepoint. Results of statistical tests between L2C-FNN and other approaches are reported in Figure 6.

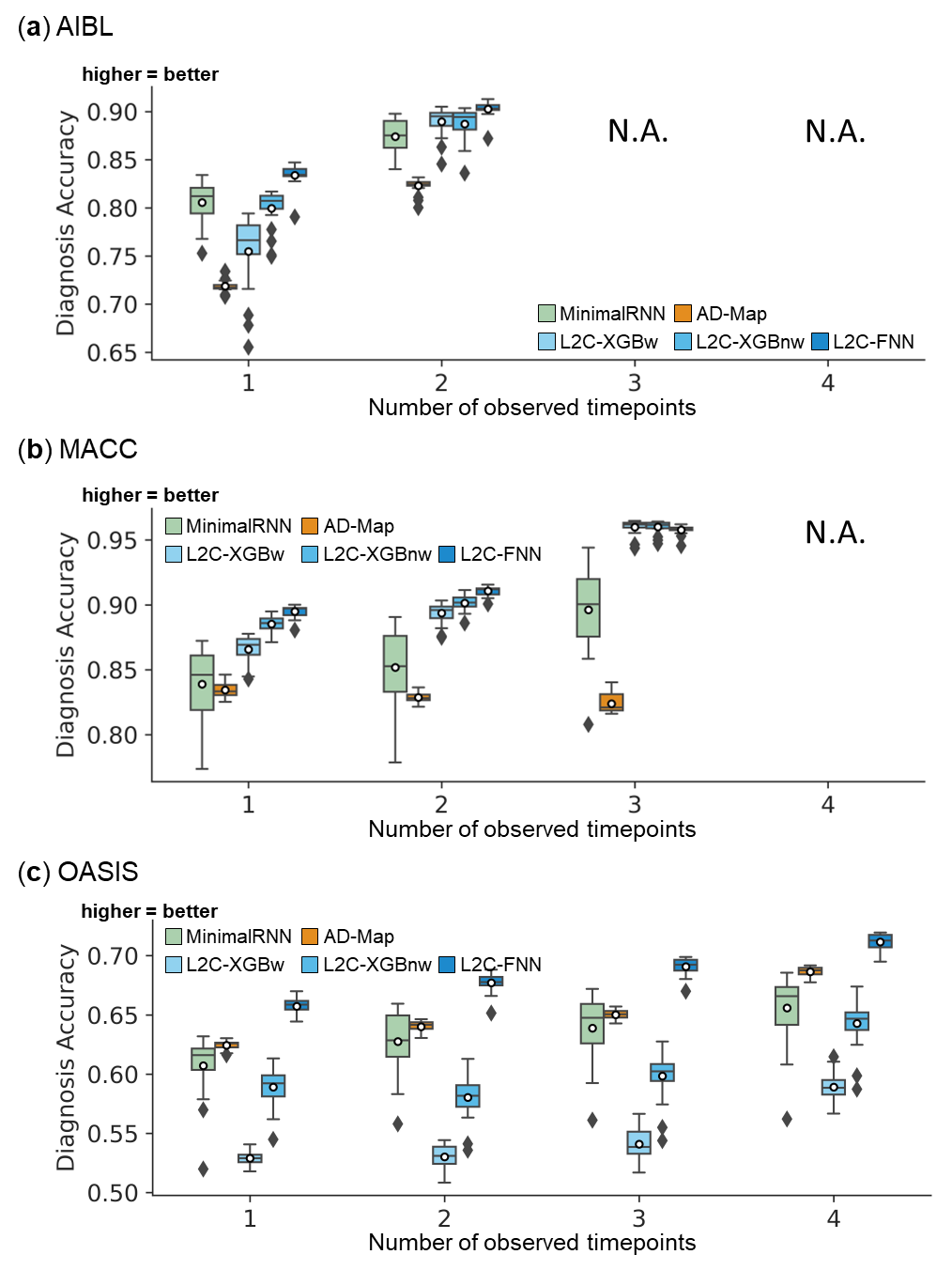

**Figure S8.** Cross-cohort clinical diagnosis prediction performance using different numbers of input timepoints (after training with all timepoints in ADNI). L2C-FNN significantly outperformed almost all other approaches across three external test datasets using different number of input timepoints. Results of statistical tests between L2C-FNN and other approaches are reported in Figure 6.

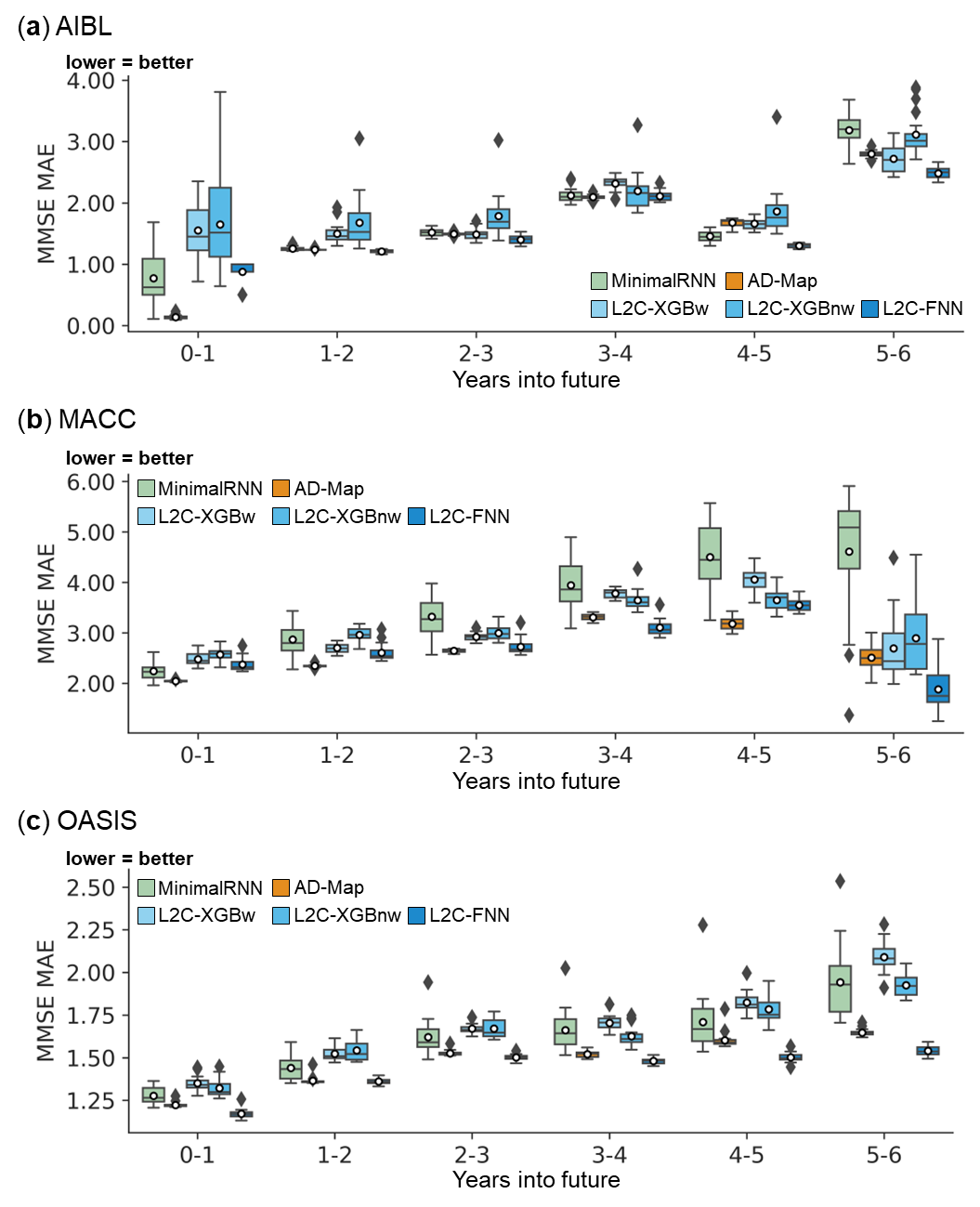

**Figure S9.** Cross-cohort MMSE prediction performance broken down into yearly intervals up to 6 years into the future. Note that the last observed time point is at month 0, so year 0-1 means that the prediction was for a future observation at 0 < month ≤ 12, year 1-2 means that the prediction was for a future observation at 12 < month ≤ 24, etc. All algorithms became worse further into the future. L2C-FNN was comparable to or better than all models across all years in three external test datasets except AD-Map in MACC for years 0-1 and 1-2. Results of statistical tests between L2C-FNN and other approaches are reported in Figure 7.

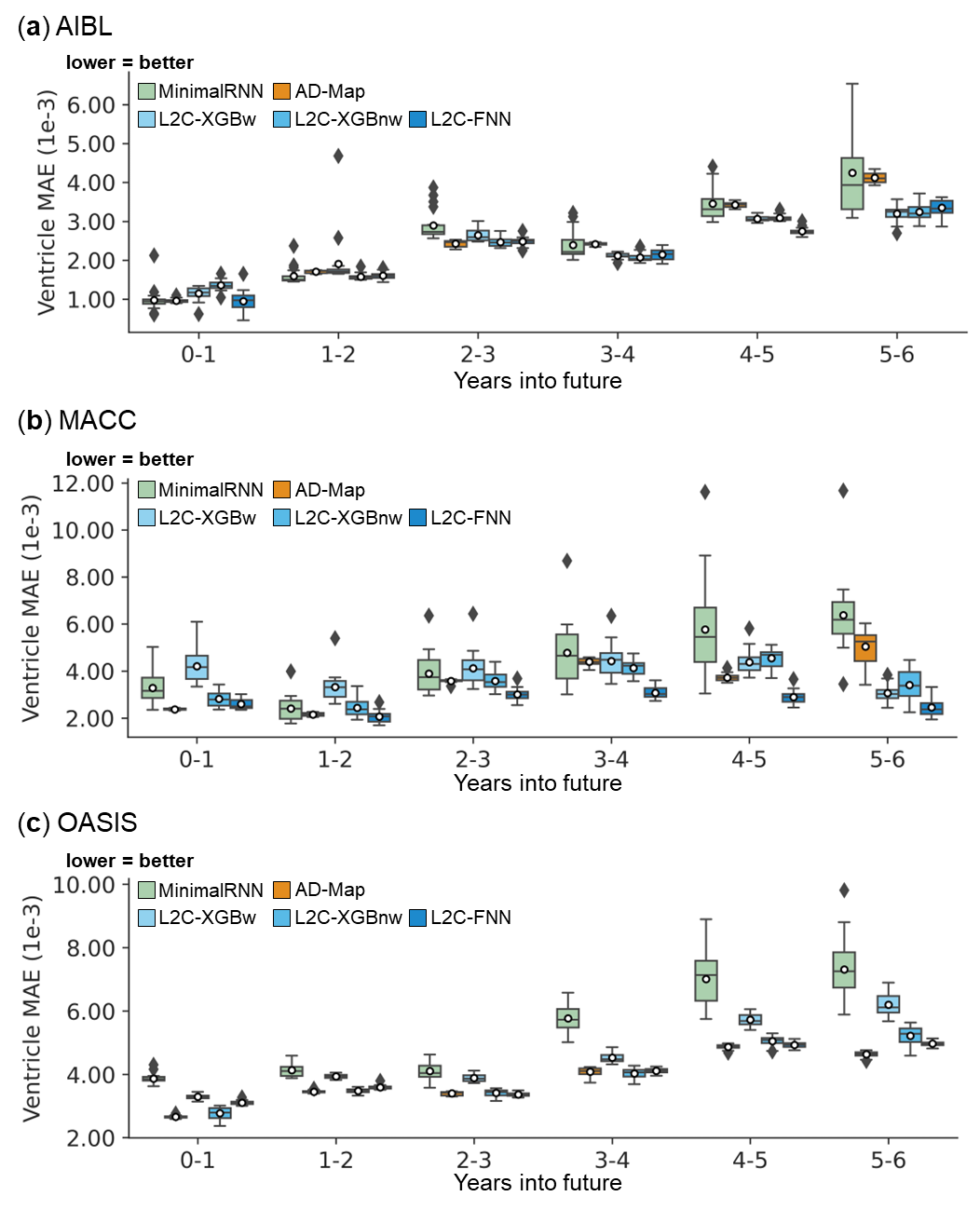

**Figure S10.** Cross-cohort ventricle volume prediction performance broken down into yearly intervals up to 6 years into the future. Note that the last observed time point is at month 0, so year 0-1 means that the prediction was for a future observation at 0 < month ≤ 12, year 1-2 means that the prediction was for a future observation at 12 < month ≤ 24, etc. All algorithms became worse further into the future. L2C-FNN was comparable to or better than all models across all years in three external test datasets. Results of statistical tests between L2C-FNN and other approaches are reported in Figure 7.

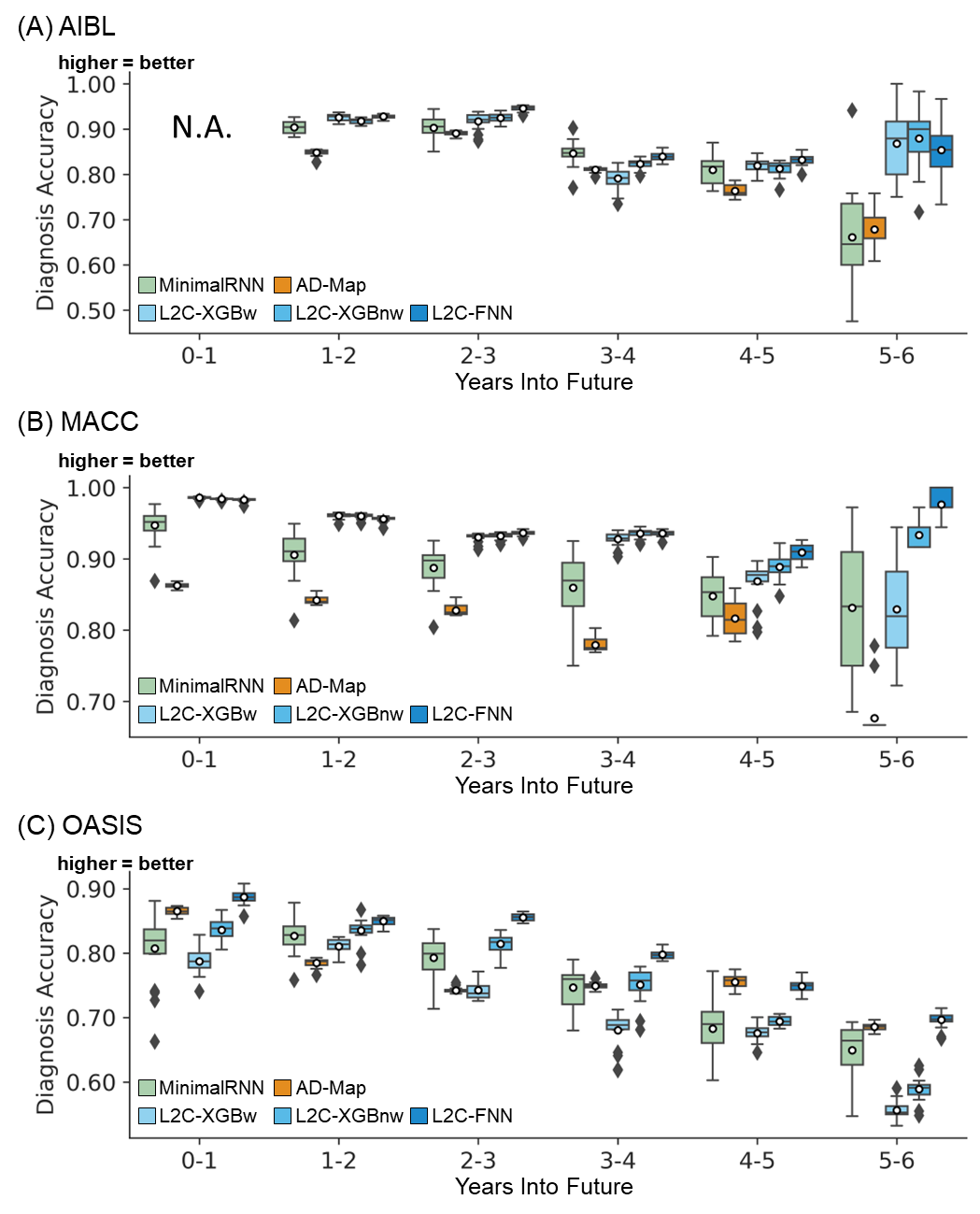

**Figure S11.** Cross-cohort clinical diagnosis prediction performance broken down into yearly intervals up to 6 years into the future. Note that the last observed time point is at month 0, so year 0-1 means that the prediction was for a future observation at 0 < month ≤ 12, year 1-2 means that the prediction was for a future observation at 12 < month ≤ 24, etc. All algorithms became worse further into the future. L2C-FNN was comparable to or better than all models across all years in three external test datasets. Due to dataset constraints, AIBL includes only one diagnostic class in year 0-1, making mAUC undefined in this case. Therefore, results for AIBL at year 0-1 is marked as “N.A.” Results of statistical tests between L2C-FNN and other approaches are reported in Figure 7.

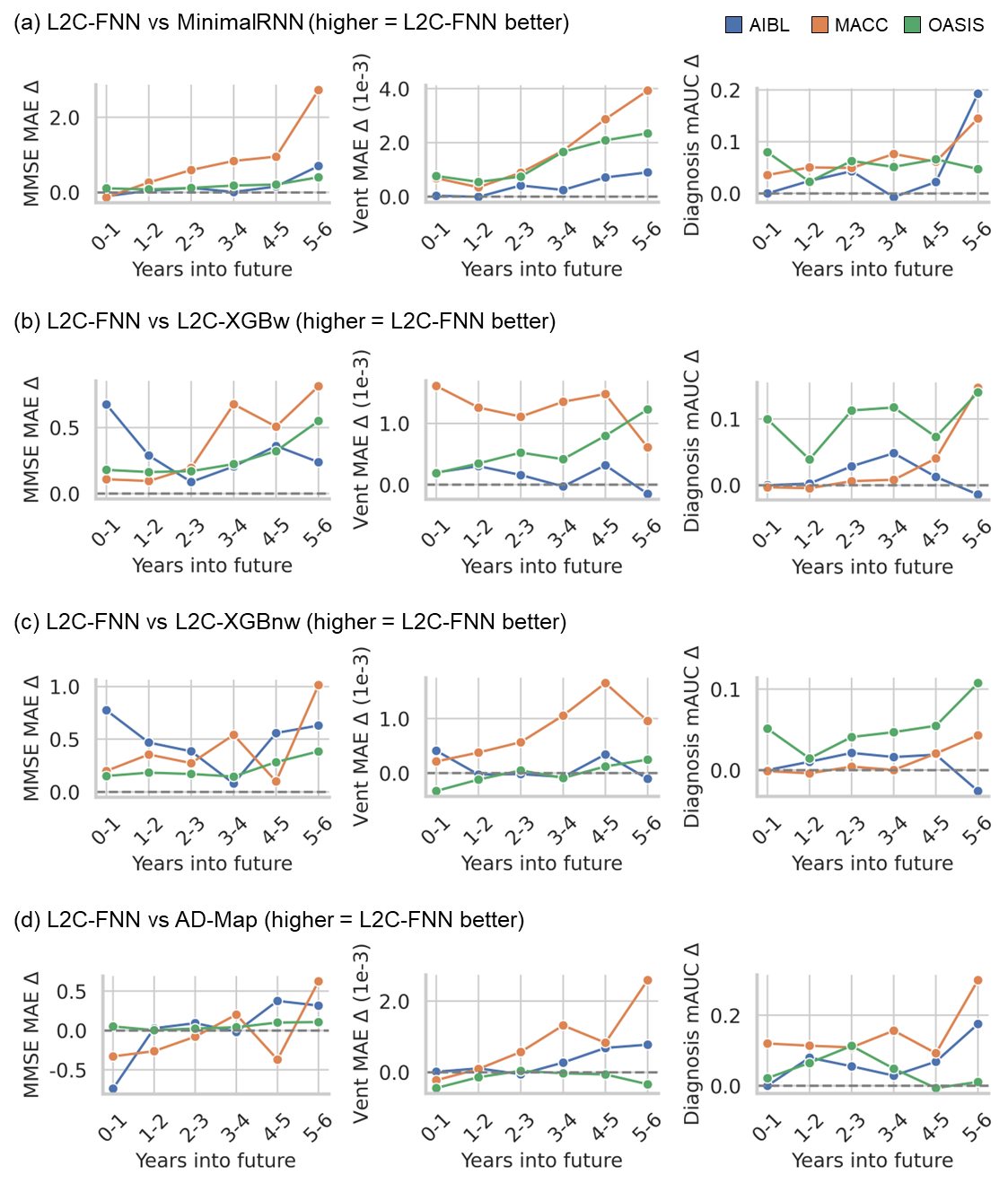

**Figure S12.** Relative performance of L2C-FNN versus the models at yearly intervals up to 6 years into the future. (a) Here we compared L2C-FNN versus minimalRNN. Left panel shows MinimalRNN MMSE MAE minus L2C-FNN MMSE MAE, so a positive value indicates that L2C-FNN performs better. Middle panel shows MinimalRNN ventricle volume MAE minus L2C-FNN ventricle volume MAE, so a positive value indicates that L2C-FNN performs better. Right panel shows L2C-FNN diagnosis mAUC minus MinimalRNN diagnosis mAUC, so a positive value indicates that L2C-FNN performs better. (b) same as (a) but showing L2C-FNN versus L2C-XGBw. (c) same as (a) but showing L2C-FNN versus L2C-XGBnw. (d) Same as panel (a) but showing L2C-FNN versus AD-Map. As shown in panel (a), compared with MinimalRNN, the advantage of L2C-FNN generally increased further into the future. On the other hand, there was not an obvious trend when comparing L2C-FNN and other models.
